# Alcohol use and risk of dementia in diverse populations

**DOI:** 10.1101/2024.05.20.24307606

**Authors:** Anya Topiwala, Daniel F. Levey, Hang Zhou, Joseph D. Deak, Keyrun Adhikari, Klaus P. Ebmeier, Steven Bell, Stephen Burgess, Thomas E. Nichols, J. Michael Gaziano, Murray B Stein, Joel Gelernter

## Abstract

**Objective:** To examine the causal relationship between alcohol use and dementia risk across multiple ancestry groups.

**Design:** We triangulated evidence from observational and univariable and multivariable Mendelian randomization.

**Setting and participants:** Cross-ancestry observational analyses were conducted in two large prospective studies: the US Million Veteran Program and UK Biobank. One- and two-sample univariable and multivariable Mendelian randomization used de novo data from a genome-wide association study in Million Veteran Program plus publicly available data.

**Main outcome measure:** All-cause dementia.

**Results:** Among 559,559 participants (aged 56-72 years old at baseline) included in observational analyses, 14,540 received developed dementia and 48,034 died during follow-up. Observational associations between alcohol and dementia were U-shaped. Non-, heavy (>40 drinks per week - hazard ratio (HR)1.41; 95% confidence interval [CI], 1.15 to 1.74]) and dependent (1.51[1.42-1.60]) drinkers were at higher dementia risk than light drinkers. In contrast, genetic analyses revealed a monotonically increasing association between alcohol dose and dementia, with no evidence supporting a protective effect of any level of drinking. A two-fold increase in genetically-predicted alcohol use disorder prevalence was associated with a 16% increase in dementia cases (IVW OR=1.16[1.03-1.30]), and a one standard deviation increase in log-transformed drinks per week was associated with a 15% increase (IVW OR=1.15[1.03-1.27]).

**Conclusions:** Alcohol consumption has a causal role for dementia. These findings challenge a purported protective effect of moderate drinking. Reducing alcohol use could be an effective dementia prevention strategy.

## Introduction

Alcohol consumption is widespread, modifiable, and associated with many medical harms, but whether there is a causal link with dementia remains contentious. Heavy drinking has been associated with increased dementia risk in some cohorts but not others.^1^ In moderate drinkers the picture is even less clear, with some studies purporting protective effects.^2^ However, recent neuroimaging studies have found adverse associations with dementia endophenotypes even at low alcohol consumption levels.^3^ Clarifying the impact of alcohol on dementia is of critical importance for public health.

Methodological differences may explain contradictions between existing studies.^4^ Most studies have included few heavy or dependent drinkers, limiting power.^4^ Many have involved elderly participants whose cognitive impairment might affect drinking rather than the reverse.^5^ Nondrinking reference groups likely included now-abstinent former heavy drinkers,^6^ so that confounding is a concern. Therefore, at present, we cannot confidently infer causality.^7^ Practical and ethical issues preclude a randomized controlled trial, which require a presumption of clinical equipoise. Under certain conditions, causal effects may be estimated from observational data. One approach is Mendelian randomization, a quasi-experimental method that can estimate causal effects via genetics.^8^ To date, there have been four published Mendelian randomization studies investigating alcohol and dementia, all reporting null findings. However, they have had limited power and scope, and solely examined associations with late-onset Alzheimer’s disease in European-ancestry populations,^9^ whereas in the context of alcohol use non-Alzheimer etiologies are also highly relevant.^10^

Here we sought to estimate the dose-response relationship of alcohol use with dementia. To strengthen causal inference, we triangulated traditional observational and genetic analyses and examined different dementia subtypes, alcohol-related traits, and genetic ancestries. The objective of our study was to evaluate the causal role of alcohol in dementia.

## Methods

### Study populations

The observational study included participants with complete phenotype data (Figure S1) from Million Veteran Program (MVP), which recruited US veterans from 2011 to the present,^11^ and from UK Biobank (UKB), which recruited volunteers aged 40-69 years from 2006 to 2010.^12^ Participants were unrelated and stratified by genetic ancestry. Individuals gave written informed consent, and studies had approval from their respective institutional and ethics review boards. Participants were followed up from recruitment until either their first dementia diagnosis, death, or the date of last follow up (December 2019 for MVP and January 2022 for UKB). The genetic studies included 2.4 million participants of European and African ancestry, comprising the 45 genome-wide association study cohorts (Table S1).

### Exposure measurement

Alcohol intake was characterized by self-reported drinks per week (DPW), derived from questionnaires or the AUDIT-C clinical screening tool (scored 0-12) (Supplementary Appendix). To limit reverse causation, the earliest recorded intake was used in primary analyses where possible. Never and former drinkers were distinguished where recorded. Alcohol use disorder cases were identified using diagnostic codes in the linked electronic health record (EHR). Beverage type and binge-pattern consumption (>6 drinks on one occasion) were ascertained from surveys and AUDIT-C questions. For genetic analyses, problematic alcohol use was defined by meta-analyzing alcohol use disorder and AUDIT-P.^13^

### Outcome measurement

Given uncertainty about whether specific dementia subtypes are impacted by alcohol, our primary outcome was all-cause dementia. Cases were identified using EHR ICD codes (Table S2). Prevalent cases (those with dementia at enrollment) were excluded from observational analyses to reduce the risk of reverse causation. As this was not a concern for genetic analyses, both incident and prevalent cases were included. Secondary outcomes were Alzheimer’s and vascular dementia, respectively.

### Covariates

Potential confounders and effect modifiers were identified from published literature (Supplementary Appendix). Self-administered questionnaires were used to collect baseline information on demographic factors (age, sex, education, income), lifestyle behaviors (smoking), and physical and psychiatric health (body mass index, head injury, post-traumatic stress disorder, diabetes). Mean blood pressure, substance use disorders, and thiamine levels (deficiency leads to alcohol-related brain harm)^14^ were determined using the EHR in MVP. Higher order age terms (age^2^ and age^3^) and age by sex interactions were included in models.

### Genetic variants

Genetic variants associated with alcohol use in the largest available previous studies at the genome-wide significant level (p<5x10^-8^) were applied as instrumental variables (Tables S3-6). Where possible, variants were identified in separate ancestry groups, using a less stringent p value threshold (p<5x10^-5^) when few variants were available (Table S4). Ancestry-specific linkage disequilibrium clumping was performed to ensure variant independence (Supplementary Appendix).

Dementia phenotype definition determines interpretation of Mendelian randomization estimates.^15^ We calculated genetic associations with all-cause dementia in 451,317 (25,473 cases) European and 114,238 (5,706 cases) African ancestry individuals in MVP (Supplementary Appendix). Associations with related phenotypes, proxy dementia (defined as parental history of Alzheimer’s or dementia) and clinically-diagnosed Alzheimer’s disease were also examined (Table S1). Proxy genetic variants were sought when not present in the outcome data.

### Statistical analyses

For observational analysis, Cox proportional hazards models were used (Supplementary Appendix). Light drinkers were used as the reference group because some current nondrinkers previously drank heavily and have reduced alcohol intake because of health concerns (‘sick quitters’). Follow-up length was defined as the interval between enrollment (when covariates were measured) and either first date of diagnosis or censoring (at death or last data collection). Random effects meta-analysis generated pooled effect sizes across comparable ancestry cohorts. Sensitivity analyses in MVP included examining: the competing risk of death using the subdistribution method, nonlinear relationships using restricted cubic splines, controlling for additional potential confounds, and different ancestry groups. The influence of potential modifiers of alcohol-dementia relationships were explored using interaction terms. These included: binge-drinking frequency, sex,^16^ *ADH1B\**rs1229984 (affecting rate of alcohol metabolism)^17^ and *ApoE4* genotypes.^18^ In MVP, participants had numerous AUDIT-C scores available, allowing examination of how drinking behaviors changed over the time preceding dementia diagnosis, relevant to reverse causation (Supplementary Appendix).

For the genetic analyses, associations with all-cause dementia were calculated in a genome-wide association study (Supplementary Appendix). The inverse variance weighted (IVW) method was used as the main linear Mendelian randomization analysis, with several robust methods used as sensitivity analyses.

Additionally, reverse Mendelian randomization was used to estimate effects of dementia on alcohol use, and multivariable analyses used to test whether causal effects of alcohol on dementia were altered by adjustment for key potential confounds or mediators. Nonlinear analysis of five exposure strata were performed to assess for potential nonlinear effects of alcohol on dementia, using individual-level data on unrelated participants of European ancestry in MVP (n=313,873, including 16,932 dementia cases), which had greater power than UKB for nonlinear analyses. Sensitivity analyses excluded current nondrinkers, to test the independence assumption in Mendelian randomization, and used age and sex as negative controls to assess for possible bias in nonlinear analyses.^19^ Analyses were performed in R (v4.1.2), unless otherwise stated.

### Patients and public involvement

A public engagement group, Friends of Oxford Dementia and Ageing Research, was consulted by the lead author (AT) early in the development of the research idea.

## Results

### Observational associations between alcohol use and dementia

559,559 participants were included in observational analyses, of whom 10,564 developed incident all-cause dementia over follow-up in MVP (mean=4.3 years, median=4.4 years, maximum=9.0 years), and 3976 in UKB (mean=12.4 years, median=12.7 years, maximum=15.0 years), consistent with a lower mean age in UKB (Table 1). 28,738 died during follow-up in MVP and 19,296 in UKB. Over 90% of participants in both cohorts reported consuming alcohol at first measurement. Compared to drinkers, nondrinker groups were older, with a higher proportion of females and had lower educational qualifications.

**Table 1:**
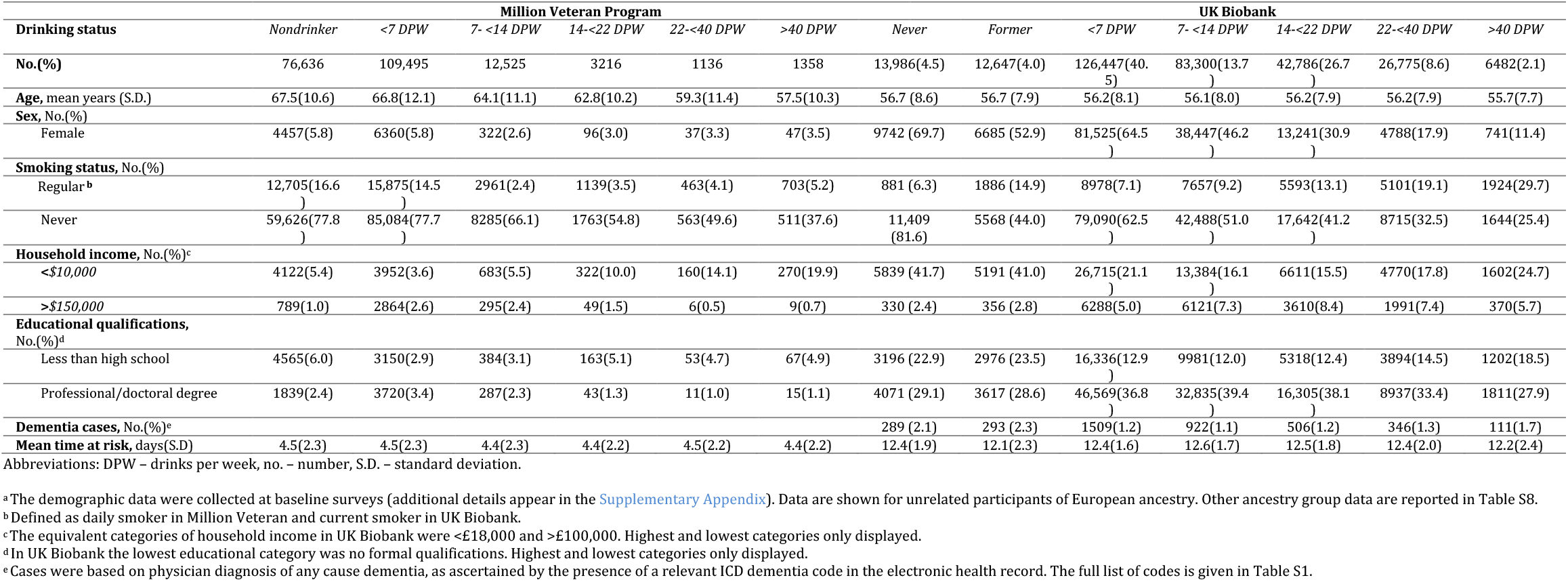
Characteristics Of Unrelated European Ancestry Participants in Million Veteran Program and UK Biobank According to Earliest Recorded Alcohol Intake^a^.

Conventional observational analyses found a U-shaped relationship between self-reported alcohol intake and incident dementia (Figure 1 & Figure S2). Nondrinkers (irrespective of subdivision into never and former drinkers), heavy (>40 DPW) and dependent drinkers (HR=1.51[1.42-1.60]) had higher incident all-cause dementia compared with light drinkers (<7 DPW). In UKB, moderate drinkers (7-14 DPW) had lower dementia incidence than light drinkers.

**Figure 1:**
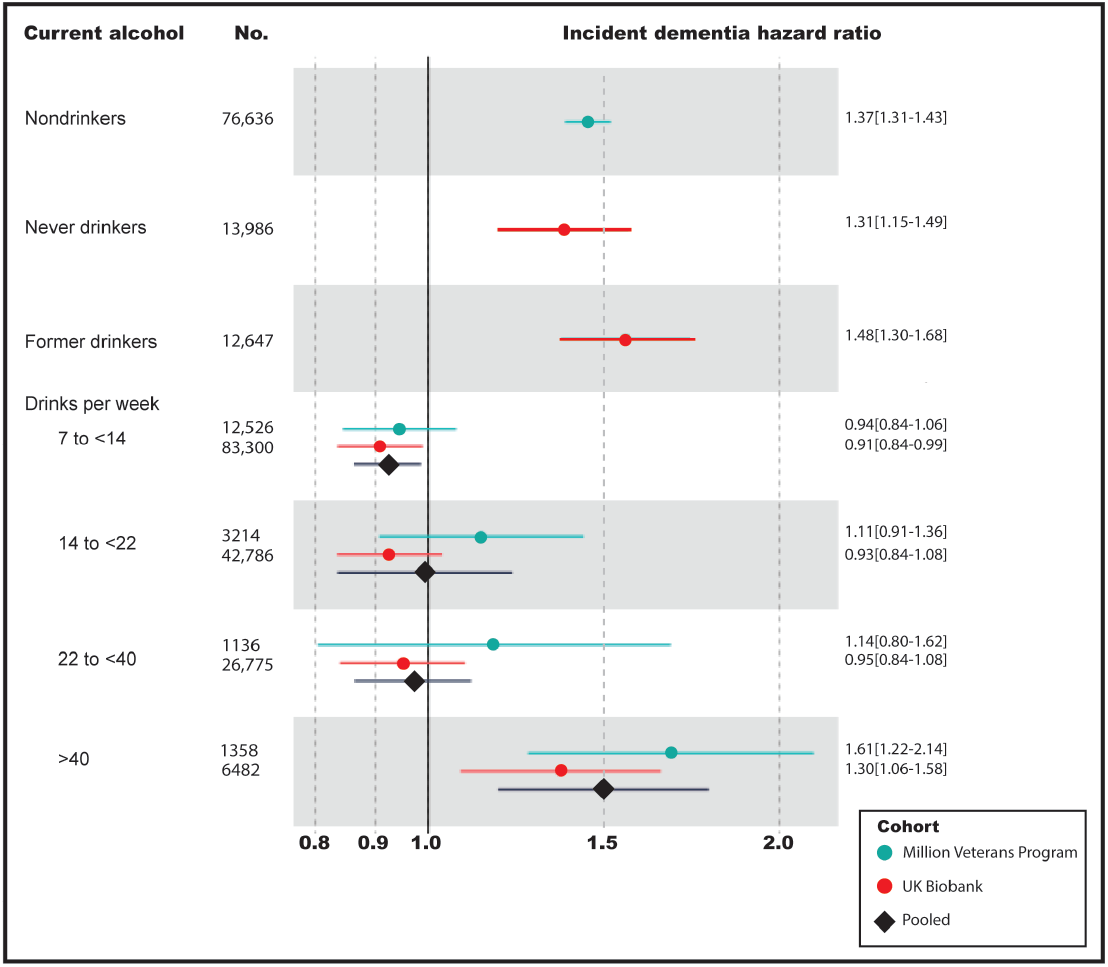
Observational Associations Between Alcohol Intake and Incident Dementia. Hazard ratios (dots or diamonds) and 95% confidence intervals (lines) of all-cause incident dementia according to current alcohol intake, as compared to reference group (solid black line) of individuals consuming <7 drinks per week. Choice of light drinkers as a reference group is motivated by concerns about current-nondrinkers including individuals who previously drank heavily but have reduced their alcohol intake in response to a health concern. Dementia cases were identified by relevant codes in the electronic health record (Table S2). Alcohol intake was ascertained at first recording, from the AUDIT-C screening questionnaire in Million Veteran Program, and the baseline questionnaire in UK Biobank. Individuals classed as current nondrinkers in Million Veteran Program were those with an AUDIT-C score of zero. Never and former drinkers could not be distinguished within this group based on the AUDIT-C questions, whereas in UK Biobank never and former drinkers were identifiable from questionnaire answers. Estimates were generated from Cox proportional hazards models, using unrelated European individuals, and adjusted for age, sex, income, education, smoking, body mass index; and additionally in the Million Veteran Program: head injury, post-traumatic stress disorder and substance use. Estimates for other ancestry groups are given in Table S13. Pooled estimates across the two cohorts (black diamonds) were generated using random effects meta-analysis.

Several sensitivity analyses were conducted to evaluate the competing risk of death (Tables S10&11), the potential mediating role of thiamine (HR after adjustment for dietary thiamine=1.50[1.39-1.61], serum thiamine=1.32[1.15-1.53], Table S12), and cardiovascular risk (HR after adjustment=1.53[1.44-1.63]), and differences between ancestry groups (Table S13). None changed the findings materially. Higher incidence of secondary outcomes, Alzheimer’s and vascular dementia, were also observed in those with an alcohol use disorder and nondrinkers (Table S14).

Frequent binge-drinking (HR=1.31[1.05-1.64]) (Table S15) was identified as conferring an independent risk to DPW on dementia. There was a significant interaction between alcohol use disorder and *ApoE4* (p=0.03), such that the risk of alcohol use disorder on dementia was only evident in *ApoE4* noncarriers (HR=1.40[1.20-1.62]; carriers=1.15[0.84-1.56]). In contrast, sex [AUD*sex HR=1.09[0.79-1.50]], beverage type (Table S9) and *ADH1B* (p<0.7) did not moderate alcohol-dementia associations.

Given previous concerns about reverse causation, we leveraged the longitudinal EHR in MVP to explore how alcohol consumption behaviors changed in the dementia prodrome. Among drinkers, consumption declined over time. However, this decline was faster among those who went on to develop dementia than for controls (dementia*time beta=0.05, se=0.02, p=0.0003, Figure S3). Furthermore, the accelerated drop in drinking was magnified if the original level of drinking was high (first recorded AUDIT-C>4 dementia status*time beta=0.09, se=0.02, p<0.001). Relatedly, associations of alcohol with dementia diagnosis varied with the temporal proximity of the two (Figure S4). When alcohol was measured closer to diagnosis, harmful risks of heavy drinking were attenuated, and moderate drinking was associated with an apparent protective effect (7-14 DPW HR=0.87[0.78-0.97]).

### Genetic associations between alcohol use and dementia

641 independent genetic variants were used to instrument DPW, 80 for problematic alcohol use, and 66 for alcohol use disorder (Supplementary Appendix, Tables S3-6). Higher genetic liability to each alcohol use measure was associated with higher risk of all-cause dementia in the linear analyses in European ancestry (Figure 2). Associations among African ancestry populations were weaker (Table S18) in lower-power analyses. A two-fold increase in alcohol use disorder prevalence was associated with an increase of 16% in dementia risk (IVW OR=1.16[1.03-1.30]).^20^ One standard deviation increase in log-transformed DPW (representing an increase from 1 to 3 DPW, or 5 to 16 DPW) was associated with a 15% increase in dementia risk (IVW OR=1.15[1.03-1.27]).

**Figure 2:**
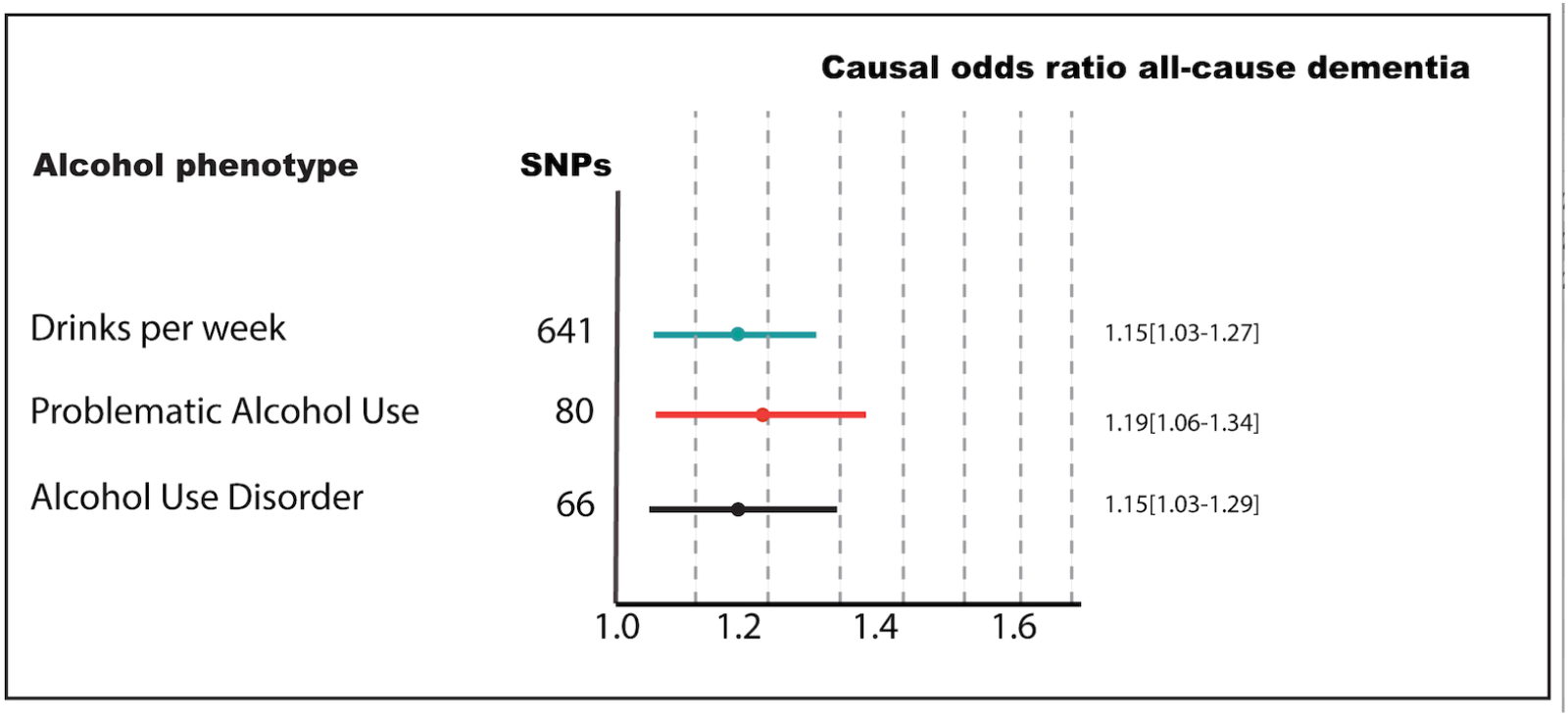
Linear Genetic Associations Between Alcohol Use and Dementia. Forest plot shows causal odds ratios for all-cause dementia for either a 1 standard deviation increase in log-drinks per week or a two-fold increase in prevalence of problematic alcohol use of alcohol use disorder. A stringent (p<5x10^-8^) threshold was used to select genetic instruments from genome-wide association analyses (see Supplementary Appendix for details). Alcohol use is characterized by three phenotypes: a clinical diagnosis of Alcohol Use Disorder in the electronic health record, problematic use (meta-analyzing Alcohol Use Disorder and AUDIT-P, a screening tool for problematic drinking), and number of drinks per week. All-cause dementia was determined by any clinical dementia diagnosis in the electronic health record (Table S2). Estimates were generated from one- and two-sample inverse variance weighted Mendelian randomization from a combined sample size across source genome-wide association studies of over 2.4 million individuals (Table S1).

Sensitivity analyses were used to assess the strength of causal evidence, including correcting for sample overlap bias (Table S19), removal of outliers (Table S18), and examining for reverse direction effects (Table S20), which did not undermine the key findings. *ADH1B*\*rs1229984 was a notable outlier (Figures S8-16). Associations between alcohol use disorder and dementia were attenuated when controlling for income, post-traumatic stress disorder and white matter hyperintensity volume in multivariable Mendelian randomization (Table S22). Genetic predisposition to DPW, unlike problematic or dependent alcohol use, was also associated with proxy dementia and Alzheimer’s disease risk (Figure S17&Table S2) in the best-powered such analysis.

In nonlinear genetic analyses, in contrast to observational analyses, there was no U-shaped association with dementia risk, nor any evidence of any protective effects of low levels of alcohol intake (Figure 3 & S18). Dementia risk increased across the whole range of genetically predicted DPW. For a stratum of the population with a mean consumption of 12 drinks per week, there was evidence for a causal effect on dementia (OR=1.09[1.04 to 1.15)]) (Table S24). Excluding nondrinkers steepened the dose-response relationship, and significant associations could be observed in three out of five strata (Figure S19-21, Table S25). Negative controls for age and sex generally showed no association with genetically-predicted alcohol consumption across strata (Figure S21 & Table S26).

**Figure 3:**
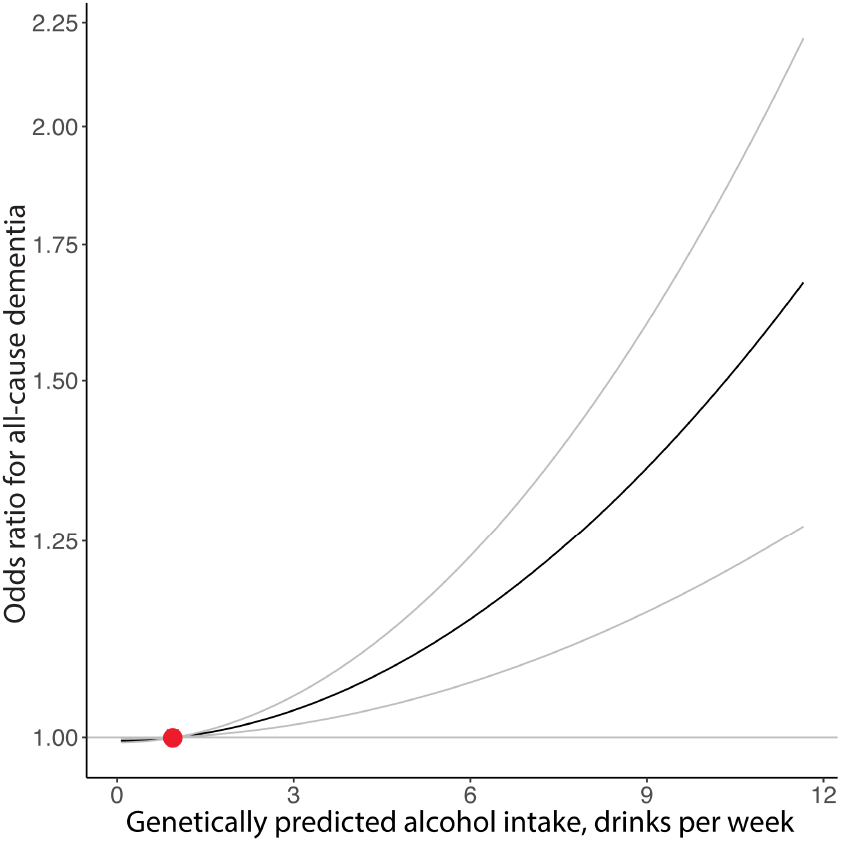
Nonlinear Mendelian Randomization Estimation Of The Dose-Response Curve Between Alcoholic Drinks Per Week And All-Cause Dementia. Nonlinear Mendelian randomization was performed using the doubly-ranked method, in unrelated European ancestry Million Veteran Program participants. N=313,873 (16,932 dementia cases) were included in the analyses. The gradient at each point of the curve is the localized average causal effect. The x-axis shows the alcohol intake in drinks per week at enrollment. Values are based on mean intake in five strata of exposure (quintiles). The y-axis shows the odds ratio for the respective all-cause dementia risk. Grey lines represent the 95% confidence intervals. The reference value (in red) for alcohol intake was taken as 1 drink per week. P-value for linearity: 0.8 (a non-significant result indicates that the best fitting quadratic model is not strongly preferred over the linear model), p-value for trend: 0.5 (a nonsignificant result indicates no strong evidence for trend in the estimates calculated in different strata).

## Discussion

Conventional observational analyses showed a U-shaped association between alcohol and dementia, but genetic analyses provided support for a monotonically increasing causal risk of alcohol on dementia. Drinkers who went on to develop dementia typically reduced their alcohol consumption in the years preceding diagnosis.

In line with other studies,^5^ current nondrinkers had higher incidence of dementia in the observational analyses. Former, and even reportedly ‘never’^21^ drinkers may be ‘sick quitters’ with prior alcohol use disorder, a supposition supported by their on average earlier deaths. Nondrinkers also had lower socioeconomic advantage and education, characteristics highly predictive of lower pre-morbid cognitive performance, and thus increased vulnerability to being diagnosed with dementia. The genetic analyses did not support previously cited protective effects of moderate drinking on dementia. We therefore conclude that associations with alcohol intake ascertained in late-life are likely to have resulted from reverse causation and residual confounding.^5,22^ Our observation that hazardous drinking declines preceding dementia diagnosis supports this supposition. Similar trajectories have been described for other dementia risk factors.^23^ This has important general implications for study design and for the interpretation of prior studies.

Risks of heavy drinking have been described in many,^1,4^ but not all studies.^5^ Heterogeneity in alcohol phenotype, timing of alcohol self-report at mid-versus late-life,^5^ differential adjustment for confounds, and lack of analytic power may explain the discrepant results. With notable exceptions,^5^ the overwhelming majority of studies have been in European ancestry participants. Non-European studies have been sparse and smaller. We observed similar risks of alcohol use across ancestries. Neuroimaging studies have suggested a lower threshold of alcohol consumption for inducing brain changes than we observed for dementia.^3^ Higher sensitivity of imaging over clinical diagnosis may be responsible.

Mendelian randomization estimates offered support for a causal role of alcohol intake on dementia. Nonlinear analyses suggested a monotonically increasing risk of alcohol dose on dementia risk, with no support for a J- or U-shaped curve. Causal inference was strengthened by consistent estimates across multiple alcohol and dementia phenotypes and in multivariable Mendelian randomization but weakened by some stringent methods. Heterogeneity in estimates between genetic variants may plausibly be due to alcohol acting via different pathways and organs to damage the brain, or failure of the homogeneity or linearity assumptions.^24^ We suggest two explanations for discrepancies between our Mendelian randomization results and previous null result studies.^25^ Earlier studies considered a narrow range of alcohol phenotypes and focused on late-onset Alzheimer’s disease, with one exception,^26^ whereas we included a broader range of alcohol and neurodegenerative pathologies. Second, our study had greater statistical power.

Frequent binge-drinking independently conferred a risk of dementia. This is consistent with findings of brain structure,^3^ where frequent binging and drinking to blackout, a binging proxy, steepened the inverse association between alcohol dose and grey matter volume.^27^ Conversely, alcoholic beverage type appeared immaterial, not substantiating a protective effect of wine constituents.^28^

We probed several possible mechanisms by which alcohol could lead to dementia. We found no support for a neurotoxic role of acetaldehyde, a key breakdown product of ethanol, using as a proxy the nonsignificant alcohol\**ADH1B* interaction.^29^ However in Mendelian randomization analyses *ADH1B* was an outlier. There may be a competing effect of cancer diagnosis (the protective allele increases cancer risk),^17^ or *ADH1B* may have a biologically distinct action, through acetaldehyde for instance. Thiamine deficiency in alcohol abuse is a recognized pathway to brain damage including Korsakoff’s syndrome.^14^ However, we found no support for a mediating role of thiamine, cardiometabolic disease^30^ nor of cholesterol dysregulation. We hypothesize that survival bias underlies the weaker alcohol use disorder-dementia association in *ApoE4* carriers, of whom there were relatively few in the sample. Ethanol may be directly neurotoxic.^14^ Since alcohol dehydrogenase is present at the blood brain barrier, high systemic ethanol levels would be necessary for ethanol brain entry. Our observed harmful impact of binge-drinking supports this hypothesis.

This study has several strengths, including a large sample size, cross-ancestry analyses, triangulation of observational and genetically informed approaches, examination of drinking dose and pattern, and of postulated confound and mediators.

First among its limitations, analyses had greatest power to detect effects in European ancestry groups. Second, EHR dementia diagnoses may be subject to ascertainment bias. However, this would likely be skewed to more severe cases, biasing associations towards the null. In view of the age distribution of the cohorts, many early onset dementia cases would not be captured.^11^ Third, different mean age of MVP and UKB participants should be considered when interpretating the pooled estimates from the meta-analysis. Fourth, blood thiamine measurements were only available on a small nonrandom subset, and dietary thiamine was captured later. Fifth, Mendelian randomization relies on unverifiable assumptions, and estimates the effect of lifelong exposure. Such estimates do not necessarily translate into potential consequences resulting from an adult life intervention. We observed alcohol intake changing over time in MVP, hence MR estimates should be interpreted as the accumulated effect of alcohol over time rather than point estimates. Sixth, estimates from one-sample Mendelian randomization may be biased towards observational estimates,^20^ and for binary exposures should be interpreted as for genetic compliers, likely a population subgroup. Finally, nonlinear Mendelian randomization estimates at the lower end of alcohol distribution have less precision. However, negative controls for age and sex did not indicate bias across strata.^19^

To summarize, observational methods initially suggested a U-shaped association between alcohol and dementia, aligning with existing literature. However, genetic analyses revealed a more nuanced picture, indicating a consistent, dose-responsive relationship between alcohol and dementia risk. Intriguingly, we observed a trend of reduced alcohol use prior to dementia diagnosis, a pattern that might reflect either a subclinical manifestation of disease or behavioral adaptation to early cognitive changes. Divergence between observational and genetic data underscores the complexity of delineating causal pathways in alcohol-related neurodegeneration and highlights the potential for confounding factors, such as reverse causation, in conventional observational studies. Our findings suggest a causal detrimental impact of alcohol on dementia, challenging previous perceptions of a potentially protective effect at lower levels of consumption. Reducing alcohol may be an effective dementia prevention strategy.

## Data Availability

Access to UK Biobank data is available following a successful application. Access to Million Veteran Program summary statistics is available through via dbGAP.

## Contributions

AT conceived the study design, performed statistical analyses and wrote the first draft of the manuscript. DL accessed the data and performed analyses. KA performed analysis. HZ and JD advised on statistical analyses. MS, MG, KPE and TN advised on study design. JG supervised the project. All authors helped to write the paper and had final responsibility for the decision to submit for publication. AT and DL accessed and verified the data. We wish to thank Maria Christodoulou and the University of Oxford Statistics Consulting service for providing advice on the survival model analyses.

## Acknowledgments

This research used data from the Million Veteran Program and was supported by funding from the Department of Veterans Affairs Office of Research and Development, Million Veteran Program Grant #I01CX001849, and the VA Cooperative Studies Program (CSP) study #575B, and MVP025. This publication does not represent the views of the Department of Veterans Affairs or the United States Government. AT is supported by the Wellcome Trust (216462/Z/19/Z). SBu is supported by the Wellcome Trust (225790/Z/22/Z) and the United Kingdom Research and Innovation Medical Research Council (MC_UU_00002/7). This research was supported by the National Institute for Health Research Cambridge Biomedical Research Centre (NIHR203312).

