## Supplementary material for "Alcohol use and risk of dementia in diverse populations": Online supplement

Supplementary Appendix

[Table S3: Alcohol use disorder instruments p<5x10^-8^.	31](#_Toc160195683)

[Table S4: Alcohol use disorder instruments, p<5x10^-5^, African ancestry.	34](#_Toc160195684)

[Table S5: Problematic alcohol use instruments, p<5x10-8, European ancestry.	40](#_Toc160195685)

[Table S6: Drinks per week instruments, p<5x10^-8^, European ancestry.	43](#_Toc160195686)

### Supplementary methods

#### Alcohol measurement

Drinks per week were calculated in Million Veteran Program (MVP) as the product of the midpoints of frequency of drinking and number of drinks consumed per day. One drink equates to ~14 grams ethanol. The AUDIT is a 10-item questionnaire developed to assess quickly for hazardous patterns of alcohol consumption and related alcohol problems. The AUDIT questionnaire can be split into multiple sub scores, including the first 3 items assessing patterns of alcohol consumption (the AUDIT-C), the final 7 items assessing alcohol-related problems (the AUDIT-P), and an overall total score aggregated across all 10 items (the AUDIT-T). AUDIT-C was used clinically to identify high-risk drinkers recorded at multiple time points in the electronic health record preceding enrolment. The AUDIT-C summed score (range 0-12) collates answers about the frequency of drinking (never to 4+ times a week), the number of standard drinks consumed on a typical drinking day (1-10 drinks), and the frequency of consuming more than 6 drinks in one episode (‘binging’). AUDIT-C score was additionally categorized according to previously suggested cut-offs: non or occasional drinker (0,1), low risk (2-3), high risk (>4).^1,2^ For sensitivity analyses, drinkers were grouped as primarily wine, beer or liquor drinkers if they reported weekly intake of one beverage, and irregular (<weekly) intake of others on the lifestyle survey.

#### Covariate measurement

Potential confounds and effect modifiers were identified based on published literature.^3-5^

**MVP:** Education was treated as a categorical variable as self-reported: less than high school, high school diploma, some college credit, associates degree, Bachelor’s degree, Master’s degree, Professional or Doctoral degree. Income of participant’s household was recorded as a categorical variable: less than $10,000, $10,000-19,999, $20,000-29,999, $30,000 to $39,999, $40,000 to $49,999, $50,000 to $59,999, $60,000 to $149,000, or $150,000 or more. Smoking was recorded as daily, occasionally, or not at all. Body mass index was calculated from self-reported height and weight at enrollment. History of head injury and post-traumatic stress disorder were self-reported and coded as binary variables. Substance use disorders were defined as a lifetime history of opioid or cannabis dependence ICD code in the electronic health record. Diabetes mellitus was recorded at enrollment survey. Mean systolic and diastolic blood pressure was calculated across multiple measurements recorded in the electronic health record. Two measures of thiamine were used:^6^ 1) dietary intake (mg/day with and without supplementation) at enrollment, 2) serum/whole blood recorded in electronic health record. To ensure comparability between serum and whole blood thiamine measurements (differing sensitivities), measures were standardized using z-scores.

**UKB:** Educational qualifications, from high to low, were reported as: college or university degree, A levels or equivalent, O levels or equivalent, CSEs (Certificate of Secondary Education) or equivalent, NVQ (National Vocational Qualification) or equivalent, other professional qualifications, or none (lowest level used as reference). Smoking status was reported in categories: never/previous/current. Systolic (SBP) and diastolic (DBP) blood pressure were automated measurements at baseline. Body mass index (BMI) was calculated from measured height and weight. Diabetes mellitus diagnoses were generated by a UKB algorithm using self-report, hospital care records, and death certificates. Subtypes of diabetes mellitus (insulin-dependent, noninsulin-dependent, unspecified were combined to generate a binary diabetes mellitus (present/absent) variable.

#### Genotyping

Genotyping and imputation of Million Veteran Program participants has been described previously.^7^ We used genetic data release 4. Briefly, a customized Affymetrix Axiom Array was used for genotyping. Genotype data for biallelic single nucleotide polymorphisms were imputed using Minimac4 and a reference panel from the African Genome Resources panel by the Sanger Institute. Indels and complex variants were imputed independently using the 1000 Genomes (1KG) phase 3 panel and merged in an approach similar to that employed by the UK Biobank. Designation of broad ancestries was based on genetic assignment with comparison to 1KG reference panels.^8^

#### Statistical analyses

A study protocol was outlined in a fellowship proposal but not pre-registered.

##### Cox proportional hazards regression:

Time at risk was calculated from baseline (when covariates were measured) to diagnosis of censoring. Influential observations were assessed by plotting deviance residuals. Proportional hazards were assessed visually using Schoenfeld residuals and formally with time interactions. For covariates not of primary interest (age and body mass index) which violated the proportional hazards assumption, stratified models were fitted without the constraint of non-proportionality. Separate baseline hazard functions were fitted for each stratum. The Aalen-Johansen estimator^9^ was used to assess if death was a competing risk. Competing risk of death was accounted for using subdistribution method.^10^

##### Restricted cubic splines:

Five knots were applied to AUDIT-C sum scores.[31] Non-linearity was formally tested (H0: β2=β3=...=βk−1= 0) with an F-test.

##### Longitudinal modelling of AUDIT-C:

To assess how drinking behaviours changed over time, binomial regression was used. Models included the following fixed effects: time (diagnosis/study-end – date of alcohol measurement), dementia (case/control), enrollment age, sex, education and income, and a three-way interaction term between time*dementia status*AUDIT-C category (non-(0/1)/low-(<4)/high-risk (>4) drinker). We used Wald tests, estimating the overall effect of interactions between dementia status and time on the models, to test the null hypothesis that AUDIT-C trajectory did not differ according to dementia diagnosis. Participant identification was included as a random effect. Resultant models were visually presented using graphs showing predicted longitudinal trends in AUDIT-C scores for a typical participant.

##### Linkage disequilibrium clumping:

Ancestry-specific linkage disequilibrium clumping was performed using PLINK v2.0 with the respective 1000 Genomes Project phase 3 linkage disequilibrium reference panels. Lead variants were identified within 10,000 kb and LD r^2^=0.001.

##### Genome-wide association studies:

Related (kinship coefficient >0.088) participants were excluded. Logistic regression was performed in PLINK 2.0 using the first 10 principal components, sex, and age as covariates. The Euclidean distances between each Million Veteran Program participant and the centres of the five reference ancestral groups from 1000 Genomes Project were calculated using the first 10 principal components, with each participant assigned to the nearest reference ancestry. A second round of principal component analysis within each assigned ancestral group was performed and outliers with principal component scores >6 standard deviations from the mean of any of the 10 principal components were removed. Variants were excluded if call missingness in the best-guess genotype exceeded 20%. Alleles with minor allele frequency <0.1% were excluded in European, and African defined ancestries. For the genome-wide association study of European ancestry, after excluding those with missing data, there were 25,473 cases and 425,844 controls, and for African ancestry there were 5,706 cases and 108,532 controls. Insufficient power precluded analysis of the Admixed American ancestry group.

##### Linkage disequilibrium score regression:

Single nucleotide-based heritability was calculated for common variants mapped to HapMap3. Linkage disequilibrium score regression was also used to estimate the genetic correlation between alcohol and dementia phenotypes (excluding clinical Alzheimer’s disease due to unavailability of full summary statistics).

##### Selection of genetic variants

Instruments were selected on the basis of the largest available genome-wide association studies. For alcoholic drinks per week, instruments were selected at genome-wide significance in the trans-ancestry analyses, but then ancestry-specific betas and standard errors were used. As a result, some instruments had higher p values and lower F statistics. Post hoc choice of instruments, genetic models or data based on measured F-statistics can exacerbate bias. In particular, the commonly cited rule of thumb that F > 10 avoids bias in IV analysis is misleading.^11^ Multi-allelic instruments were included given that all datasets clearly report multiple alleles allowing comparison. The same covariate set (age, sex, ancestry principal components) was used for adjustment in all comprising genome-wide association studies.

##### Linear Mendelian randomization:

MR rests on three assumptions: relevance, independence and exclusion restriction. Where alcohol-associated genetic variants were not available in the outcome summary statistics, proxy variants (linkage disequilibrium r^2^>0.8) were searched for using LDproxy (<https://ldlink.nih.gov/?tab=ldproxy>). 61(92%) of AUD^12^-associated genetic variants, 78(96%) of problematic alcohol use^12^-associated and 499(59%) of drinks per week^13^-associated genetic variants were present in the Million Veteran Program dementia dataset. Proxy variants were available for 5 alcohol use disorder, 5 problematic alcohol use and 142 drinks per week variants (Tables S3-6). All 4 alcohol use disorder-associated variants in African ancestry were available, and 144 (69%) of variants (+1 proxy) identified with a less stringent threshold (p<5x10^-5^). Analyses were conducted using R packages *MendelianRandomization* (version 0.5.1), *TwoSampleMR* (version 0.5.6) and *MRlap* (version 0.0.3). Inverse variance weighted analysis (multiplicative random effects) regresses the effect sizes of the variant-dementia associations against the effect sizes of the variant-alcohol associations. Dementia estimates for drinks per week were converted for interpretability from log-transformed drinks per week based on calculations in UK Biobank of drinkers, from standard errors of log-transformed weekly intake. The MR-Egger method uses a weighted regression with an unconstrained intercept to relax the assumption that all genetic variants are valid instrumental variables (under the Instrument Strength Independent of Direct Effect (InSIDE) assumption)^14^. A non-zero intercept term can be interpreted as evidence of directional pleiotropy, where an instrument is independently associated with the outcome violating an MR assumption. The median and modal Mendelian randomization methods are also more resistant to pleiotropy, as they are robust when up to 50% of genetic variants or more than not, respectively, are invalid. MRlap corrects for bias in inverse variance weighted estimates due to overlapping samples^15^. MR-PRESSO tests for and corrects horizontal pleiotropy^16^. These methods are recommended in practice for sensitivity analyses as they require different assumptions to be satisfied, and therefore if estimates from such methods are similar, then any causal claim inferred is more credible. Heterogeneity of inverse variance weighted estimates was assessed using Cochran’s Q statistic. Multivariable Mendelian randomization was used to test whether the causal effect of alcohol on dementia was altered by adjustment for key potential confounds or mediators: smoking, income, post-traumatic stress disorder, cannabis dependence or measures of brain structure linked to dementia (volumes of global brain, hippocampus and white matter hyperintensities).^17^ Based on a R^2^ of 0.088 and a significance level of 0.05, the sample size of n~400,000 (including 30,000 cases) has 80% power to detect a causal odds ratio of 1.20 per standard deviation change in alcohol consumption.^18^

##### Nonlinear Mendelian randomization:

A weighted genetic risk score for alcoholic drinks per week was calculated for each individual by multiplying the number of alcohol-increasing alleles the individual carries by the effect size of the allele with alcohol, and summing across the 641 SNPs. To estimate the nonlinear relationship between genetically-predicted alcohol and dementia, a fractional polynomial method was applied. First, the sample was divided into five strata using the doubly-ranked method.^19^ Any choice of stratum number is arbitrary. Here we selected 5 as a balance between allowing assessment of the shape of the relationship and the decreasing power of analyses as group size reduces. The linear MR estimates (localized average causal effects, LACE), in each stratum were then calculated as a ratio of coefficients: the association of the genetic score with the dementia divided by the association of the genetic score with alcohol. Drinks per week measured as a continuous variable at enrolment were used, as earlier measures were in DPW categories only. The former was calculated using logistic regression, the latter with linear regression. All associations were adjusted for age, age^2^, sex and top 10 principal ancestry components. We performed meta-regression of the LACE estimates against the mean of alcohol in each stratum in a flexible semiparametric framework using the derivative of the fractional polynomial model of degrees 1 and 2. The reference point in analyses was set (arbitrarily) to 1 drink per week. We report two tests for nonlinearity: a linearity test (assessing whether a nonlinear model fits better than linear), and a trend test, which tests for a linear trend amongst LACE estimates. All statistical analyses were performed using the SUMnlmr package. In light of recent suggestions, we performed negative control analyses of age and sex.^20^

### Supplementary figures

#### Figure S1: Analyses overview

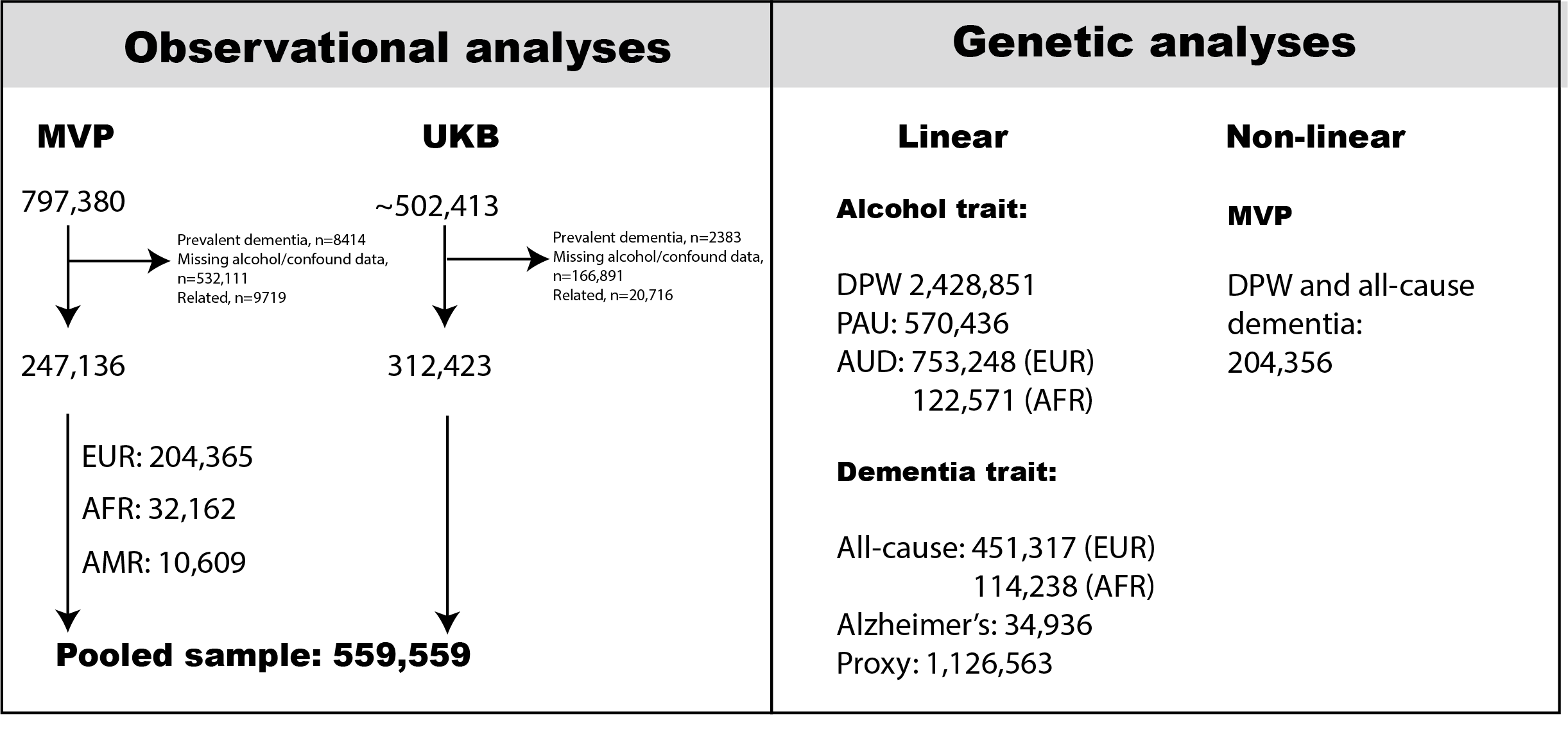

Abbreviations: MVP – Million Veteran Program, UKB – UK Biobank, EUR - European ancestry, AFR – African American ancestry, AMR - admixed American ancestry, DPW – drinks per week, PAU – problematic alcohol use, AUD – alcohol use disorder.

#### Figure S2: First recorded AUDIT-C score (sum) and hazard of incident all-cause dementia.

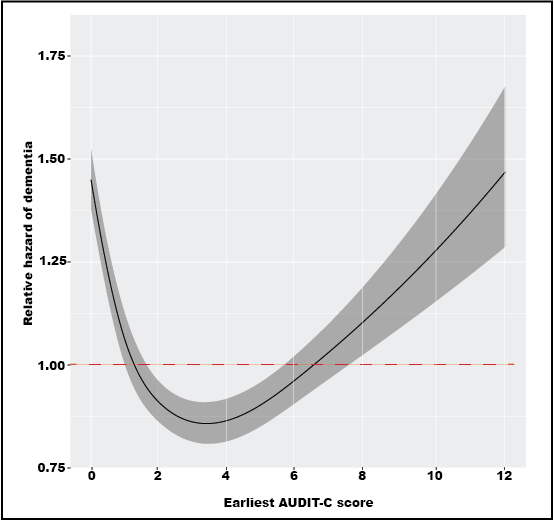

AUDIT-C measured mean 9 years prior to dementia or end of follow-up in unrelated participants of Europeans ancestry (total n=176,868, n=7667 cases). Estimates were generated using Cox proportional hazards models adjusted for: age, age^2^, age^3^, sex, age*sex, education, income, smoking, body mass index, time AUDIT before enrollment. Restricted cubic splines (5 knots) were applied AUDIT-C scores. Plots depict associations for an average participant (male of mean age and body mass index, non-smoker and median education and income level for sample). Nonlinear chi-square=110.79, p<0.0001.

#### Figure S3: Longitudinal Trends in Alcohol Use Preceding Dementia Diagnosis.

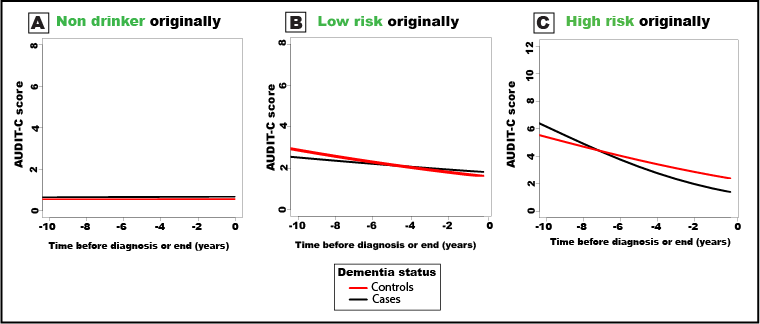

Alcohol use is defined using the AUDIT-C screening questionnaire which was administered and documented at multiple time points in the electronic health record. Dementia status was ascertained by the presence of a relevant clinical code in the electronic health record. The plots show predictions of how drinking behavior changes over time for individuals who develop dementia (cases) and those who remain dementia-free (controls) for: A, individuals who were non- or occasional drinkers at first record (AUDIT-C<1, No.=108,544); B, individuals who were low-risk drinkers at first record (AUDIT-C 2- <4, No.=51,443); C, individuals who were high-risk drinkers at first record (AUDIT-C >4, No.=16,881). Time 0 is the time of diagnosis for cases or last follow up for controls. Predictions are based on mixed effects models adjusted for age, sex, body mass index, smoking, educational qualification and household income. Graphs show predictions for a male participant of average age, body mass index, education and income.

#### Figure S4: Association of alcohol intake and dementia, according to timing of alcohol self-report, in the Million Veteran Program.

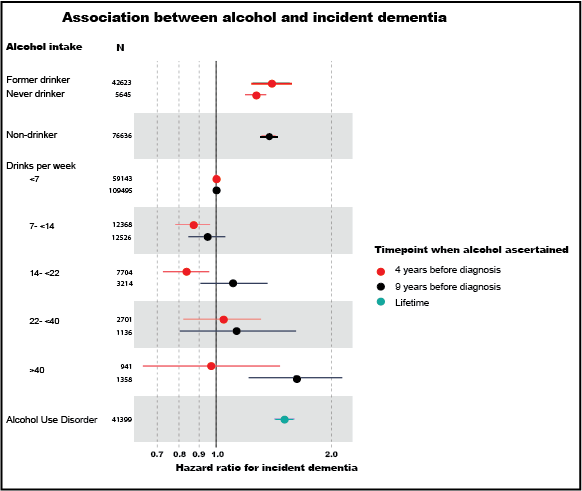

Estimates were generated from Cox proportional hazards models in European ancestry individuals from Million Veteran Program, adjusted for: age, sex, income, education, smoking, body mass index, head injury, post-traumatic stress disorder, substance use. The reference group was <7 drinks per week or controls. Alcohol was ascertained on average 9 years before diagnosis from the electronic health record AUDIT-C score, and on average 4 years before diagnosis at baseline surveys. Non-drinkers 9 years before diagnosis were those with AUDIT-C=0 and could not be separated into never and former drinkers.

#### Figure S5: Manhattan plot showing genome-wide associations with all-cause dementia in Million Veteran Program participants of European ancestry.

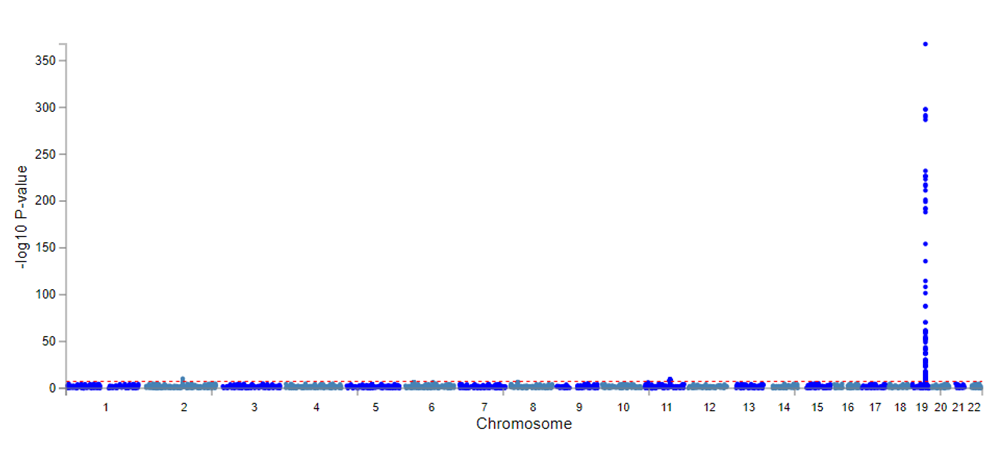

All-cause dementia was determined by the presence of a relevant ICD dementia code in the linked electronic health record (Table S2). Analysis was conducted in 25,473 all-cause dementia cases and 425,844 controls of European ancestry who were unrelated.

#### Figure S6: Manhattan plot showing genome-wide associations with all-cause dementia in Million Veteran Program participants of African American ancestry.

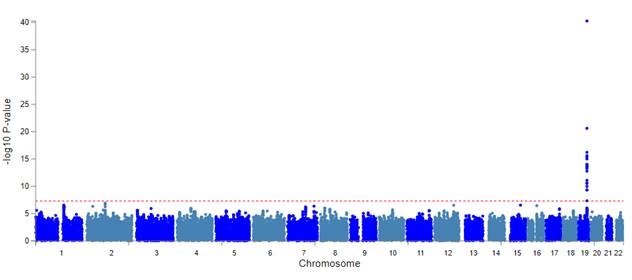

All-cause dementia was determined by the presence of a relevant ICD dementia code in the linked electronic health record (Table S1). Analysis was conducted in 5,706 all-cause dementia cases and 108,532 controls of African ancestry who were unrelated.

#### Figure S7: Genetic Heritability and Correlation of Alcohol Use and Dementia.

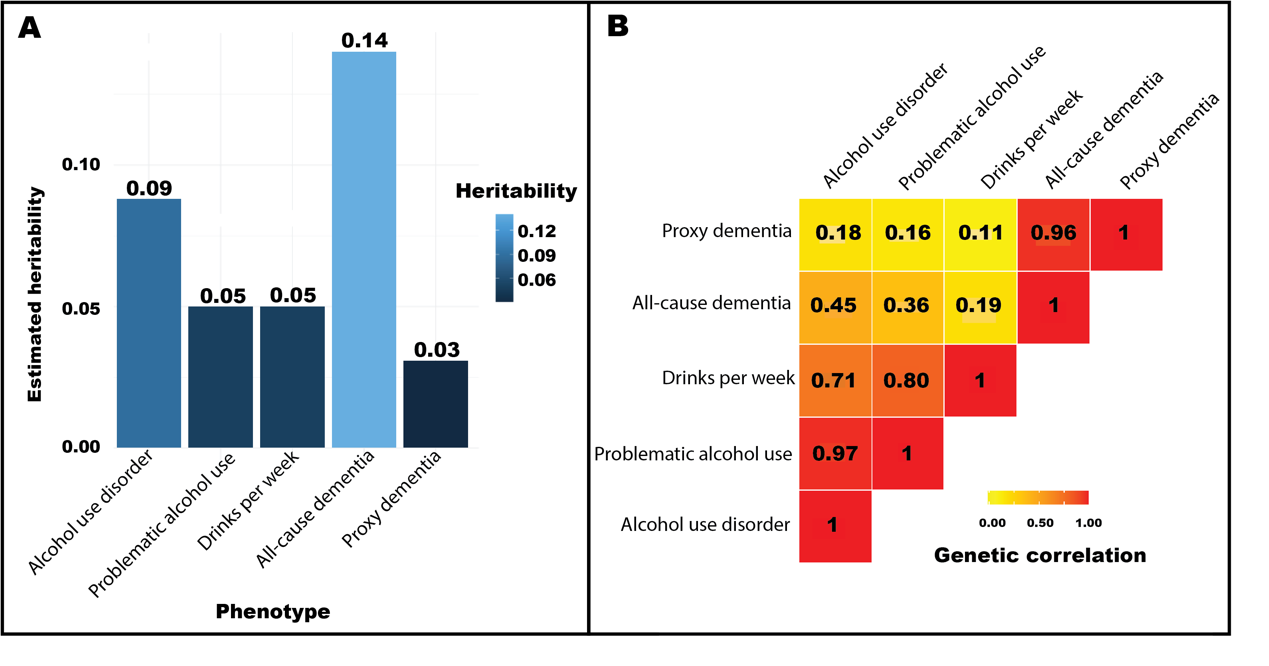

**A,** Bar chart shows the genetic heritability of alcohol and dementia phenotypes. Heritability was estimated using linkage disequilibrium score regression using single nucleotide polymorphisms and shown on the observed scale for alcohol phenotypes and the liability scale for dementia phenotypes. **B,** Heat map shows genetic correlations between alcohol and dementia phenotypes, as estimated by linkage disequilibrium score regression. Alcohol use is characterized by three phenotypes: a clinical diagnosis of Alcohol Use Disorder in the electronic health record, problematic use (meta-analyzing Alcohol Use Disorder and AUDIT-P, a screening tool for problematic drinking), and number of drinks per week. All-cause dementia was determined by any clinical dementia diagnosis in the electronic health record (Table S2). Proxy dementia is defined as either a diagnosis of Alzheimer’s disease or a parental history of dementia. Genetic associations with all-cause dementia were calculated in this study in Million Veteran Program. Those for alcohol phenotypes and proxy dementia were previously published. All results refer to individuals of European ancestry.

#### Figure S8: Scatter plot of Mendelian randomization analysis of alcohol use disorder and all-cause dementia.

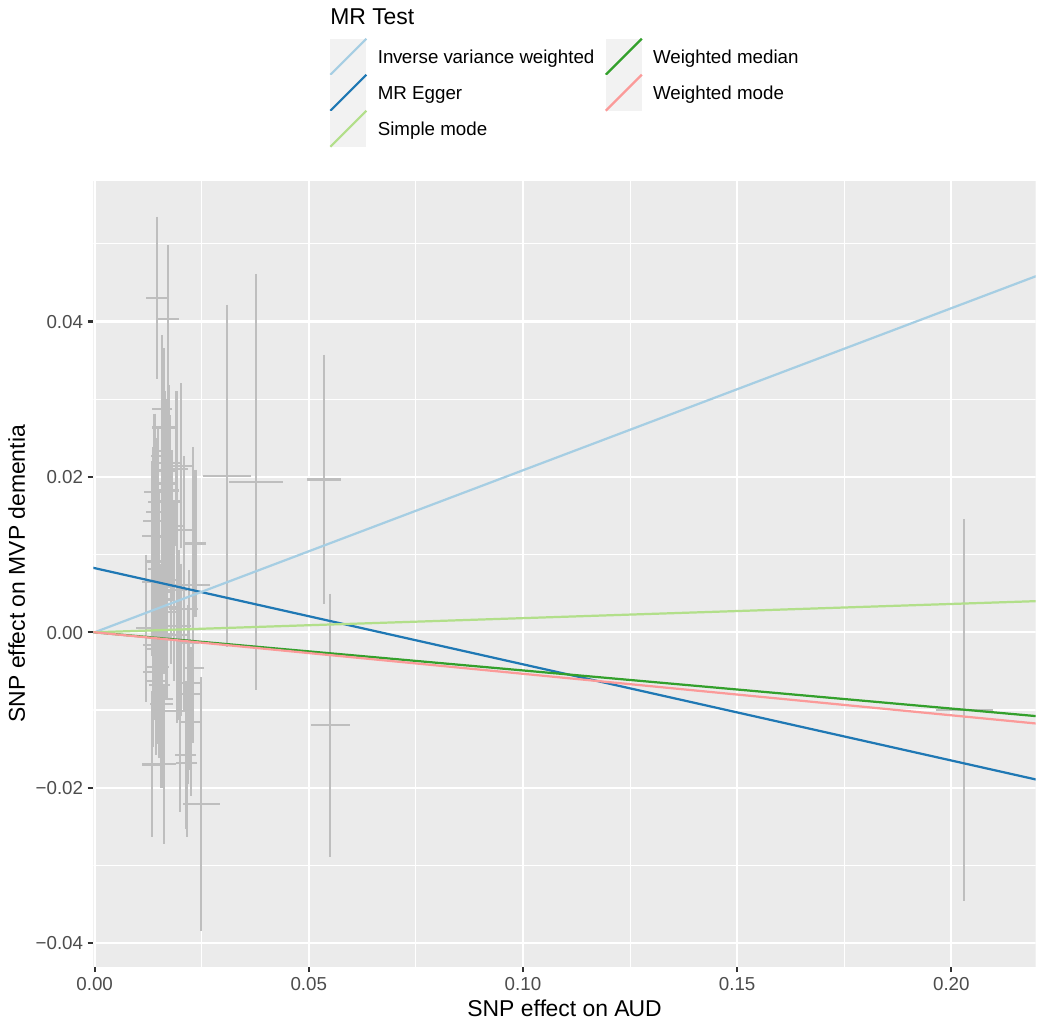

All-cause dementia and alcohol use disorder (AUD) were defined by the presence of a relevant ICD code in the electronic health record (Table S2). Genetic associations with dementia were calculated *de novo* in this study in Million Veteran Program, and with alcohol use disorder from Zhou et al.^12^

#### Figure S9: Scatter plot of Mendelian randomization analysis of problematic alcohol use and all-cause dementia.

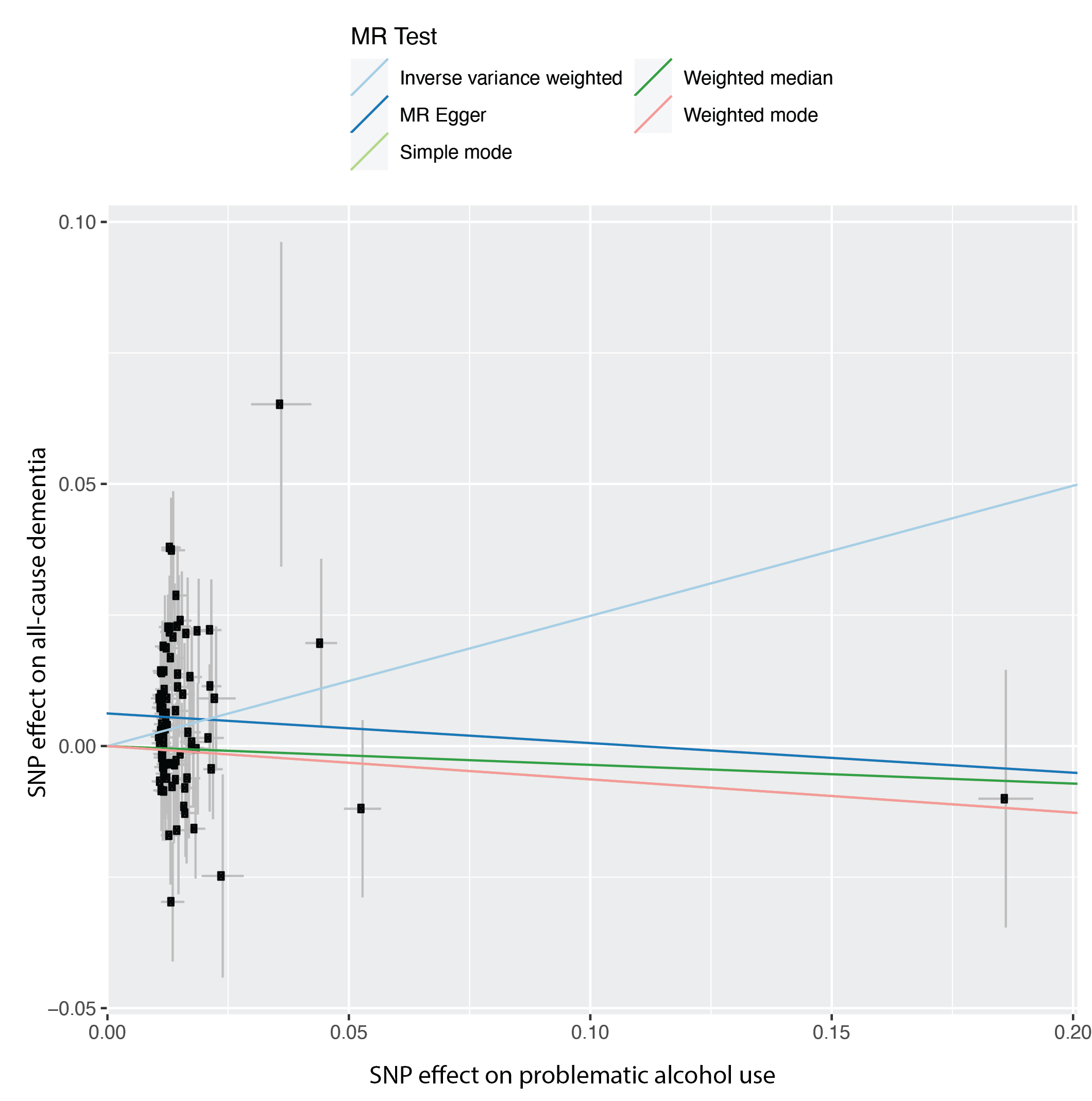

All-cause was defined by the presence of a relevant ICD code in the electronic health record (Table S2). Problematic alcohol use was defined by meta-analyzing alcohol use disorder and AUDIT-P. Genetic associations with dementia were calculated *de novo* in this study in Million Veteran Program, and with problematic alcohol use from Zhou et al.^12^

#### Figure S10: Scatter plot of Mendelian randomization analysis of drinks per week and all-cause dementia.

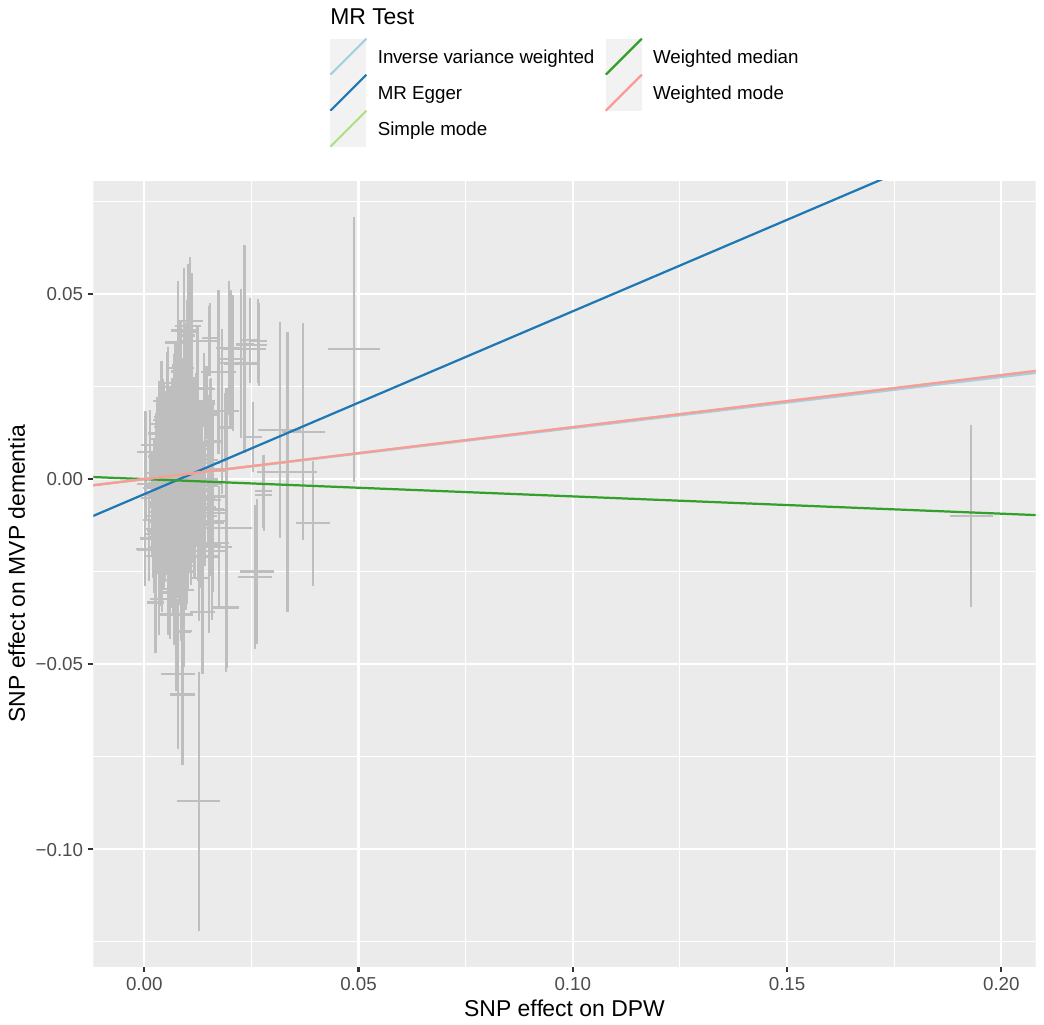

All-cause was defined by the presence of a relevant ICD code in the electronic health record (Table S2). Genetic associations with dementia were calculated de novo in this study in Million Veteran Program, and with drinks per week from Saunders et al^.13^

#### Figure S11: Scatter plot of Mendelian randomization analysis of alcohol use disorder and proxy dementia.

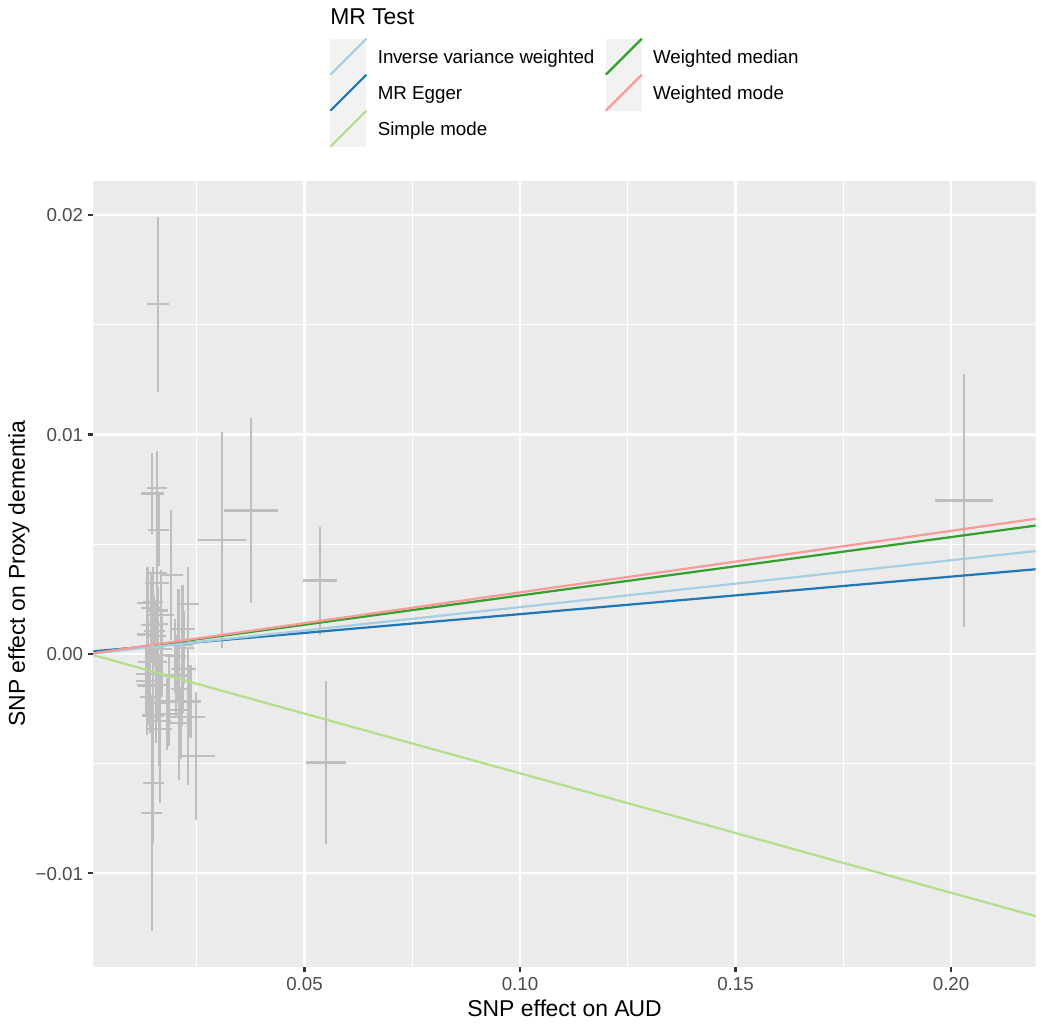

Alcohol use disorder (AUD) was defined by the presence of a relevant ICD code in the electronic health record. Proxy dementia was defined as a history of Alzheimer’s disease or a parental history of dementia. Genetic associations with alcohol use disorder originated from Zhou et al.,^12^ and with proxy dementia from Wightman et al.^21^

#### Figure S12: Scatter plot of Mendelian randomization analysis of problematic alcohol use and proxy dementia.

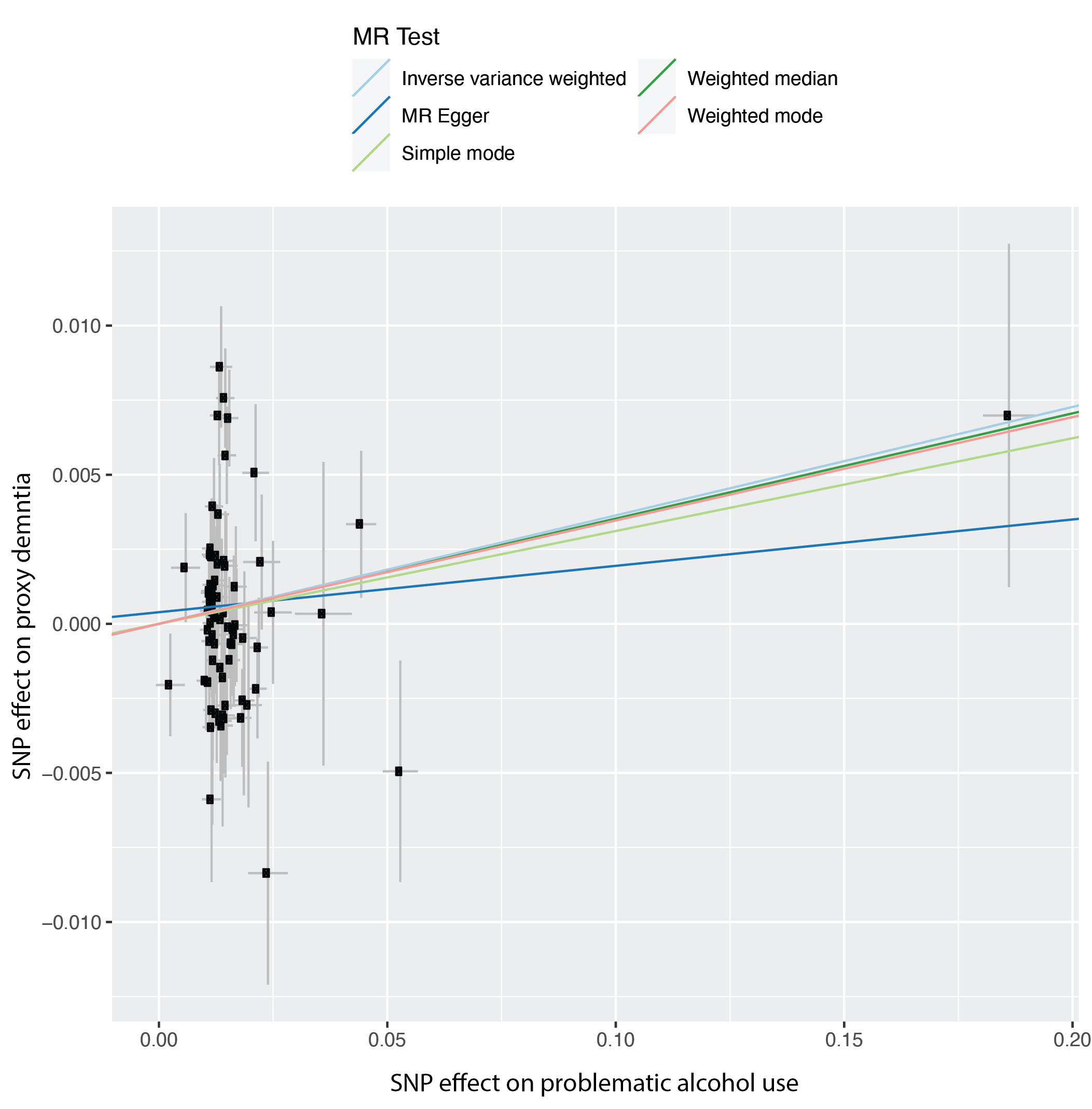

Problematic alcohol use was defined by meta-analyzing alcohol use disorder and AUDIT-P. Proxy dementia was defined as a history of Alzheimer’s disease or a parental history of dementia. Genetic associations with problematic alcohol use were sought from Zhou et al.,^12^ and with proxy dementia from Wightman et al.^21^

#### Figure S13: Scatter plot of Mendelian randomization analysis of drinks per week proxy dementia.

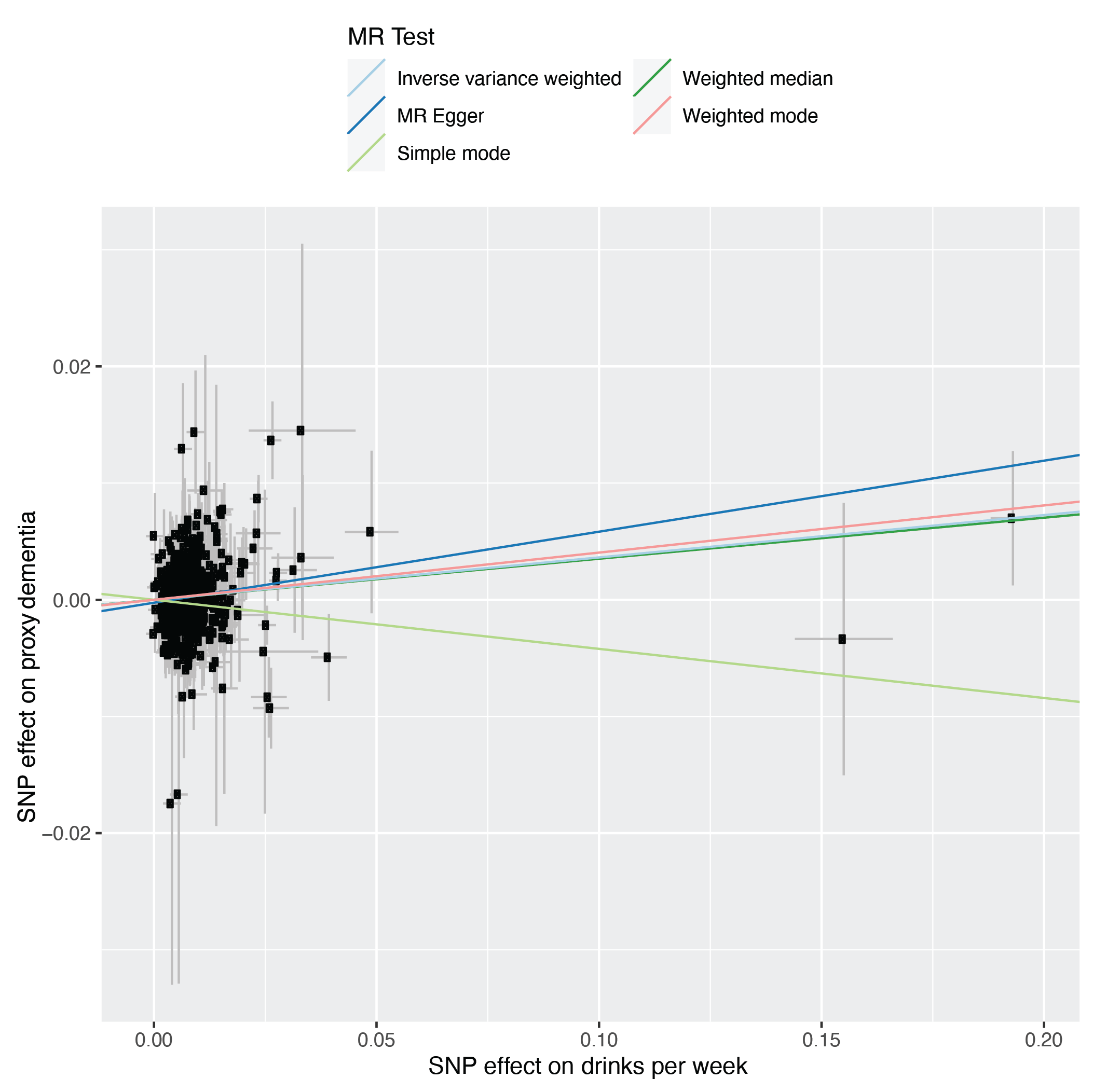

Proxy dementia was defined as a history of Alzheimer’s disease or a parental history of dementia. Genetic associations with drinks per week were sought from Saunders et al.,^13^ and with proxy dementia from Wightman et al.^21^

#### Figure S14: Scatterplot of Mendelian randomization analyses of alcohol use disorder and clinically diagnosed Alzheimer’s disease.

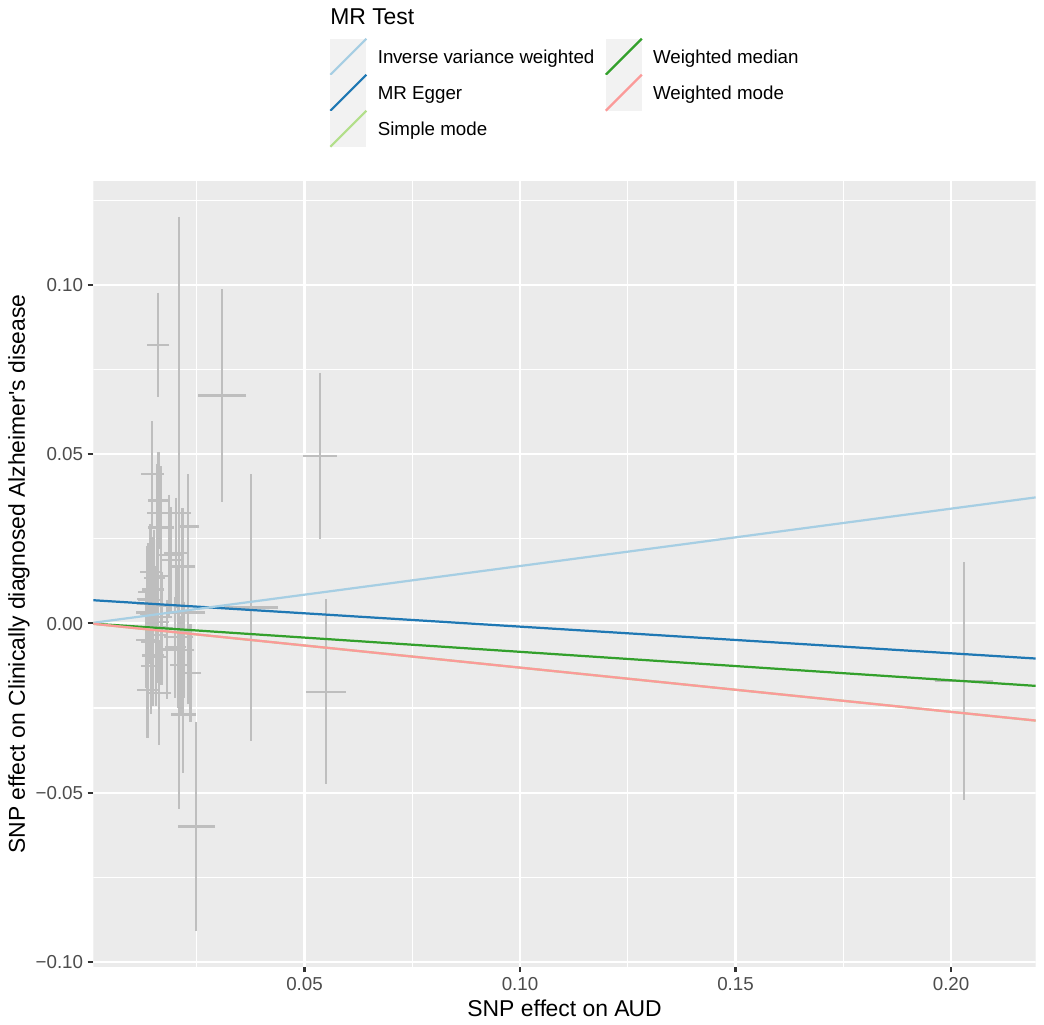

Alcohol use disorder (AUD) was defined by the presence of a relevant ICD code in the electronic health record. Alzheimer’s disease was clinically diagnosed. Genetic associations with alcohol use disorder originated from Zhou et al.,^12^ and with Alzheimer’s disease from Kunkle et al.^22^

#### Figure S15: Scatterplot of Mendelian randomization analyses of problematic alcohol use and clinically diagnosed Alzheimer’s disease.

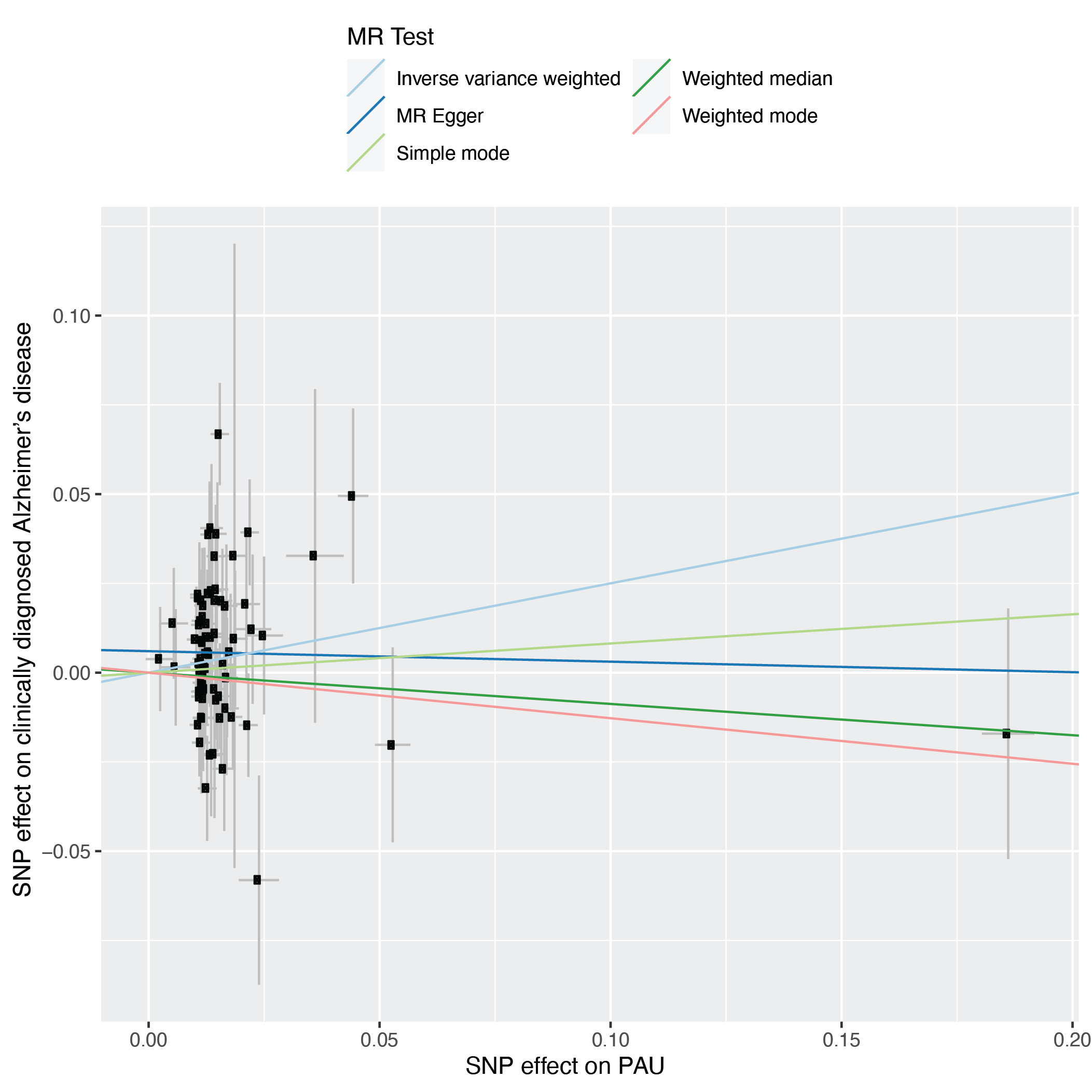

Genetic associations with problematic alcohol use (meta-analyzed alcohol use disorder and AUDIT-P) were derived from Zhou et al.,^12^ and those with clinically diagnosed Alzheimer’s disease from Kunkle et al.^22^

#### Figure S16: Scatterplot of Mendelian randomization analyses of drinks per week and clinically diagnosed Alzheimer’s disease.

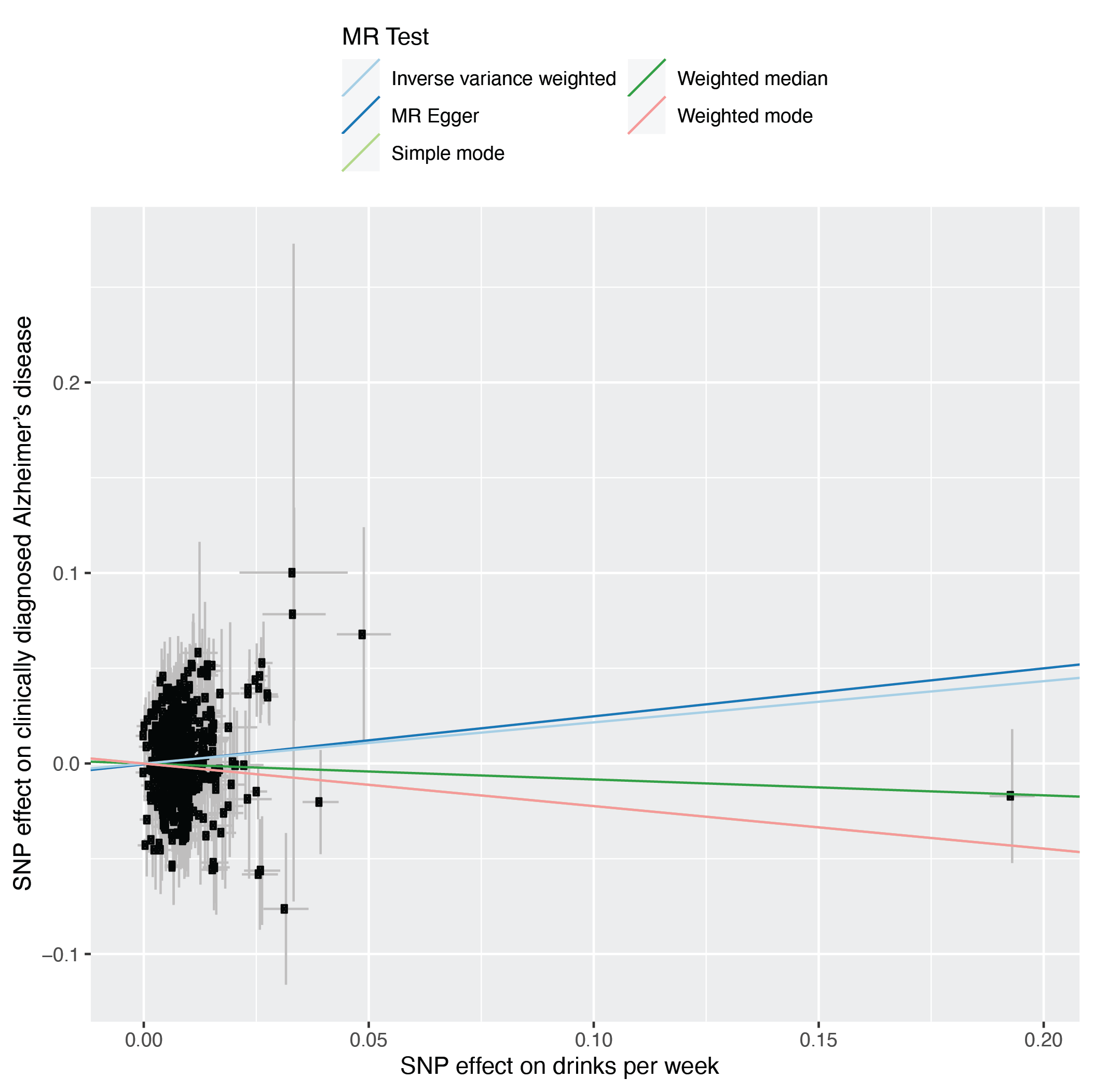

Genetic associations with drinks per week were derived from Saunders et al.,^13^ and those with clinically diagnosed Alzheimer’s disease from Kunkle et al.^22^

#### Figure S17: Genetic associations between alcohol phenotypes and differing dementia phenotypes.

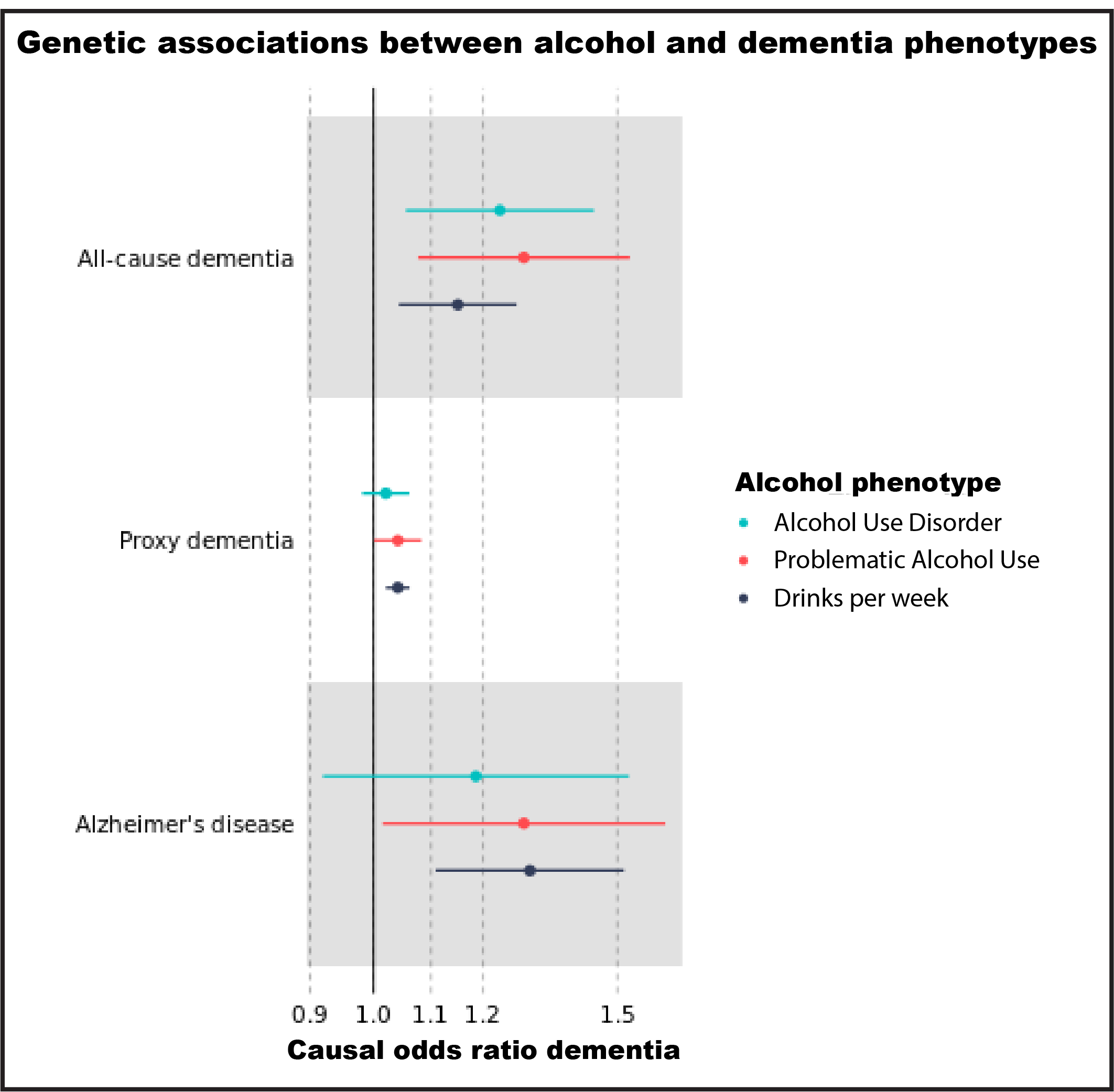

Estimates show odds ratios for dementia (95% confidence intervals) for a 1 standard deviation increase in log drinks per week, or a 1-log unit increase in problematic or dependent alcohol use. All-cause dementia was defined in Million Veteran Program using ICD codes in the electronic health record (Table S2) and genetic associations were calculated *de novo* in this study. Proxy dementia was defined as a diagnosis of Alzheimer’s disease or a parental history of dementia, and genetic associations originated from Wightman et al. as combined clinical cases and parental history of dementia.^21^ Alzheimer’s disease was clinically diagnosed and genetic associations originated from Kunkle et al.^22^

#### Figure S18: Localized average causal effect estimates for five strata of alcohol exposure, from nonlinear Mendelian randomization.

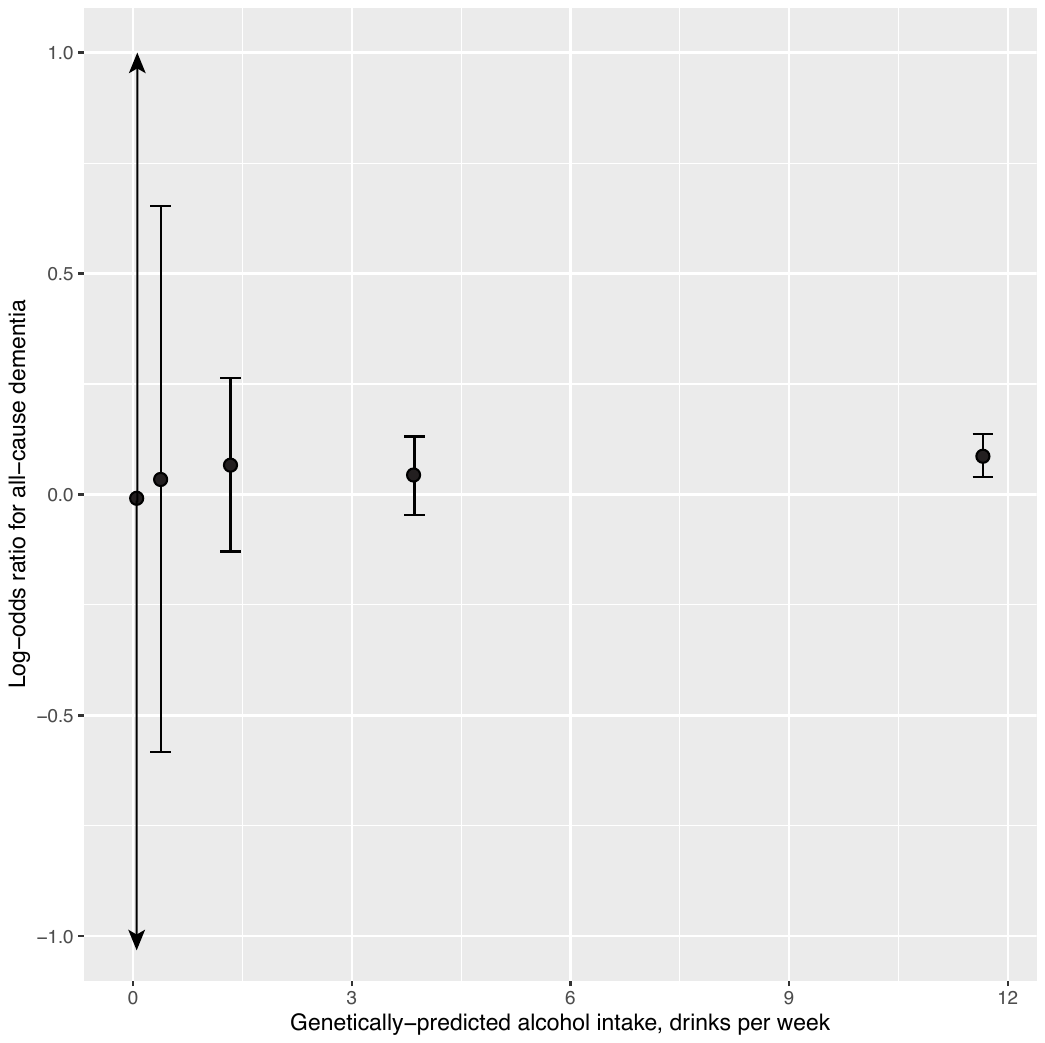

The doubly-ranked method was used for nonlinear Mendelian randomization. N=313,873 (16,932 dementia cases) Million Veteran Program participants, including non-drinkers, were included. X-axis shows the mean genetically-predicted alcohol intake in each stratum, the y-axis the association with all-cause dementia. Stratum 1 has the lowest average alcohol intake (0.1 drinks per week), and stratum 5 the highest (11.7 drinks per week). The points represent association estimates from logistic regression, adjusted for age, age^2^, sex and the top ten principal components for ancestry. Error bars represent 95% confidence intervals.

#### Figure S19: Localized average causal effect estimates for five strata of alcohol exposure, from nonlinear Mendelian randomization, excluding non-drinkers.

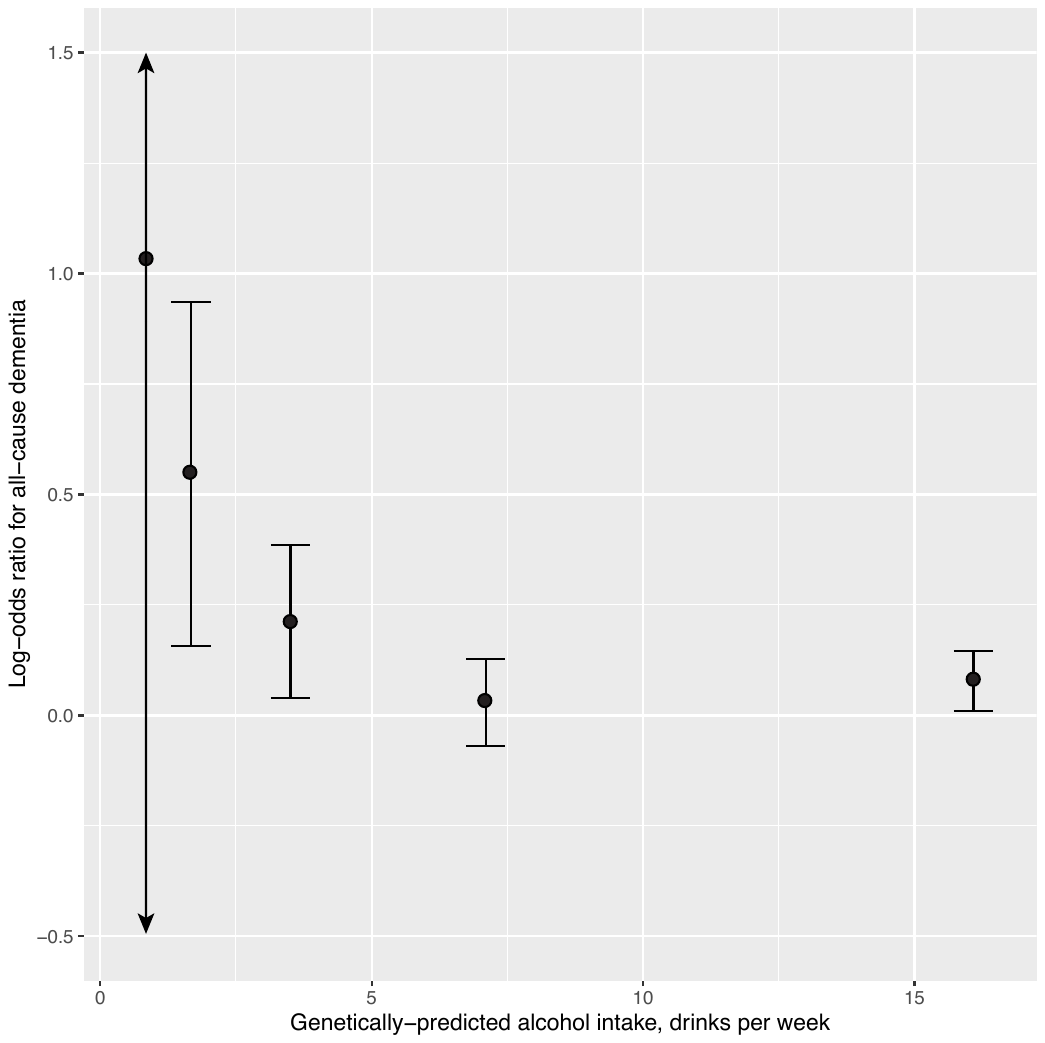

The doubly-ranked method was used for nonlinear Mendelian randomization. Million Veteran Program participants (n=185,733), excluding nondrinkers. X-axis shows the mean genetically-predicted alcohol intake in each stratum, the y-axis the association with all-cause dementia. Stratum 1 has the lowest average alcohol intake (0.9 drinks per week), and stratum 5 the highest (16.1 drinks per week). The points represent association estimates from logistic regression. Error bars represent 95% confidence intervals.

#### Figure S20: Nonlinear Mendelian randomization estimation of the dose-response curve between alcoholic drinks per week and all-cause dementia, excluding non-drinkers.

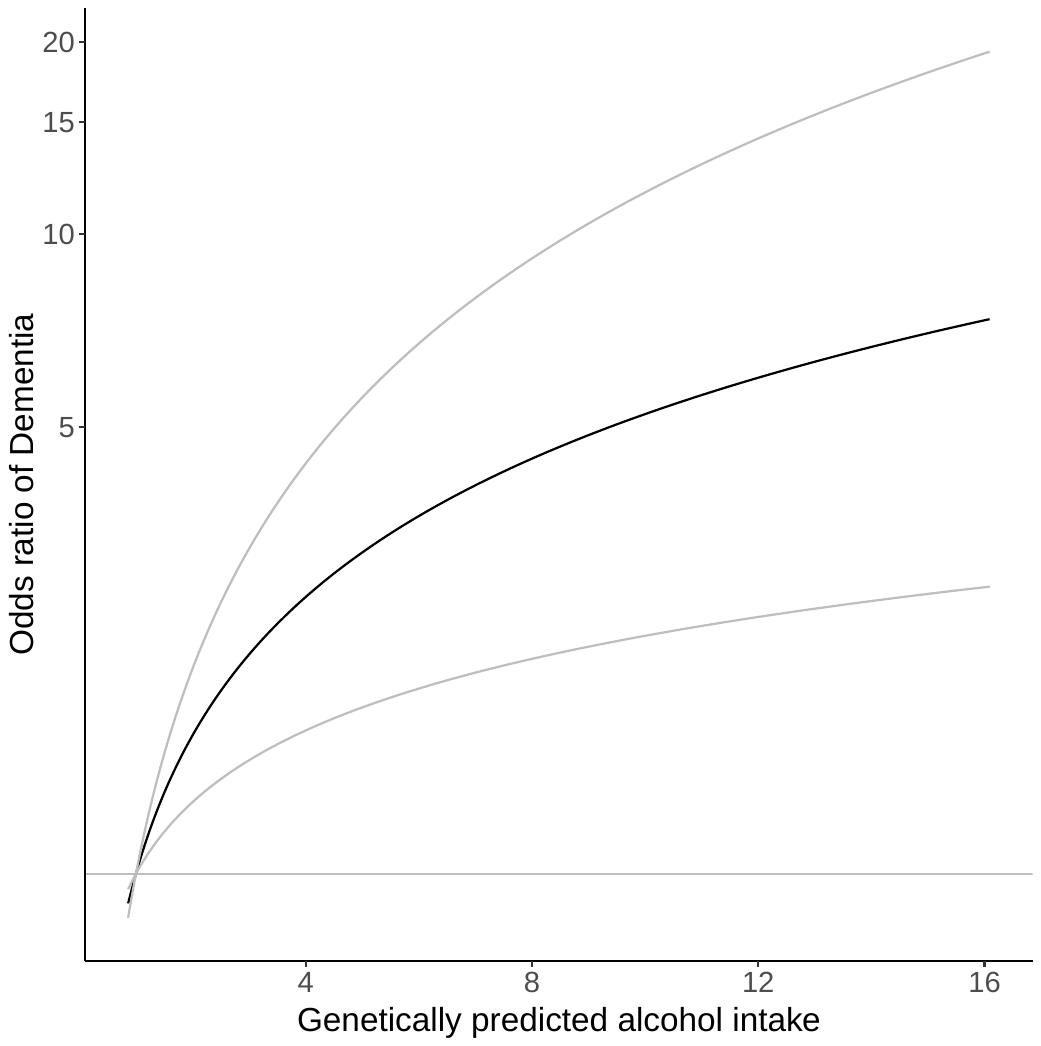

Nonlinear Mendelian randomization was performed using the doubly-ranked method, in unrelated European ancestry Million Veteran Program participants (n=185,733), excluding nondrinkers. The gradient at each point of the curve is the localized average causal effect. The x-axis shows the alcohol intake in drinks per week. The y-axis shows the odds ratio for the respective all-cause dementia risk. Grey lines represent the 95% confidence intervals. The reference value for alcohol intake was taken as 1 drink per week. P-value for linearity: 0.3, p-value for trend: 0.01.

#### Figure S21: Predicted causal effect of alcoholic drinks per week on age as a negative control.

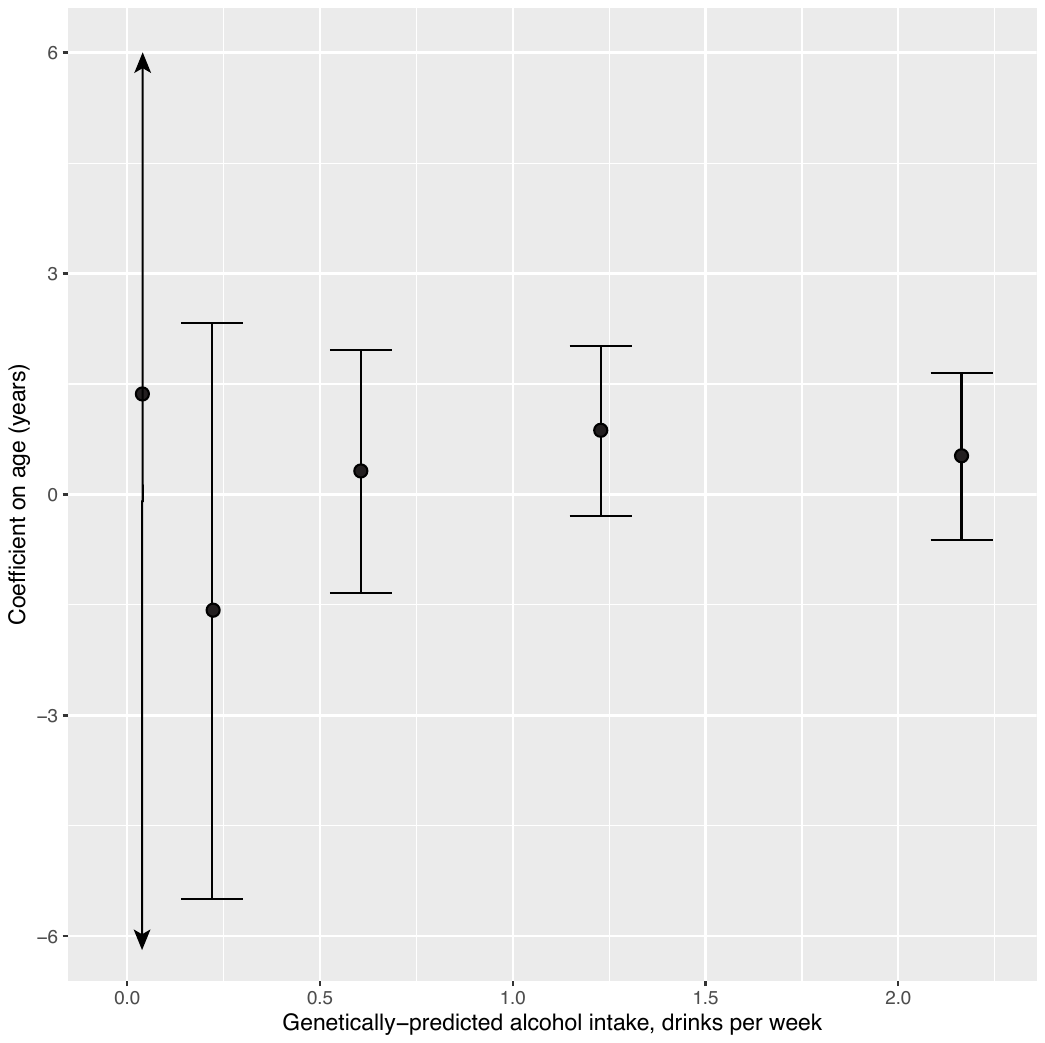

Strata were generated using the doubly-ranked method. Estimates (detailed in Table S26) were calculated using linear regression and adjusted for sex and the first ten genetic principal components. N=313,873 (16,932 dementia cases) were included in the analyses.

#### Figure S22: Predicted causal effect of alcoholic drinks per week on sex as a negative control.

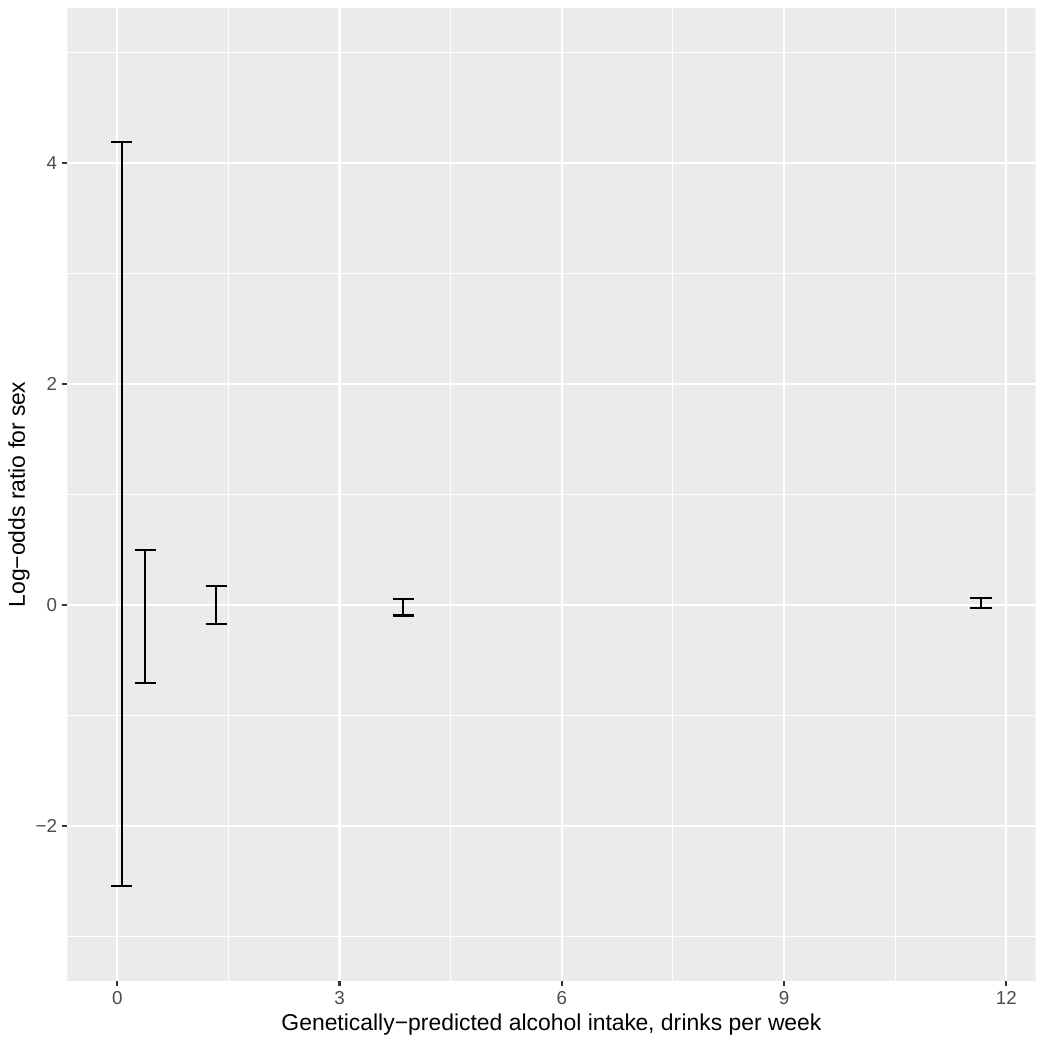

Strata were generated using the doubly-ranked method. Estimates (detailed in Table S27) were calculated using logistic regression and adjusted for age and the first ten genetic principal components. N=313,873 (16,932 dementia cases) were included in the analyses.

### Supplementary tables

#### Table S1: Source genome-wide association studies for genetic associations.

| **Phenotype** | **Traits** | **Cohorts** | **Ntotal** | **Neffective** | **Data source and original publication where relevant** |
| --- | --- | --- | --- | --- | --- |
| **Alcohol Use Disorder** | AUD | Million Veteran Program | 448,141 | 262,947 | European ancestry, Zhou et al., 2023 |
|  | AUD | FinnGen | 218,792 | 34,027 |  |
|  | AUD | Psychiatric Genetics Consortium | 40,930 | 23,075 |  |
|  | AUD | QIMR-AGDS | 11,193 | 10,737 |  |
|  | AUD | QIMR-TWINS | 8,402 | 7,430 |  |
|  | AUD | QIMR-GBD | 2,038 | 1,897 |  |
|  | AUD | iPSYCH 1 | 15,355 | 7,301 |  |
|  | AUD | iPSYCH 2 | 6,756 | 3,475 |  |
|  | AUD | Yale-Penn 3 | 1,641 | 1,484 |  |
| **Alcohol Use Disorder** | AUD | MVP | 115,430 | 99,583 | African ancestry, Zhou et al., 2023 |
|  | AD | PGC | 6,280 | 4,991 |  |
|  | AD | YP3 | 861 | 959 |  |
|  | AUD | Million Veteran Program | 115,430 | 99,583 |  |
| **Problematic Alcohol Use** | AUDIT-P | UK Biobank | 149899 | 149899 | European ancestry, Zhou et al., 2023 |
|  | AUD | FinnGen | 218,792 | 34,027 |  |
|  | AUD | Psychiatric Genetics Consortium | 40,930 | 23,075 |  |
|  | AUD | QIMR-AGDS | 11,193 | 10,737 |  |
|  | AUD | QIMR-TWINS | 8,402 | 7,430 |  |
|  | AUD | QIMR-GBD | 2,038 | 1,897 |  |
|  | AUD | iPSYCH 1 | 15,355 | 7,301 |  |
|  | AUD | iPSYCH 2 | 6,756 | 3,475 |  |
|  | AUD | Yale-Penn 3 | 1,641 | 1,484 |  |
| **Drinks per week** | DPW | 23andMe - EUR Males | 801,737 |  | European ancestry, Saunders at al., 2022 |
|  | DPW | 23andMe - EUR Females | 960,136 |  |  |
|  | DPW | ALSPAC | 8,913 |  |  |
|  | DPW | ARIC - TOPMed | 6,250 |  |  |
|  | DPW | CADD | 916 |  |  |
|  | DPW | COGEND | 1,924 |  |  |
|  | DPW | deCODE | 36,818 |  |  |
|  | DPW | EGCUT | 39,046 |  |  |
|  | DPW | FinnTwin 1 | 969 |  |  |
|  | DPW | FinnTwin 2 | 7,264 |  |  |
|  | DPW | Framingham - TOPMed | 5,105 |  |  |
|  | DPW | Genes for Good | 4,286 |  |  |
|  | DPW | GERA | 47,967 |  |  |
|  | DPW | Harvard - Affy | 4,479 |  |  |
|  | DPW | Harvard - Illumina | 4,118 |  |  |
|  | DPW | Harvard - HumanCore | 4,597 |  |  |
|  | DPW | Harvard - OmniExpress | 4,965 |  |  |
|  | DPW | Harvard - OncoArray | 5,309 |  |  |
|  | DPW | HRS | 5,635 |  |  |
|  | DPW | HUNT | 47,325 |  |  |
|  | DPW | MCTFR | 3,899 |  |  |
|  | DPW | MESA - TOPMed | 1,632 |  |  |
|  | DPW | METSIM | 7,403 |  |  |
|  | DPW | NESCOG | 429 |  |  |
|  | DPW | NAG-FIN | 1,697 |  |  |
|  | DPW | NTR | 5,084 |  |  |
|  | DPW | QIMR | 9,947 |  |  |
|  | DPW | SardiNIA | 2,570 |  |  |
|  | DPW | UKB | 362,656 |  |  |
|  | DPW | FINRISK | 16,855 |  |  |
|  | DPW | WLS | 6,194 |  |  |
|  | DPW | Spit for Science | 2,314 |  |  |
|  | DPW | WHI - MOPMAP | 1,406 |  |  |
|  | DPW | WHI - HIPFX | 1,912 |  |  |
|  | DPW | WHI - GARNET | 2,192 |  |  |
|  | DPW | WHI - LLS | 671 |  |  |
|  | DPW | WHI - GECCO | 941 |  |  |
|  | DPW | WHI - WHIMS+ | 3,290 |  |  |
| **All-cause dementia** | ICD 9/10 code | Million Veteran Program | 451317 |  | European ancestry |
| **All-cause dementia** | ICD 9/10 code | Million Veteran Program | 114238 |  | African ancestry |
| **Proxy dementia** | Clinically diagnosed Alzheimer's | IGAP | 63926 |  | European ancestry, Wightman et al., 2021 |
|  | Proxy dementia | UKB (Proxy) | 364859 |  |  |
|  | Clinically diagnosed Alzheimer's | DemGene | 7697 |  |  |
|  | Clinically diagnosed Alzheimer's | TwinGene | 6545 |  |  |
|  | Clinically diagnosed Alzheimer's | STSA | 1070 |  |  |
|  | Clinically diagnosed Alzheimer's | deCODE (partially included) | 188575 |  |  |
|  | ICD 10 code Alzheimer's | FinnGen | 74004 |  |  |
|  | Clinically diagnosed | GR@CE | 7409 |  |  |
|  | ICD 9/10 code Alzheimer's | HUNT | 8313 |  |  |
|  | ICD 9/10 code Alzheimer's | BioVU | 36659 |  |  |
|  | Self-reported Alzheimer's | 23andMe | 363646 |  |  |
|  | Clinically diagnosed Alzheimer's | Gothenburg H70 Birth Cohort Studies and Clinical AD from Sweden (Gothenburg) | 3235 |  |  |
|  | Clinically diagnosed Alzheimer's | ANMerge | 625 |  |  |
| **Clinically diagnosed Alzheimer's** | Clinically diagnosed Alzheimer's | ADGC | 28990* |  | European ancestry, Kunkle et al., 2019 |
|  | Clinically diagnosed Alzheimer's | CHARGE | 15,611 |  |  |
|  | Clinically diagnosed Alzheimer's | EADI | 8,871 |  |  |
|  | Clinically diagnosed Alzheimer's | GERAD | 10,454 |  |  |
| **Cannabis Use** |  | Million Veterans Program |  |  | Levey et al., 2023 |
| **PTSD symptoms** | PCL | Million Veterans Program | 214,408 |  | Stein et al., 2021 |
| **Smoking** | Cigarettes daily | GSCAN | 784,353 |  | Saunders et al., 2023 |
| **Income** | Household income | UK Biobank | 286,301 |  | Hill et al., 2019 |
| **Neuroimaging phenotypes** | Global brain volume, hippocampal volume, white matter hyperintensity volume | UK Biobank | 39,691 |  | Smith et al., 2021 |

| *Stage 1 sample |
| --- |
| Abbreviations: PTSD - post-traumatic stress disorder, AUD - alcohol use disorder, AD - alcohol dependence, DPW - drinks per week, PCL - PTSD checklist. |

#### Table S2: International Classification of Disease codes used to define dementia.

|  | **ICD 9 codes** | **ICD 10 codes** |
| --- | --- | --- |
| **All-cause dementia** | 290.0-290.4; 291.2; 294.1; 331.0-2; 331.5; 331.82; 331.9; 332 | F00-F03; F05.1; F1x.73; G30-G31; I67.3; R54; F10.73; G31.2; F00.2; F01; I67.3; I60-I64; I69; Z86.70; G45; Z86.60; F1x.74; F06.7 |
| **Alzheimer's disease** | 331 | G30.0, G30.1, G30.8, G30.9 |
| **Vascular dementia** | 290.40, 290.41, 290.42 | F01.50, F01.51 |

#### Table S3: Alcohol use disorder instruments p<5x10^-8^.

| **Ancestry** | **SNP** | **Chromosome** | **Position** | **Effect allele** | **Other allele** | **EAF** | **Beta** | **SE** | **P value** | **F statistic** |
| --- | --- | --- | --- | --- | --- | --- | --- | --- | --- | --- |
| EUR | rs10253153 | 7 | 1001974 | A | G | 0.2611 | -0.02 | 2.72E-03 | 8.87E-09 | 33.07 |
| EUR | rs10630359 | 14 | 58767085 | A | ATATACT | 0.301 | 0.02 | 2.74E-03 | 8.99E-11 | 42.03 |
| EUR | rs11214596 | 11 | 113264511 | C | G | 0.4774 | 0.02 | 2.38E-03 | 7.22E-11 | 42.46 |
| EUR | rs11366076 | 9 | 17237272 | T | TA | 0.1661 | -0.02 | 3.37E-03 | 3.91E-09 | 34.67 |
| EUR | rs11468270 | 20 | 37306245 | G | GTGGACTGATT | 0.6833 | -0.02 | 2.64E-03 | 1.38E-09 | 36.70 |
| EUR | rs1155397 | 7 | 114953597 | A | G | 0.3281 | 0.02 | 2.54E-03 | 4.64E-11 | 43.32 |
| EUR | rs117623407 | 19 | 32204489 | A | G | 0.8538 | 0.02 | 3.42E-03 | 1.19E-09 | 36.98 |
| EUR | rs1229984 | 4 | 100239319 | T | C | 0.0341 | -0.20 | 6.65E-03 | 1.81E-204 | 930.98 |
| EUR | rs1248850 | 3 | 84887559 | T | C | 0.5106 | 0.01 | 2.38E-03 | 5.33E-10 | 38.55 |
| EUR | rs1260326 | 2 | 27730940 | T | C | 0.405 | -0.02 | 2.43E-03 | 3.63E-22 | 93.72 |
| EUR | rs12620018 | 2 | 73906408 | A | G | 0.6363 | -0.01 | 2.49E-03 | 2.89E-09 | 35.26 |
| EUR | rs1291854 | 10 | 11110195 | T | C | 0.4409 | -0.01 | 2.40E-03 | 2.14E-08 | 31.36 |
| EUR | rs13107325 | 4 | 103188709 | T | C | 0.0739 | -0.05 | 4.58E-03 | 3.77E-33 | 143.88 |
| EUR | rs13146907 | 4 | 39425248 | A | G | 0.6183 | 0.02 | 2.48E-03 | 1.29E-20 | 86.66 |
| EUR | rs13276082 | 8 | 21823184 | A | G | 0.5777 | 0.01 | 2.41E-03 | 4.57E-09 | 34.36 |
| EUR | rs1327631 | 13 | 96848503 | A | C | 0.4997 | -0.01 | 2.38E-03 | 3.98E-07 | 25.70 |
| EUR | rs1405238 | 14 | 99733954 | T | C | 0.5889 | 0.01 | 2.44E-03 | 2.92E-08 | 30.76 |
| EUR | rs1421085 | 16 | 53800954 | T | C | 0.5923 | 0.02 | 2.42E-03 | 6.69E-19 | 78.85 |
| EUR | rs1822717 | 8 | 64956228 | T | C | 0.4805 | 0.02 | 2.38E-03 | 3.31E-11 | 43.98 |
| EUR | rs1921035 | 12 | 81602449 | A | G | 0.5209 | 0.01 | 2.38E-03 | 2.10E-09 | 35.88 |
| EUR | rs192979726 | 10 | 110732490 | T | TAGAG | 0.17 | -0.02 | 3.36E-03 | 1.22E-08 | 32.46 |
| EUR | rs2002094 | 17 | 28044991 | A | C | 0.0859 | -0.02 | 4.35E-03 | 1.03E-08 | 32.79 |
| EUR | rs200588406 | 11 | 57498460 | CTTAAA | C | 0.6589 | -0.02 | 2.65E-03 | 6.89E-11 | 42.55 |
| EUR | rs2022028 | 1 | 44606565 | A | G | 0.3778 | -0.02 | 2.46E-03 | 1.70E-10 | 40.78 |
| EUR | rs2059924 | 2 | 185785947 | T | G | 0.3966 | 0.01 | 2.43E-03 | 3.33E-08 | 30.50 |
| EUR | rs2098112 | 7 | 153487944 | A | G | 0.4653 | 0.01 | 2.51E-03 | 3.18E-09 | 35.07 |
| EUR | rs2310819 | 1 | 66440096 | T | C | 0.5361 | 0.02 | 2.39E-03 | 1.54E-11 | 45.48 |
| EUR | rs2576589 | 8 | 57425647 | T | C | 0.7522 | -0.02 | 2.76E-03 | 1.14E-09 | 37.08 |
| EUR | rs2683632 | 2 | 58116640 | T | C | 0.6118 | -0.02 | 2.44E-03 | 5.03E-11 | 43.17 |
| EUR | rs2696884 | 7 | 135144662 | T | C | 0.4015 | 0.02 | 2.43E-03 | 8.85E-14 | 55.61 |
| EUR | rs2784867 | 6 | 163840529 | A | G | 0.6733 | 0.01 | 2.54E-03 | 4.04E-08 | 30.13 |
| EUR | rs3217512 | 11 | 64135541 | G | GTC | 0.3808 | -0.01 | 2.51E-03 | 4.38E-08 | 29.98 |
| EUR | rs34488670 | 15 | 47684936 | T | C | 0.7845 | -0.02 | 2.90E-03 | 5.11E-14 | 56.69 |
| EUR | rs35942385 | 2 | 144208523 | T | G | 0.3554 | -0.02 | 2.49E-03 | 8.15E-16 | 64.83 |
| EUR | rs372701525 | 1 | 80860383 | A | C | 0.2775 | 0.02 | 2.81E-03 | 1.66E-08 | 31.85 |
| EUR | rs3781624 | 11 | 47443251 | T | C | 0.3193 | -0.02 | 2.61E-03 | 7.70E-10 | 37.83 |
| EUR | rs403598 | 20 | 31634788 | A | G | 0.0528 | -0.03 | 5.64E-03 | 4.40E-08 | 29.96 |
| EUR | rs439523 | 19 | 49236102 | T | C | 0.4966 | -0.02 | 2.39E-03 | 3.92E-11 | 43.65 |
| EUR | rs4546191 | 4 | 139847208 | A | G | 0.7974 | -0.02 | 2.96E-03 | 3.75E-10 | 39.24 |
| EUR | rs472140 | 2 | 45139904 | T | C | 0.586 | 0.02 | 2.43E-03 | 2.08E-18 | 76.62 |
| EUR | rs4761975 | 12 | 51880523 | T | C | 0.5246 | -0.01 | 2.39E-03 | 1.52E-09 | 36.51 |
| EUR | rs4838249 | 9 | 127911031 | A | G | 0.7071 | 0.02 | 2.62E-03 | 1.62E-09 | 36.39 |
| EUR | rs542448643 | 20 | 31634788 | A | G | 0.0528 | -0.03 | 5.64E-03 | 4.40E-08 | 29.96 |
| EUR | rs543725 | 11 | 57483039 | A | T | 0.6723 | -0.02 | 2.73E-03 | 2.81E-12 | 48.82 |
| EUR | rs56009681 | 4 | 100285772 | A | T | 0.8965 | -0.05 | 3.92E-03 | 1.47E-42 | 186.95 |
| EUR | rs566835533 | 2 | 27667520 | T | TTTTG | 0.5614 | -0.02 | 2.54E-03 | 2.40E-16 | 67.24 |
| EUR | rs567678299 | 3 | 49088127 | A | AAAC | 0.747 | -0.02 | 2.97E-03 | 3.70E-09 | 34.77 |
| EUR | rs572635656 | 16 | 53799278 | G | GT | 0.5787 | 0.02 | 2.54E-03 | 1.10E-18 | 77.86 |
| EUR | rs57761252 | 2 | 161865998 | A | G | 0.2047 | -0.02 | 2.99E-03 | 2.02E-08 | 31.47 |
| EUR | rs59899558 | 13 | 89072227 | T | G | 0.6665 | 0.02 | 2.73E-03 | 1.62E-09 | 36.39 |
| EUR | rs60427825 | 1 | 73852274 | A | T | 0.4124 | 0.01 | 2.48E-03 | 4.46E-09 | 34.41 |
| EUR | rs60612094 | 16 | 30045905 | CAAT | C | 0.4626 | -0.02 | 2.52E-03 | 1.19E-11 | 46.00 |
| EUR | rs6442644 | 3 | 16852853 | T | G | 0.6885 | -0.02 | 2.80E-03 | 5.71E-09 | 33.93 |
| EUR | rs6449591 | 5 | 61535659 | A | G | 0.5836 | 0.01 | 2.48E-03 | 1.81E-09 | 36.17 |
| EUR | rs6589386 | 11 | 113443753 | T | C | 0.4132 | -0.02 | 2.42E-03 | 8.55E-20 | 82.92 |
| EUR | rs6809204 | 3 | 49369383 | T | C | 0.7328 | -0.02 | 2.69E-03 | 6.58E-14 | 56.19 |
| EUR | rs7093143 | 10 | 110478002 | T | G | 0.7492 | 0.02 | 2.75E-03 | 1.05E-12 | 50.75 |
| EUR | rs7106615 | 11 | 121632466 | A | G | 0.7813 | 0.02 | 2.88E-03 | 1.77E-14 | 58.77 |
| EUR | rs7177599 | 15 | 83717246 | T | G | 0.4197 | -0.01 | 2.41E-03 | 1.36E-08 | 32.24 |
| EUR | rs7272308 | 20 | 48583726 | A | G | 0.2782 | 0.01 | 2.66E-03 | 3.65E-08 | 30.33 |
| EUR | rs72859290 | 2 | 147981914 | A | G | 0.0376 | 0.04 | 6.30E-03 | 2.32E-09 | 35.69 |
| EUR | rs73591198 | 13 | 108213021 | A | G | 0.0938 | -0.02 | 4.10E-03 | 2.19E-08 | 31.32 |
| EUR | rs7531138 | 1 | 97923499 | A | T | 0.4484 | -0.01 | 2.40E-03 | 2.10E-09 | 35.88 |
| EUR | rs766406 | 6 | 26319588 | T | G | 0.6422 | -0.02 | 2.48E-03 | 1.67E-10 | 40.82 |
| EUR | rs850254 | 14 | 57350427 | A | G | 0.5527 | -0.01 | 2.40E-03 | 1.43E-08 | 32.15 |
| EUR | rs854784 | 17 | 18040690 | T | C | 0.5388 | 0.02 | 2.39E-03 | 2.42E-10 | 40.09 |
| AFR | rs1229984 | 4 | 100239319 | T | C | 0.0107 | -0.17 | 2.13E-02 | 1.24E-15 | 64.00 |
| AFR | rs2066702 | 4 | 100229017 | A | G | 0.1907 | -0.09 | 5.54E-03 | 8.47E-57 | 252.00 |
| AFR | rs534756470 | 4 | 100057858 | T | TA | 0.8934 | -0.04 | 7.08E-03 | 1.79E-08 | 31.70 |
| AFR | rs55853380 | 4 | 100273668 | T | TTCTAATTTAGATATGAATTTTAAATTATTCATCA | 0.0257 | 0.08 | 1.39E-02 | 2.69E-08 | 30.90 |

Abbreviations: SNP - single nucleotide polymorphism, EAF - effect allele frequency, SE - standard error, EUR - European, AFR - African American.

Source genome-wide association study was Zhou et al.^23^

Multi-allelic instruments were included given that all datasets clearly report multiple alleles allowing comparison.

#### Table S4: Alcohol use disorder instruments, p<5x10^-5^, African ancestry.

| **SNP** | **Chromosome** | **Position** | **Effect allele** | **Other allele** | **EAF** | **Beta** | **SE** | **P value** | **F statistic** |
| --- | --- | --- | --- | --- | --- | --- | --- | --- | --- |
| rs10036298 | 5 | 112865189 | C | A | 0.55 | 2.00E-02 | 4.38E-03 | 1.15E-07 | 28.10 |
| rs10053730 | 5 | 100950397 | C | A | 0.38 | 2.00E-02 | 4.47E-03 | 5.40E-06 | 20.69 |
| rs10121722 | 9 | 6174048 | A | C | 0.02 | -7.00E-02 | 1.61E-02 | 2.52E-05 | 17.75 |
| rs10934311 | 3 | 115582025 | A | G | 0.33 | 2.00E-02 | 4.65E-03 | 5.56E-07 | 25.06 |
| rs10941268 | 5 | 35923689 | G | A | 0.08 | 4.00E-02 | 8.15E-03 | 5.68E-06 | 20.59 |
| rs11101086 | 10 | 50527064 | T | C | 0.05 | -4.00E-02 | 1.06E-02 | 2.74E-05 | 17.59 |
| rs111445299 | 2 | 37277831 | T | C | 0.03 | -6.00E-02 | 1.36E-02 | 4.40E-05 | 16.69 |
| rs114126697 | 20 | 31569412 | G | A | 0.02 | -8.00E-02 | 1.79E-02 | 9.55E-06 | 19.60 |
| rs114438914 | 2 | 132245205 | T | C | 0.02 | 8.00E-02 | 1.69E-02 | 1.94E-06 | 22.65 |
| rs114461676 | 7 | 100840326 | C | A | 0.03 | -6.00E-02 | 1.37E-02 | 6.26E-06 | 20.41 |
| rs114530819 | 21 | 46374059 | T | G | 0.05 | 4.00E-02 | 1.02E-02 | 2.56E-05 | 17.72 |
| rs114561376 | 9 | 125226597 | G | A | 0.03 | 6.00E-02 | 1.38E-02 | 4.08E-05 | 16.83 |
| rs114843286 | 1 | 53838405 | G | A | 0.03 | -6.00E-02 | 1.38E-02 | 3.41E-05 | 17.17 |
| rs114854961 | 12 | 98787763 | C | T | 0.03 | 6.00E-02 | 1.35E-02 | 4.05E-05 | 16.85 |
| rs115148403 | 22 | 45206935 | C | G | 0.06 | 4.00E-02 | 9.30E-03 | 4.08E-05 | 16.83 |
| rs115158582 | 10 | 65437789 | C | T | 0.04 | 5.00E-02 | 1.04E-02 | 8.99E-06 | 19.71 |
| rs115414568 | 2 | 53879110 | T | C | 0.01 | 9.00E-02 | 1.97E-02 | 1.01E-05 | 19.50 |
| rs115496212 | 4 | 110526447 | G | A | 0.04 | 5.00E-02 | 1.07E-02 | 2.52E-05 | 17.75 |
| rs115734449 | 3 | 36846635 | C | T | 0.01 | 9.00E-02 | 2.09E-02 | 3.83E-05 | 16.96 |
| rs115946221 | 5 | 30073025 | A | T | 0.02 | 6.00E-02 | 1.54E-02 | 4.27E-05 | 16.74 |
| rs116030331 | 14 | 52336896 | C | T | 0.02 | 7.00E-02 | 1.61E-02 | 3.61E-05 | 17.07 |
| rs116092999 | 8 | 60839511 | G | C | 0.03 | 6.00E-02 | 1.32E-02 | 2.74E-06 | 22.00 |
| rs116390638 | 10 | 13111394 | C | G | 0.04 | -5.00E-02 | 1.13E-02 | 3.00E-05 | 17.41 |
| rs11952141 | 5 | 18024800 | A | G | 0.35 | 2.00E-02 | 4.57E-03 | 2.82E-05 | 17.54 |
| rs12005444 | 9 | 137999047 | G | A | 0.01 | -9.00E-02 | 2.10E-02 | 1.38E-05 | 18.90 |
| rs12049376 | 1 | 73764712 | G | A | 0.39 | -2.00E-02 | 4.46E-03 | 2.73E-06 | 22.00 |
| rs12124206 | 1 | 216423098 | T | C | 0.02 | -6.00E-02 | 1.50E-02 | 2.78E-05 | 17.56 |
| rs1229984 | 4 | 100239319 | T | C | 0.99 | -1.70E-01 | 2.13E-02 | 1.24E-15 | 64.00 |
| rs12747999 | 1 | 228796705 | C | A | 0.04 | 5.00E-02 | 1.15E-02 | 2.82E-05 | 17.54 |
| rs12883188 | 14 | 39327372 | G | A | 0.1 | -3.00E-02 | 7.46E-03 | 8.27E-06 | 19.87 |
| rs12942912 | 17 | 43795899 | G | A | 0.12 | 3.00E-02 | 6.74E-03 | 8.14E-06 | 19.91 |
| rs1359711 | 9 | 113578430 | G | A | 0.05 | 4.00E-02 | 1.01E-02 | 1.59E-05 | 18.64 |
| rs138745179 | 11 | 1907623 | CA | C | 0.04 | -5.00E-02 | 1.16E-02 | 1.64E-05 | 18.57 |
| rs139028827 | 4 | 1130970 | A | G | 0.02 | -6.00E-02 | 1.51E-02 | 2.40E-05 | 17.84 |
| rs139992632 | 16 | 17117248 | G | T | 0.01 | -8.00E-02 | 2.02E-02 | 4.58E-05 | 16.61 |
| rs140012947 | 8 | 39831417 | C | T | 0.08 | 3.00E-02 | 8.11E-03 | 2.11E-05 | 18.09 |
| rs141128408 | 14 | 78276242 | G | A | 0.03 | -6.00E-02 | 1.37E-02 | 3.20E-05 | 17.30 |
| rs141456449 | 10 | 110824689 | T | C | 0.02 | -8.00E-02 | 1.78E-02 | 1.97E-05 | 18.22 |
| rs141665614 | 4 | 30635275 | C | T | 0.01 | 8.00E-02 | 1.89E-02 | 2.84E-05 | 17.52 |
| rs142078973 | 4 | 146376867 | T | C | 0.01 | -9.00E-02 | 2.06E-02 | 2.70E-05 | 17.61 |
| rs142369630 | 1 | 94878145 | A | G | 0.01 | -8.00E-02 | 2.20E-02 | 0.00051 | 12.08 |
| rs142609658 | 2 | 209331927 | T | A | 0.03 | -5.00E-02 | 1.24E-02 | 2.69E-05 | 17.62 |
| rs143684057 | 12 | 70542997 | G | A | 0.03 | 6.00E-02 | 1.34E-02 | 1.85E-05 | 18.34 |
| rs143941899 | 11 | 35466092 | T | C | 0.03 | -6.00E-02 | 1.39E-02 | 2.84E-05 | 17.52 |
| rs144062959 | 8 | 90852550 | T | C | 0.02 | -7.00E-02 | 1.63E-02 | 2.38E-05 | 17.86 |
| rs144637117 | 3 | 142601785 | G | A | 0.03 | 5.00E-02 | 1.29E-02 | 2.66E-05 | 17.65 |
| rs145014179 | 3 | 73634710 | C | T | 0.02 | -8.00E-02 | 1.62E-02 | 1.11E-06 | 23.73 |
| rs145763437 | 7 | 103478259 | C | T | 0.02 | 7.00E-02 | 1.66E-02 | 4.90E-05 | 16.48 |
| rs147457614 | 1 | 225876447 | C | T | 0.02 | 8.00E-02 | 1.69E-02 | 3.23E-06 | 21.67 |
| rs148680916 | 8 | 11727762 | G | T | 0.04 | 5.00E-02 | 1.17E-02 | 2.79E-05 | 17.56 |
| rs150314176 | 2 | 21147421 | G | A | 0.02 | -8.00E-02 | 1.73E-02 | 1.45E-05 | 18.80 |
| rs180730110 | 5 | 107252750 | G | T | 0.01 | 8.00E-02 | 1.93E-02 | 3.31E-05 | 17.23 |
| rs1815354 | 13 | 96849493 | A | T | 0.62 | -2.00E-02 | 4.50E-03 | 1.27E-06 | 23.46 |
| rs181886745 | 19 | 5814025 | T | A | 0.02 | -8.00E-02 | 1.77E-02 | 1.30E-05 | 19.01 |
| rs182879050 | 15 | 101027191 | T | C | 0.02 | 8.00E-02 | 1.72E-02 | 2.37E-06 | 22.27 |
| rs186175237 | 19 | 46012522 | C | G | 0.02 | 8.00E-02 | 1.80E-02 | 8.21E-06 | 19.89 |
| rs188193183 | 3 | 192989590 | C | T | 0.02 | -7.00E-02 | 1.73E-02 | 1.50E-05 | 18.74 |
| rs189713116 | 2 | 103967040 | T | G | 0.02 | -8.00E-02 | 1.71E-02 | 8.60E-06 | 19.80 |
| rs199612276 | 6 | 121479308 | TC | T | 0.28 | 2.00E-02 | 4.88E-03 | 3.08E-05 | 17.37 |
| rs200589817 | 5 | 85844083 | CCTT | C | 0.02 | -7.00E-02 | 1.69E-02 | 1.38E-05 | 18.90 |
| rs200907192 | 2 | 185158876 | A | AT | 0.05 | 4.00E-02 | 1.03E-02 | 4.69E-05 | 16.57 |
| rs201298067 | 5 | 168608465 | GC | G | 0.02 | -7.00E-02 | 1.61E-02 | 1.76E-05 | 18.43 |
| rs202200286 | 5 | 52792978 | CA | C | 0.04 | -5.00E-02 | 1.18E-02 | 1.95E-05 | 18.24 |
| rs2066702 | 4 | 100229017 | G | A | 0.19 | -9.00E-02 | 5.54E-03 | 8.47E-57 | 252.24 |
| rs2221460 | 1 | 75649654 | T | C | 0.25 | -2.00E-02 | 5.02E-03 | 1.43E-05 | 18.83 |
| rs2224198 | 6 | 52605909 | A | G | 0.93 | 4.00E-02 | 9.01E-03 | 7.95E-06 | 19.95 |
| rs229574 | 14 | 65226915 | T | G | 0.43 | -2.00E-02 | 4.47E-03 | 2.32E-06 | 22.32 |
| rs2409667 | 8 | 10729623 | A | G | 0.34 | -2.00E-02 | 4.60E-03 | 3.09E-05 | 17.36 |
| rs2547123 | 19 | 16802557 | G | A | 0.18 | -3.00E-02 | 5.75E-03 | 3.65E-06 | 21.44 |
| rs2667096 | 4 | 142969773 | G | T | 0.19 | -2.00E-02 | 5.55E-03 | 2.23E-05 | 17.98 |
| rs28451506 | 21 | 19490068 | T | A | 0.17 | 3.00E-02 | 5.89E-03 | 2.17E-06 | 22.44 |
| rs28988573 | 7 | 99380301 | G | C | 0.04 | -5.00E-02 | 1.15E-02 | 4.38E-05 | 16.70 |
| rs323878 | 3 | 51999847 | C | G | 0.12 | 3.00E-02 | 6.63E-03 | 1.08E-06 | 23.79 |
| rs340356 | 2 | 179948427 | T | C | 0.24 | 2.00E-02 | 5.10E-03 | 3.01E-05 | 17.41 |
| rs34296005 | 7 | 146817883 | A | T | 0.09 | -4.00E-02 | 7.59E-03 | 1.41E-06 | 23.26 |
| rs34569950 | 7 | 85836675 | A | T | 0.16 | 2.00E-02 | 5.92E-03 | 3.16E-05 | 17.32 |
| rs34670695 | 7 | 25608897 | TG | T | 0.04 | -5.00E-02 | 1.08E-02 | 6.19E-06 | 20.43 |
| rs34745087 | 17 | 7557394 | A | T | 0.03 | -5.00E-02 | 1.24E-02 | 3.34E-05 | 17.21 |
| rs35172302 | 6 | 97991869 | G | A | 0.08 | -3.00E-02 | 7.88E-03 | 4.98E-05 | 16.45 |
| rs359880 | 2 | 185439359 | T | C | 0.27 | -2.00E-02 | 4.91E-03 | 3.21E-05 | 17.29 |
| rs373444346 | 20 | 15928564 | T | G | 0.02 | 6.00E-02 | 1.56E-02 | 4.05E-05 | 16.84 |
| rs3809368 | 13 | 100624890 | G | A | 0.12 | 3.00E-02 | 6.82E-03 | 6.20E-06 | 20.42 |
| rs3851040 | 10 | 79318685 | C | T | 0.24 | -9.00E-02 | 2.07E-02 | 2.04E-05 | 18.16 |
| rs3910983 | 2 | 226742402 | A | G | 0.29 | -2.00E-02 | 4.80E-03 | 8.11E-06 | 19.91 |
| rs4477111 | 9 | 86884578 | C | T | 0.02 | -4.00E-02 | 1.68E-02 | 0.02644 | 4.93 |
| rs4482174 | 13 | 62613195 | T | C | 0.42 | -8.00E-02 | 1.86E-02 | 3.94E-05 | 16.90 |
| rs4820988 | 22 | 32065807 | G | A | 0.11 | 3.00E-02 | 7.02E-03 | 3.38E-05 | 17.19 |
| rs533017055 | 2 | 75035799 | G | A | 0.04 | -5.00E-02 | 1.14E-02 | 5.40E-06 | 20.69 |
| rs534756470 | 4 | 100057858 | T | TA | 0.11 | -4.00E-02 | 7.08E-03 | 1.79E-08 | 31.71 |
| rs537139946 | 11 | 89043109 | G | C | 0.01 | 1.00E-01 | 2.18E-02 | 7.40E-06 | 20.09 |
| rs538608853 | 15 | 84497860 | C | T | 0.02 | 6.00E-02 | 1.46E-02 | 4.32E-05 | 16.73 |
| rs541552624 | 6 | 13692774 | A | C | 0.02 | -7.00E-02 | 1.59E-02 | 2.14E-05 | 18.06 |
| rs543314539 | 8 | 66274001 | T | A | 0.02 | -6.00E-02 | 1.50E-02 | 4.77E-05 | 16.53 |
| rs546972304 | 16 | 31455651 | TG | T | 0.14 | 3.00E-02 | 6.26E-03 | 2.33E-05 | 17.89 |
| rs547455577 | 12 | 106976979 | T | G | 0.01 | 9.00E-02 | 2.03E-02 | 2.24E-05 | 17.98 |
| rs55853380 | 4 | 100273668 | TTCTAATTTAGATATGAATTTTAAATTATTCATCA | T | 0.03 | 8.00E-02 | 1.39E-02 | 2.69E-08 | 30.92 |
| rs55964790 | 7 | 70682535 | G | T | 0.11 | -3.00E-02 | 7.22E-03 | 4.22E-05 | 16.77 |
| rs56139883 | 7 | 78110371 | T | C | 0.05 | 4.00E-02 | 1.01E-02 | 8.07E-06 | 19.92 |
| rs56389751 | 12 | 107839142 | A | G | 0.01 | 8.00E-02 | 1.85E-02 | 3.13E-05 | 17.34 |
| rs57200947 | 11 | 116623674 | C | T | 0.17 | -3.00E-02 | 5.78E-03 | 1.49E-06 | 23.16 |
| rs572578713 | 5 | 79502814 | C | CTATA | 0.04 | 5.00E-02 | 1.10E-02 | 2.36E-05 | 17.88 |
| rs57985049 | 3 | 155169962 | A | G | 0.13 | 3.00E-02 | 6.54E-03 | 1.18E-05 | 19.20 |
| rs59537959 | 8 | 481195 | T | C | 0.03 | -5.00E-02 | 1.22E-02 | 2.67E-05 | 17.64 |
| rs5998557 | 22 | 33007038 | G | C | 0.53 | -2.00E-02 | 4.44E-03 | 3.20E-05 | 17.30 |
| rs6033569 | 20 | 12982750 | T | C | 0.02 | -6.00E-02 | 1.40E-02 | 2.15E-05 | 18.05 |
| rs6037463 | 20 | 3032272 | C | T | 0.85 | -3.00E-02 | 6.20E-03 | 3.35E-05 | 17.21 |
| rs6045222 | 20 | 1868173 | G | A | 0.07 | -4.00E-02 | 8.53E-03 | 1.48E-05 | 18.77 |
| rs61767666 | 1 | 4652923 | C | T | 0.03 | 6.00E-02 | 1.21E-02 | 1.55E-06 | 23.09 |
| rs61928263 | 12 | 50588659 | T | C | 0.06 | -4.00E-02 | 9.55E-03 | 1.27E-05 | 19.06 |
| rs62316542 | 4 | 95726170 | T | G | 0.03 | -6.00E-02 | 1.29E-02 | 1.37E-06 | 23.33 |
| rs6589894 | 11 | 121636060 | C | T | 0.24 | 2.00E-02 | 5.16E-03 | 6.84E-06 | 20.24 |
| rs680446 | 19 | 50789940 | G | A | 0.03 | -6.00E-02 | 1.20E-02 | 2.53E-06 | 22.15 |
| rs6889239 | 5 | 150457771 | T | C | 0.69 | 2.00E-02 | 4.71E-03 | 8.57E-06 | 19.80 |
| rs6958496 | 7 | 20694502 | G | C | 0.08 | 3.00E-02 | 8.00E-03 | 1.58E-05 | 18.64 |
| rs7070104 | 10 | 99345406 | A | G | 0.12 | -3.00E-02 | 6.84E-03 | 4.85E-05 | 16.51 |
| rs72903444 | 6 | 86096993 | C | A | 0.05 | 4.00E-02 | 1.01E-02 | 4.84E-05 | 16.51 |
| rs73220678 | 5 | 109231692 | T | C | 0.03 | -5.00E-02 | 1.25E-02 | 2.68E-05 | 17.63 |
| rs73403809 | 15 | 54833980 | A | G | 0.09 | -3.00E-02 | 7.75E-03 | 3.57E-05 | 17.09 |
| rs73631781 | 11 | 126153321 | C | G | 0.02 | 6.00E-02 | 1.42E-02 | 1.97E-05 | 18.22 |
| rs73723764 | 7 | 131951639 | A | T | 0.07 | 4.00E-02 | 8.57E-03 | 4.16E-06 | 21.19 |
| rs73792114 | 4 | 2976504 | G | A | 0.07 | -4.00E-02 | 8.72E-03 | 3.85E-05 | 16.94 |
| rs73976255 | 2 | 177946536 | C | T | 0.08 | 4.00E-02 | 8.10E-03 | 1.29E-05 | 19.04 |
| rs74153328 | 1 | 207782038 | A | G | 0.02 | 7.00E-02 | 1.41E-02 | 3.17E-06 | 21.71 |
| rs74444731 | 12 | 83974563 | A | G | 0.06 | -4.00E-02 | 8.99E-03 | 3.54E-05 | 17.11 |
| rs74543892 | 6 | 138050277 | T | G | 0.02 | 6.00E-02 | 1.50E-02 | 3.85E-05 | 16.94 |
| rs74774435 | 5 | 146438719 | T | C | 0.03 | 6.00E-02 | 1.26E-02 | 5.15E-06 | 20.78 |
| rs74787839 | 1 | 61565113 | C | A | 0.02 | 7.00E-02 | 1.56E-02 | 8.18E-06 | 19.89 |
| rs7493793 | 14 | 66299595 | G | A | 0.14 | -3.00E-02 | 6.28E-03 | 4.79E-05 | 16.53 |
| rs75177739 | 13 | 58846615 | C | T | 0.01 | -8.00E-02 | 1.90E-02 | 8.12E-06 | 19.91 |
| rs75454720 | 2 | 11189902 | C | G | 0.04 | -4.00E-02 | 1.09E-02 | 4.19E-05 | 16.79 |
| rs755683 | 8 | 18560687 | C | T | 0.22 | -3.00E-02 | 5.35E-03 | 1.16E-07 | 28.09 |
| rs7589627 | 2 | 27159786 | C | T | 0.58 | 2.00E-02 | 4.43E-03 | 4.69E-05 | 16.57 |
| rs7637333 | 3 | 185551152 | G | A | 0.03 | 5.00E-02 | 1.20E-02 | 2.04E-05 | 18.16 |
| rs76727778 | 15 | 44430599 | C | T | 0.08 | 3.00E-02 | 7.95E-03 | 1.42E-05 | 18.84 |
| rs77135115 | 7 | 47657502 | A | G | 0.05 | -4.00E-02 | 9.78E-03 | 6.53E-06 | 20.32 |
| rs77776018 | 1 | 42103083 | A | C | 0.09 | -3.00E-02 | 7.40E-03 | 2.46E-06 | 22.20 |
| rs7862308 | 9 | 11741707 | C | G | 0.06 | 4.00E-02 | 9.03E-03 | 2.22E-05 | 17.99 |
| rs78644671 | 3 | 14450997 | A | G | 0.02 | 7.00E-02 | 1.49E-02 | 5.63E-07 | 25.03 |
| rs79835808 | 3 | 192951 | G | T | 0.02 | -7.00E-02 | 1.71E-02 | 3.02E-05 | 17.41 |
| rs8069396 | 17 | 6287284 | G | T | 0.33 | -2.00E-02 | 4.70E-03 | 9.14E-06 | 19.69 |
| rs922439 | 5 | 43370366 | T | C | 0.2 | 2.00E-02 | 5.43E-03 | 3.48E-05 | 17.14 |
| rs9287892 | 2 | 168892318 | T | C | 0.84 | -3.00E-02 | 5.99E-03 | 5.56E-06 | 20.64 |
| rs9315421 | 13 | 37050799 | T | C | 0.04 | 5.00E-02 | 1.17E-02 | 3.00E-05 | 17.41 |
| rs9402037 | 6 | 128714425 | G | A | 0.23 | 2.00E-02 | 5.17E-03 | 1.64E-05 | 18.57 |
| rs9990594 | 4 | 113358286 | G | T | 0.48 | -8.00E-02 | 1.85E-02 | 9.73E-06 | 19.56 |

Abbreviations: SNP - single nucleotide polymorphism, EAF - effect allele frequency, SE - standard error.

Source genome-wide association study was Zhou et al.^23^

Multi-allelic instruments were included given that all datasets clearly report multiple alleles allowing comparison.

#### Table S5: Problematic alcohol use instruments, p<5x10-8, European ancestry.

| **SNPs** | **Chromosome** | **Position** | **Effect allele** | **Other allele** | **EAF** | **Beta** | **SE** | **P** | **F statistic** |
| --- | --- | --- | --- | --- | --- | --- | --- | --- | --- |
| rs10447987 | 8 | 113935783 | A | G | 0.97 | -4.00E-02 | 6.23E-03 | 7.61E-09 | 33.37 |
| rs10630359 | 14 | 58767085 | A | ATATACT | 0.29 | 1.00E-02 | 2.27E-03 | 1.91E-10 | 40.55 |
| rs10753661 | 1 | 165119792 | A | G | 0.69 | -1.00E-02 | 2.15E-03 | 3.73E-08 | 30.28 |
| rs10912807 | 1 | 174516366 | T | C | 0.54 | 1.00E-02 | 2.00E-03 | 2.11E-08 | 31.39 |
| rs11190658 | 10 | 102443374 | T | C | 0.22 | -1.00E-02 | 2.42E-03 | 1.41E-08 | 32.18 |
| rs11468270 | 20 | 37306245 | G | GTGGACTGATT | 0.69 | -1.00E-02 | 2.25E-03 | 3.61E-08 | 30.35 |
| rs1155397 | 7 | 114953597 | A | G | 0.32 | 1.00E-02 | 2.14E-03 | 7.94E-09 | 33.29 |
| rs12031120 | 1 | 73860821 | A | G | 0.60 | -1.00E-02 | 2.06E-03 | 1.96E-08 | 31.54 |
| rs12048727 | 1 | 71769465 | A | G | 0.66 | -1.00E-02 | 2.11E-03 | 1.73E-09 | 36.26 |
| rs12249880 | 10 | 110487071 | A | C | 0.75 | 2.00E-02 | 2.29E-03 | 1.04E-14 | 59.83 |
| rs1229984 | 4 | 100239319 | T | C | 0.03 | -1.90E-01 | 5.67E-03 | 4.14E-236 | 1076.56 |
| rs1260326 | 2 | 27730940 | T | C | 0.40 | -2.00E-02 | 2.04E-03 | 3.72E-26 | 111.92 |
| rs12826108 | 12 | 81603369 | A | G | 0.48 | -1.00E-02 | 2.00E-03 | 2.03E-09 | 35.95 |
| rs12885018 | 14 | 104273812 | A | G | 0.59 | -1.00E-02 | 2.03E-03 | 1.56E-09 | 36.46 |
| rs1291854 | 10 | 11110195 | T | C | 0.44 | -1.00E-02 | 2.01E-03 | 7.89E-11 | 42.29 |
| rs13024996 | 2 | 144225215 | A | C | 0.36 | -2.00E-02 | 2.07E-03 | 1.09E-13 | 55.20 |
| rs13102826 | 4 | 143602404 | A | G | 0.16 | -2.00E-02 | 2.72E-03 | 5.95E-12 | 47.35 |
| rs13107325 | 4 | 103188709 | T | C | 0.07 | -5.00E-02 | 3.83E-03 | 3.43E-43 | 189.86 |
| rs13149518 | 4 | 39406987 | T | C | 0.46 | -2.00E-02 | 2.01E-03 | 1.20E-27 | 118.72 |
| rs13276082 | 8 | 21823184 | A | G | 0.57 | 1.00E-02 | 2.02E-03 | 4.42E-08 | 29.95 |
| rs1327632 | 13 | 96848558 | T | C | 0.51 | -1.00E-02 | 2.00E-03 | 2.10E-08 | 31.39 |
| rs132902 | 22 | 41797758 | A | G | 0.78 | 1.00E-02 | 2.42E-03 | 2.29E-08 | 31.24 |
| rs1368742 | 3 | 85442208 | A | G | 0.39 | 2.00E-02 | 2.12E-03 | 4.94E-14 | 56.76 |
| rs142670988 | 8 | 143351879 | CAT | C | 0.38 | -1.00E-02 | 2.23E-03 | 8.18E-09 | 33.24 |
| rs1518394 | 2 | 58171287 | A | G | 0.40 | -1.00E-02 | 2.04E-03 | 1.03E-12 | 50.79 |
| rs16859408 | 3 | 147107665 | T | C | 0.85 | 2.00E-02 | 4.04E-03 | 2.56E-08 | 31.01 |
| rs1822717 | 8 | 64956228 | T | C | 0.48 | 1.00E-02 | 2.00E-03 | 1.81E-09 | 36.17 |
| rs192979726 | 10 | 110732490 | T | TAGAG | 0.17 | -2.00E-02 | 2.75E-03 | 9.12E-11 | 42.00 |
| rs200588406 | 11 | 57498460 | CTTAAA | C | 0.66 | -1.00E-02 | 2.19E-03 | 2.05E-09 | 35.92 |
| rs2022028 | 1 | 44606565 | A | G | 0.38 | -1.00E-02 | 2.06E-03 | 1.02E-10 | 41.78 |
| rs2098112 | 7 | 153487944 | A | G | 0.47 | 1.00E-02 | 2.07E-03 | 9.69E-10 | 37.38 |
| rs2159102 | 12 | 2288945 | A | T | 0.39 | 1.00E-02 | 2.06E-03 | 7.76E-09 | 33.34 |
| rs2245405 | 5 | 153364650 | A | G | 0.45 | -1.00E-02 | 2.04E-03 | 2.52E-08 | 31.05 |
| rs2375518 | 2 | 138270224 | A | G | 0.57 | -1.00E-02 | 2.02E-03 | 5.09E-10 | 38.64 |
| rs2576589 | 8 | 57425647 | T | C | 0.76 | -2.00E-02 | 2.32E-03 | 2.44E-13 | 53.61 |
| rs2664299 | 14 | 99742187 | T | C | 0.58 | 1.00E-02 | 2.03E-03 | 3.16E-09 | 35.08 |
| rs2696884 | 7 | 135144662 | T | C | 0.40 | 1.00E-02 | 2.04E-03 | 2.88E-13 | 53.29 |
| rs28562191 | 16 | 53799303 | T | C | 0.42 | -2.00E-02 | 2.09E-03 | 5.21E-15 | 61.18 |
| rs3217512 | 11 | 64135541 | G | GTC | 0.39 | -1.00E-02 | 2.08E-03 | 1.38E-08 | 32.22 |
| rs34338716 | 11 | 38989811 | CA | C | 0.33 | -1.00E-02 | 2.16E-03 | 6.07E-09 | 33.81 |
| rs34488670 | 15 | 47684936 | T | C | 0.79 | -2.00E-02 | 2.44E-03 | 2.04E-11 | 44.93 |
| rs34632468 | 11 | 113334227 | CT | C | 0.64 | 2.00E-02 | 2.11E-03 | 1.84E-24 | 104.18 |
| rs371206899 | 11 | 57522200 | CCCCT | C | 0.67 | -2.00E-02 | 2.84E-03 | 2.93E-11 | 44.22 |
| rs3953834 | 2 | 104343485 | A | T | 0.46 | -1.00E-02 | 2.11E-03 | 3.13E-08 | 30.63 |
| rs41301394 | 7 | 75612803 | T | C | 0.29 | -1.00E-02 | 2.21E-03 | 4.22E-08 | 30.04 |
| rs439523 | 19 | 49236102 | T | C | 0.49 | -1.00E-02 | 2.00E-03 | 3.83E-13 | 52.72 |
| rs4413606 | 6 | 26338257 | A | G | 0.36 | 1.00E-02 | 2.48E-03 | 2.49E-09 | 35.55 |
| rs460888 | 9 | 136913123 | C | G | 0.72 | -1.00E-02 | 2.23E-03 | 1.85E-08 | 31.65 |
| rs472140 | 2 | 45139904 | T | C | 0.58 | 2.00E-02 | 2.03E-03 | 2.26E-19 | 81.00 |
| rs551130066 | 4 | 39280428 | A | AT | 0.43 | 1.00E-02 | 2.05E-03 | 9.36E-09 | 32.97 |
| rs56009681 | 4 | 100285772 | A | T | 0.90 | -4.00E-02 | 3.29E-03 | 3.25E-41 | 180.79 |
| rs566835533 | 2 | 27667520 | T | TTTTG | 0.56 | -2.00E-02 | 2.09E-03 | 5.79E-17 | 70.06 |
| rs567678299 | 3 | 49088127 | A | AAAC | 0.75 | -1.00E-02 | 2.45E-03 | 4.39E-08 | 29.96 |
| rs572635656 | 16 | 53799278 | G | GT | 0.58 | 2.00E-02 | 2.09E-03 | 1.22E-14 | 59.51 |
| rs58976407 | 4 | 139853859 | T | C | 0.52 | 1.00E-02 | 2.00E-03 | 3.82E-08 | 30.24 |
| rs59899558 | 13 | 89072227 | T | G | 0.67 | 1.00E-02 | 2.24E-03 | 4.81E-10 | 38.75 |
| rs61904960 | 11 | 113481872 | T | C | 0.13 | 2.00E-02 | 2.92E-03 | 4.46E-13 | 52.43 |
| rs62061708 | 17 | 44006956 | A | G | 0.79 | 1.00E-02 | 2.44E-03 | 2.36E-08 | 31.17 |
| rs6265 | 11 | 27679916 | T | C | 0.19 | -1.00E-02 | 2.56E-03 | 2.50E-08 | 31.06 |
| rs6421482 | 1 | 66419905 | A | G | 0.44 | -1.00E-02 | 2.01E-03 | 1.39E-13 | 54.73 |
| rs6449591 | 5 | 61535659 | A | G | 0.58 | 1.00E-02 | 2.06E-03 | 2.21E-08 | 31.30 |
| rs6546862 | 2 | 73860348 | A | G | 0.64 | -1.00E-02 | 2.08E-03 | 1.41E-08 | 32.17 |
| rs66518420 | 3 | 49564316 | A | ATAT | 0.26 | 2.00E-02 | 2.32E-03 | 7.28E-13 | 51.47 |
| rs6841654 | 4 | 79570878 | A | G | 0.25 | 1.00E-02 | 2.33E-03 | 3.64E-09 | 34.81 |
| rs698988 | 3 | 157942860 | C | G | 0.57 | 1.00E-02 | 2.02E-03 | 1.94E-08 | 31.56 |
| rs7107356 | 11 | 47676170 | A | G | 0.50 | -2.00E-02 | 2.00E-03 | 1.20E-14 | 59.54 |
| rs71268403 | 3 | 16742862 | A | AT | 0.81 | -1.00E-02 | 2.61E-03 | 1.68E-08 | 31.83 |
| rs713287 | 4 | 46940376 | T | C | 0.48 | -1.00E-02 | 2.00E-03 | 1.55E-09 | 36.47 |
| rs72768664 | 16 | 24736518 | T | C | 0.94 | 2.00E-02 | 4.35E-03 | 4.16E-08 | 30.07 |
| rs7301932 | 12 | 51894913 | A | G | 0.47 | 1.00E-02 | 2.00E-03 | 5.40E-13 | 52.06 |
| rs73530179 | 16 | 30107781 | A | G | 0.53 | 1.00E-02 | 2.01E-03 | 5.63E-11 | 42.94 |
| rs7531138 | 1 | 97923499 | A | T | 0.44 | -1.00E-02 | 2.01E-03 | 8.30E-09 | 33.20 |
| rs7787380 | 7 | 71858167 | T | C | 0.48 | 1.00E-02 | 2.00E-03 | 2.06E-09 | 35.92 |
| rs7870475 | 9 | 128134034 | T | C | 0.52 | -1.00E-02 | 2.00E-03 | 5.96E-09 | 33.85 |
| rs7937768 | 11 | 121635061 | T | C | 0.77 | 2.00E-02 | 2.37E-03 | 1.29E-12 | 50.35 |
| rs8067440 | 17 | 29708155 | A | T | 0.61 | 1.00E-02 | 2.05E-03 | 6.02E-10 | 38.32 |
| rs825704 | 16 | 73603649 | T | C | 0.64 | 1.00E-02 | 2.08E-03 | 3.02E-08 | 30.70 |
| rs850254 | 14 | 57350427 | A | G | 0.56 | -1.00E-02 | 2.01E-03 | 1.49E-08 | 32.06 |
| rs854784 | 17 | 18040690 | T | C | 0.54 | 1.00E-02 | 2.00E-03 | 2.34E-08 | 31.19 |
| rs9372196 | 6 | 109132237 | A | G | 0.42 | -1.00E-02 | 2.02E-03 | 1.34E-08 | 32.27 |

Abbreviations: SNP - single nucleotide polymorphism, EAF - effect allele frequency, SE - standard error.

Source genome-wide association study was Zhou et al.^23^

Multi-allelic instruments were included given that all datasets clearly report multiple alleles allowing comparison.

#### Table S6: Drinks per week instruments, p<5x10^-8^, European ancestry.

| **SNP** | **Effect allele** | **Other allele** | **EAF** | **Beta** | **SE** | **P value** | **F statistic** |
| --- | --- | --- | --- | --- | --- | --- | --- |
| rs1000005 | C | G | 0.61 | -8.67E-03 | 2.00E-03 | 1.46E-05 | 18.79 |
| rs10009954 | T | A | 0.42 | -8.67E-03 | 2.00E-03 | 1.46E-05 | 18.79 |
| rs10027081 | T | C | 0.68 | -8.43E-03 | 2.00E-03 | 2.50E-05 | 17.77 |
| rs10044401 | G | A | 0.27 | 5.23E-03 | 2.00E-03 | 8.92E-03 | 6.84 |
| rs1004636 | C | G | 0.37 | -5.47E-03 | 2.00E-03 | 6.24E-03 | 7.48 |
| rs10065698 | A | C | 0.45 | -5.16E-03 | 2.00E-03 | 9.88E-03 | 6.66 |
| rs10091671 | A | G | 0.53 | 6.73E-03 | 2.00E-03 | 7.65E-04 | 11.32 |
| rs10113566 | C | T | 0.30 | -9.51E-03 | 2.00E-03 | 1.98E-06 | 22.61 |
| rs10137891 | A | G | 0.90 | 8.55E-03 | 3.00E-03 | 4.37E-03 | 8.12 |
| rs10155966 | G | A | 0.14 | -1.58E-02 | 3.00E-03 | 1.39E-07 | 27.74 |
| rs10161334 | T | C | 0.48 | -8.28E-03 | 2.00E-03 | 3.47E-05 | 17.14 |
| rs10170477 | A | C | 0.20 | -1.09E-03 | 2.00E-03 | 5.86E-01 | 0.30 |
| rs10173509 | T | A | 0.25 | 4.83E-03 | 2.00E-03 | 1.57E-02 | 5.83 |
| rs1018548 | A | G | 0.43 | 7.51E-03 | 2.00E-03 | 1.73E-04 | 14.10 |
| rs10198062 | T | A | 0.12 | -8.22E-03 | 2.00E-03 | 3.96E-05 | 16.89 |
| rs10208219 | A | G | 0.21 | 9.24E-03 | 2.00E-03 | 3.84E-06 | 21.34 |
| rs10208225 | A | G | 0.21 | 9.24E-03 | 2.00E-03 | 3.84E-06 | 21.34 |
| rs10249167 | G | A | 0.14 | -1.56E-02 | 3.00E-03 | 1.99E-07 | 27.04 |
| rs10277584 | C | T | 0.55 | 4.67E-03 | 2.00E-03 | 1.95E-02 | 5.45 |
| rs1043293 | C | G | 0.21 | 6.09E-03 | 2.00E-03 | 2.33E-03 | 9.27 |
| rs1048940 | T | C | 0.10 | -7.87E-03 | 3.00E-03 | 8.71E-03 | 6.88 |
| rs10491278 | C | A | 0.15 | -1.35E-02 | 2.00E-03 | 1.48E-11 | 45.56 |
| rs1050818 | C | A | 0.21 | 6.21E-03 | 2.00E-03 | 1.90E-03 | 9.64 |
| rs10513140 | C | T | 0.32 | 4.32E-03 | 2.00E-03 | 3.08E-02 | 4.67 |
| rs10514396 | C | T | 0.56 | 3.41E-03 | 2.00E-03 | 8.82E-02 | 2.91 |
| rs10518016 | T | A | 0.54 | 1.94E-04 | 2.00E-03 | 9.23E-01 | 0.01 |
| rs10757844 | T | C | 0.63 | 5.14E-03 | 2.00E-03 | 1.02E-02 | 6.60 |
| rs10760116 | T | C | 0.70 | -4.48E-03 | 2.00E-03 | 2.51E-02 | 5.02 |
| rs10785592 | A | G | 0.50 | -7.46E-03 | 2.00E-03 | 1.91E-04 | 13.91 |
| rs10788499 | T | C | 0.33 | -2.22E-03 | 2.00E-03 | 2.67E-01 | 1.23 |
| rs10788928 | A | T | 0.46 | -5.17E-03 | 2.00E-03 | 9.74E-03 | 6.68 |
| rs10790112 | C | T | 0.83 | 1.59E-02 | 2.00E-03 | 1.87E-15 | 63.20 |
| rs10792313 | C | G | 0.85 | -8.52E-03 | 3.00E-03 | 4.51E-03 | 8.07 |
| rs10797739 | T | C | 0.83 | 6.89E-03 | 2.00E-03 | 5.71E-04 | 11.87 |
| rs10812220 | C | T | 0.62 | 6.77E-03 | 2.00E-03 | 7.12E-04 | 11.46 |
| rs10819272 | C | T | 0.43 | -8.63E-03 | 2.00E-03 | 1.60E-05 | 18.62 |
| rs10821933 | C | T | 0.55 | -3.98E-03 | 2.00E-03 | 4.66E-02 | 3.96 |
| rs10861879 | A | G | 0.32 | -8.40E-03 | 2.00E-03 | 2.67E-05 | 17.64 |
| rs10865793 | C | G | 0.30 | 4.82E-03 | 2.00E-03 | 1.60E-02 | 5.81 |
| rs10873950 | G | A | 0.76 | -4.49E-03 | 2.00E-03 | 2.48E-02 | 5.04 |
| rs10878519 | T | C | 0.40 | 6.27E-03 | 2.00E-03 | 1.72E-03 | 9.83 |
| rs10884687 | T | A | 0.16 | -1.13E-02 | 2.00E-03 | 1.60E-08 | 31.92 |
| rs10891881 | T | C | 0.82 | 7.49E-03 | 2.00E-03 | 1.80E-04 | 14.03 |
| rs10912820 | T | C | 0.28 | -1.00E-02 | 2.00E-03 | 5.73E-07 | 25.00 |
| rs10912823 | G | A | 0.28 | -1.05E-02 | 2.00E-03 | 1.52E-07 | 27.56 |
| rs10934428 | G | A | 0.30 | -6.16E-03 | 2.00E-03 | 2.07E-03 | 9.49 |
| rs10938397 | G | A | 0.42 | -5.44E-03 | 2.00E-03 | 6.53E-03 | 7.40 |
| rs10968216 | A | G | 0.33 | 4.76E-03 | 2.00E-03 | 1.73E-02 | 5.66 |
| rs10975917 | C | G | 0.33 | -5.31E-03 | 2.00E-03 | 7.93E-03 | 7.05 |
| rs10978556 | C | A | 0.29 | -1.16E-02 | 2.00E-03 | 6.63E-09 | 33.64 |
| rs1101158 | A | T | 0.44 | 9.09E-03 | 2.00E-03 | 5.49E-06 | 20.66 |
| rs11021133 | A | G | 0.11 | 1.26E-02 | 3.00E-03 | 2.67E-05 | 17.64 |
| rs11021302 | A | G | 0.41 | -2.80E-03 | 2.00E-03 | 1.62E-01 | 1.96 |
| rs11030084 | T | C | 0.19 | -1.51E-02 | 2.00E-03 | 4.35E-14 | 57.00 |
| rs11030329 | G | A | 0.51 | 5.69E-03 | 2.00E-03 | 4.44E-03 | 8.09 |
| rs11031122 | C | T | 0.25 | -3.31E-03 | 2.00E-03 | 9.79E-02 | 2.74 |
| rs11039447 | A | G | 0.57 | -1.45E-02 | 2.00E-03 | 4.17E-13 | 52.56 |
| rs11039448 | T | G | 0.57 | -1.45E-02 | 2.00E-03 | 4.17E-13 | 52.56 |
| rs11043241 | G | A | 0.52 | -1.72E-03 | 2.00E-03 | 3.90E-01 | 0.74 |
| rs11055795 | A | G | 0.23 | 2.76E-03 | 2.00E-03 | 1.68E-01 | 1.90 |
| rs111226181 | T | G | 0.17 | -7.49E-03 | 2.00E-03 | 1.80E-04 | 14.03 |
| rs11130381 | T | C | 0.53 | -4.51E-03 | 2.00E-03 | 2.41E-02 | 5.09 |
| rs11156760 | C | G | 0.45 | 5.93E-03 | 2.00E-03 | 3.03E-03 | 8.79 |
| rs11158655 | T | C | 0.07 | -1.29E-02 | 4.00E-03 | 1.26E-03 | 10.40 |
| rs1116735 | T | G | 0.53 | 6.71E-03 | 2.00E-03 | 7.94E-04 | 11.26 |
| rs11170653 | T | C | 0.17 | -6.23E-03 | 2.00E-03 | 1.84E-03 | 9.70 |
| rs11183241 | T | C | 0.50 | -7.50E-03 | 2.00E-03 | 1.77E-04 | 14.06 |
| rs112010353 | T | C | 0.24 | -2.66E-02 | 2.00E-03 | 2.31E-40 | 176.89 |
| rs112154757 | G | T | 0.09 | 9.46E-03 | 3.00E-03 | 1.61E-03 | 9.94 |
| rs112389291 | T | C | 0.18 | -9.00E-03 | 2.00E-03 | 6.80E-06 | 20.25 |
| rs11240565 | T | C | 0.41 | -7.81E-03 | 2.00E-03 | 9.42E-05 | 15.25 |
| rs112586249 | G | T | 0.37 | -9.97E-03 | 2.00E-03 | 6.20E-07 | 24.85 |
| rs112952562 | A | C | 0.19 | 6.77E-03 | 2.00E-03 | 7.12E-04 | 11.46 |
| rs113242154 | TAG | T | 0.24 | -2.67E-02 | 2.00E-03 | 1.18E-40 | 178.22 |
| rs113519699 | C | A | 0.10 | -9.25E-03 | 3.00E-03 | 2.05E-03 | 9.51 |
| rs113744258 | G | T | 0.55 | -4.56E-03 | 2.00E-03 | 2.26E-02 | 5.20 |
| rs113825099 | A | G | 0.25 | -8.27E-03 | 2.00E-03 | 3.55E-05 | 17.10 |
| rs1143676 | A | G | 0.67 | -5.02E-03 | 2.00E-03 | 1.21E-02 | 6.30 |
| rs114934862 | G | A | 0.67 | 1.09E-02 | 3.00E-03 | 2.80E-04 | 13.20 |
| rs115083730 | A | G | 0.10 | 1.06E-02 | 3.00E-03 | 4.10E-04 | 12.48 |
| rs11547301 | T | C | 0.24 | -8.70E-03 | 2.00E-03 | 1.36E-05 | 18.92 |
| rs11590198 | A | G | 0.18 | -9.35E-03 | 2.00E-03 | 2.94E-06 | 21.86 |
| rs11590410 | T | C | 0.18 | 9.39E-03 | 2.00E-03 | 2.67E-06 | 22.04 |
| rs11628894 | A | C | 0.68 | 4.74E-03 | 2.00E-03 | 1.78E-02 | 5.62 |
| rs116332912 | C | T | 0.06 | -5.80E-03 | 3.00E-03 | 5.32E-02 | 3.74 |
| rs11642015 | T | C | 0.43 | -1.16E-02 | 2.00E-03 | 6.63E-09 | 33.64 |
| rs11648483 | C | T | 0.30 | 1.02E-02 | 2.00E-03 | 3.40E-07 | 26.01 |
| rs11657366 | G | A | 0.40 | 7.42E-03 | 2.00E-03 | 2.07E-04 | 13.76 |
| rs11690748 | G | C | 0.35 | -6.00E-03 | 2.00E-03 | 2.70E-03 | 9.00 |
| rs11692435 | A | G | 0.08 | 1.91E-02 | 3.00E-03 | 1.93E-10 | 40.53 |
| rs1169286 | C | T | 0.44 | -3.22E-03 | 2.00E-03 | 1.07E-01 | 2.59 |
| rs11693852 | G | T | 0.80 | -1.13E-02 | 2.00E-03 | 1.60E-08 | 31.92 |
| rs11709092 | C | A | 0.10 | -1.07E-02 | 3.00E-03 | 3.62E-04 | 12.72 |
| rs11710277 | G | A | 0.08 | -1.10E-02 | 3.00E-03 | 2.46E-04 | 13.44 |
| rs11713946 | C | G | 0.30 | 4.91E-03 | 2.00E-03 | 1.41E-02 | 6.03 |
| rs11714337 | A | G | 0.44 | 7.40E-03 | 2.00E-03 | 2.16E-04 | 13.69 |
| rs11716614 | G | C | 0.10 | -1.02E-02 | 3.00E-03 | 6.74E-04 | 11.56 |
| rs117634579 | T | A | 0.28 | -6.50E-03 | 2.00E-03 | 1.15E-03 | 10.56 |
| rs11764779 | G | A | 0.20 | 9.91E-03 | 3.00E-03 | 9.55E-04 | 10.91 |
| rs11784243 | G | C | 0.19 | -3.42E-03 | 2.00E-03 | 8.73E-02 | 2.92 |
| rs117907741 | G | A | 0.08 | 7.84E-03 | 4.00E-03 | 5.00E-02 | 3.84 |
| rs11808967 | G | T | 0.27 | 4.03E-03 | 2.00E-03 | 4.39E-02 | 4.06 |
| rs11835638 | C | G | 0.46 | -1.16E-02 | 2.00E-03 | 6.63E-09 | 33.64 |
| rs11860773 | C | T | 0.18 | -1.57E-02 | 2.00E-03 | 4.16E-15 | 61.62 |
| rs11873164 | T | C | 0.13 | -1.15E-02 | 2.00E-03 | 8.92E-09 | 33.06 |
| rs11899999 | G | A | 0.57 | 7.96E-03 | 2.00E-03 | 6.89E-05 | 15.84 |
| rs11922956 | G | A | 0.62 | -1.58E-02 | 2.00E-03 | 2.79E-15 | 62.41 |
| rs1193509 | C | T | 0.44 | 9.16E-03 | 2.00E-03 | 4.65E-06 | 20.98 |
| rs1193510 | G | A | 0.44 | 9.16E-03 | 2.00E-03 | 4.65E-06 | 20.98 |
| rs11940694 | G | A | 0.61 | 2.79E-02 | 2.00E-03 | 3.15E-44 | 194.60 |
| rs11950819 | C | T | 0.35 | 5.73E-03 | 2.00E-03 | 4.17E-03 | 8.21 |
| rs11956421 | C | T | 0.44 | 7.74E-03 | 2.00E-03 | 1.09E-04 | 14.98 |
| rs11967609 | A | T | 0.22 | -8.10E-03 | 2.00E-03 | 5.12E-05 | 16.40 |
| rs11973460 | G | T | 0.14 | -1.61E-02 | 3.00E-03 | 8.02E-08 | 28.80 |
| rs12032052 | G | A | 0.13 | -6.79E-03 | 2.00E-03 | 6.86E-04 | 11.53 |
| rs12039810 | A | G | 0.33 | -9.68E-03 | 2.00E-03 | 1.30E-06 | 23.43 |
| rs12053569 | A | G | 0.10 | -9.76E-03 | 3.00E-03 | 1.14E-03 | 10.58 |
| rs12062170 | G | C | 0.28 | -1.04E-02 | 2.00E-03 | 1.99E-07 | 27.04 |
| rs12086693 | G | A | 0.28 | -1.05E-02 | 2.00E-03 | 1.52E-07 | 27.56 |
| rs12089565 | T | C | 0.20 | 5.74E-03 | 2.00E-03 | 4.10E-03 | 8.24 |
| rs12127789 | T | G | 0.11 | -1.30E-02 | 3.00E-03 | 1.47E-05 | 18.78 |
| rs12141218 | C | T | 0.21 | 6.09E-03 | 2.00E-03 | 2.33E-03 | 9.27 |
| rs12147929 | G | A | 0.37 | -3.95E-03 | 2.00E-03 | 4.83E-02 | 3.90 |
| rs12175393 | C | A | 0.26 | -5.54E-03 | 2.00E-03 | 5.61E-03 | 7.67 |
| rs12202969 | A | G | 0.51 | 9.62E-03 | 2.00E-03 | 1.51E-06 | 23.14 |
| rs12212034 | T | C | 0.36 | -8.82E-03 | 2.00E-03 | 1.03E-05 | 19.45 |
| rs12249740 | G | A | 0.16 | -1.17E-02 | 2.00E-03 | 4.92E-09 | 34.22 |
| rs1229984 | C | T | 0.97 | 1.93E-01 | 5.00E-03 | 0.00E+00 | 1489.96 |
| rs12314162 | T | C | 0.16 | 8.22E-03 | 2.00E-03 | 3.96E-05 | 16.89 |
| rs12412636 | T | C | 0.81 | -6.04E-03 | 2.00E-03 | 2.53E-03 | 9.12 |
| rs12446550 | A | G | 0.35 | -1.54E-02 | 2.00E-03 | 1.36E-14 | 59.29 |
| rs12457578 | T | C | 0.09 | -9.84E-03 | 3.00E-03 | 1.04E-03 | 10.76 |
| rs12458976 | G | A | 0.09 | -9.98E-03 | 3.00E-03 | 8.79E-04 | 11.07 |
| rs12462756 | A | G | 0.17 | -5.42E-03 | 2.00E-03 | 6.73E-03 | 7.34 |
| rs12463914 | T | C | 0.20 | 8.62E-03 | 2.00E-03 | 1.63E-05 | 18.58 |
| rs12465809 | C | T | 0.32 | 4.65E-03 | 2.00E-03 | 2.01E-02 | 5.41 |
| rs12472516 | C | T | 0.28 | 4.11E-03 | 2.00E-03 | 3.99E-02 | 4.22 |
| rs1248634 | A | G | 0.28 | 7.30E-03 | 2.00E-03 | 2.62E-04 | 13.32 |
| rs1248636 | A | C | 0.28 | 7.25E-03 | 2.00E-03 | 2.89E-04 | 13.14 |
| rs12487273 | G | T | 0.26 | -8.49E-03 | 2.00E-03 | 2.19E-05 | 18.02 |
| rs12507026 | T | A | 0.42 | -5.38E-03 | 2.00E-03 | 7.15E-03 | 7.24 |
| rs12513758 | G | C | 0.15 | -1.41E-02 | 3.00E-03 | 2.60E-06 | 22.09 |
| rs12566724 | C | T | 0.27 | -9.44E-03 | 2.00E-03 | 2.36E-06 | 22.28 |
| rs1260326 | C | T | 0.59 | 2.54E-02 | 2.00E-03 | 5.91E-37 | 161.29 |
| rs12620188 | G | A | 0.21 | 8.37E-03 | 3.00E-03 | 5.27E-03 | 7.78 |
| rs12641030 | G | A | 0.47 | -6.27E-03 | 2.00E-03 | 1.72E-03 | 9.83 |
| rs12651605 | C | A | 0.42 | -8.92E-03 | 2.00E-03 | 8.20E-06 | 19.89 |
| rs12658601 | A | G | 0.27 | -8.70E-03 | 2.00E-03 | 1.36E-05 | 18.92 |
| rs1267059 | C | A | 0.77 | 9.67E-03 | 2.00E-03 | 1.33E-06 | 23.38 |
| rs12696822 | A | G | 0.53 | 6.75E-03 | 2.00E-03 | 7.38E-04 | 11.39 |
| rs12698826 | G | T | 0.23 | -1.34E-02 | 2.00E-03 | 2.08E-11 | 44.89 |
| rs12702099 | C | T | 0.83 | 2.24E-03 | 3.00E-03 | 4.55E-01 | 0.56 |
| rs12739982 | G | C | 0.15 | 5.51E-03 | 2.00E-03 | 5.87E-03 | 7.59 |
| rs12739983 | A | C | 0.15 | 5.51E-03 | 2.00E-03 | 5.87E-03 | 7.59 |
| rs12743163 | A | G | 0.14 | -4.41E-03 | 3.00E-03 | 1.42E-01 | 2.16 |
| rs12778721 | T | A | 0.15 | -1.09E-02 | 3.00E-03 | 2.80E-04 | 13.20 |
| rs1285989 | T | C | 0.57 | -7.50E-03 | 2.00E-03 | 1.77E-04 | 14.06 |
| rs12896307 | C | T | 0.24 | -9.26E-03 | 2.00E-03 | 3.66E-06 | 21.44 |
| rs12901675 | G | A | 0.44 | 9.18E-03 | 2.00E-03 | 4.43E-06 | 21.07 |
| rs12904384 | G | T | 0.46 | -3.10E-03 | 2.00E-03 | 1.21E-01 | 2.40 |
| rs12924622 | T | A | 0.46 | -9.88E-03 | 2.00E-03 | 7.81E-07 | 24.40 |
| rs12924631 | A | G | 0.30 | 1.01E-02 | 2.00E-03 | 4.42E-07 | 25.50 |
| rs12941981 | A | C | 0.10 | -1.36E-02 | 3.00E-03 | 5.81E-06 | 20.55 |
| rs12986332 | C | G | 0.33 | 5.12E-03 | 2.00E-03 | 1.05E-02 | 6.55 |
| rs12991555 | T | A | 0.36 | -1.26E-02 | 2.00E-03 | 2.98E-10 | 39.69 |
| rs12998288 | T | C | 0.50 | 6.29E-03 | 2.00E-03 | 1.66E-03 | 9.89 |
| rs13024996 | A | C | 0.36 | -1.36E-02 | 2.00E-03 | 1.05E-11 | 46.24 |
| rs13056230 | T | C | 0.42 | -6.84E-03 | 2.00E-03 | 6.26E-04 | 11.70 |
| rs13107325 | T | C | 0.08 | -3.93E-02 | 4.00E-03 | 8.79E-23 | 96.53 |
| rs13147317 | A | G | 0.80 | 8.47E-03 | 2.00E-03 | 2.29E-05 | 17.94 |
| rs13158011 | C | G | 0.54 | -8.60E-03 | 2.00E-03 | 1.71E-05 | 18.49 |
| rs13165059 | T | C | 0.54 | -8.37E-03 | 2.00E-03 | 2.85E-05 | 17.51 |
| rs13173892 | T | G | 0.54 | -8.48E-03 | 2.00E-03 | 2.24E-05 | 17.98 |
| rs1318102 | T | A | 0.43 | 6.43E-03 | 2.00E-03 | 1.30E-03 | 10.34 |
| rs13208885 | G | A | 0.06 | 1.02E-02 | 3.00E-03 | 6.74E-04 | 11.56 |
| rs13259172 | G | T | 0.24 | 5.32E-03 | 2.00E-03 | 7.81E-03 | 7.08 |
| rs13273763 | A | T | 0.03 | -1.26E-02 | 4.00E-03 | 1.63E-03 | 9.92 |
| rs1329067 | A | G | 0.14 | 8.66E-03 | 3.00E-03 | 3.89E-03 | 8.33 |
| rs1331342 | T | C | 0.59 | -7.52E-03 | 2.00E-03 | 1.70E-04 | 14.14 |
| rs13389219 | T | C | 0.44 | 4.45E-03 | 2.00E-03 | 2.61E-02 | 4.95 |
| rs13425752 | C | T | 0.67 | 5.09E-03 | 2.00E-03 | 1.09E-02 | 6.48 |
| rs1346369 | G | A | 0.10 | 9.58E-03 | 3.00E-03 | 1.41E-03 | 10.20 |
| rs1350917 | A | G | 0.44 | -4.50E-03 | 2.00E-03 | 2.44E-02 | 5.06 |
| rs1353284 | T | C | 0.37 | 4.00E-03 | 2.00E-03 | 4.55E-02 | 4.00 |
| rs1363970 | C | T | 0.18 | 9.64E-03 | 2.00E-03 | 1.44E-06 | 23.23 |
| rs1377491 | T | A | 0.82 | 1.29E-02 | 2.00E-03 | 1.12E-10 | 41.60 |
| rs1379981 | C | T | 0.70 | 5.05E-03 | 2.00E-03 | 1.16E-02 | 6.38 |
| rs1381577 | C | T | 0.47 | -5.73E-03 | 2.00E-03 | 4.17E-03 | 8.21 |
| rs138459260 | A | G | 0.05 | -1.51E-02 | 4.00E-03 | 1.60E-04 | 14.25 |
| rs1387766 | A | G | 0.62 | -1.06E-02 | 2.00E-03 | 1.16E-07 | 28.09 |
| rs139532049 | G | A | 0.37 | 2.88E-03 | 2.00E-03 | 1.50E-01 | 2.07 |
| rs140204792 | A | G | 0.09 | 9.49E-03 | 3.00E-03 | 1.56E-03 | 10.01 |
| rs1421085 | C | T | 0.43 | -1.16E-02 | 2.00E-03 | 6.63E-09 | 33.64 |
| rs1452152 | G | A | 0.35 | 7.08E-03 | 2.00E-03 | 4.00E-04 | 12.53 |
| rs1453376 | C | T | 0.82 | 1.29E-02 | 2.00E-03 | 1.12E-10 | 41.60 |
| rs145830500 | T | C | 0.15 | 8.82E-03 | 3.00E-03 | 3.28E-03 | 8.64 |
| rs1471712 | A | G | 0.57 | -1.45E-02 | 2.00E-03 | 4.17E-13 | 52.56 |
| rs150306 | C | T | 0.44 | -5.00E-03 | 2.00E-03 | 1.24E-02 | 6.25 |
| rs1506719 | C | T | 0.50 | -7.68E-03 | 2.00E-03 | 1.23E-04 | 14.75 |
| rs1512133 | C | T | 0.53 | 6.76E-03 | 2.00E-03 | 7.25E-04 | 11.42 |
| rs1512134 | T | G | 0.53 | 6.76E-03 | 2.00E-03 | 7.25E-04 | 11.42 |
| rs1512183 | T | A | 0.26 | 5.89E-03 | 2.00E-03 | 3.23E-03 | 8.67 |
| rs152189 | C | T | 0.27 | -8.70E-03 | 2.00E-03 | 1.36E-05 | 18.92 |
| rs1524974 | G | T | 0.37 | 2.81E-03 | 2.00E-03 | 1.60E-01 | 1.97 |
| rs1534756 | G | C | 0.55 | 4.61E-03 | 2.00E-03 | 2.12E-02 | 5.31 |
| rs1558902 | A | T | 0.43 | -1.17E-02 | 2.00E-03 | 4.92E-09 | 34.22 |
| rs1568254 | A | C | 0.59 | 1.10E-02 | 2.00E-03 | 3.80E-08 | 30.25 |
| rs159531 | G | A | 0.75 | 8.39E-03 | 2.00E-03 | 2.73E-05 | 17.60 |
| rs1652340 | C | A | 0.65 | 9.11E-03 | 2.00E-03 | 5.24E-06 | 20.75 |
| rs1654885 | G | A | 0.93 | 3.75E-03 | 3.00E-03 | 2.11E-01 | 1.56 |
| rs1666658 | C | T | 0.41 | -5.57E-03 | 2.00E-03 | 5.35E-03 | 7.76 |
| rs1671499 | A | G | 0.61 | -5.12E-03 | 2.00E-03 | 1.05E-02 | 6.55 |
| rs16862488 | T | C | 0.15 | -8.51E-03 | 2.00E-03 | 2.09E-05 | 18.11 |
| rs16882167 | A | G | 0.52 | 6.59E-03 | 2.00E-03 | 9.84E-04 | 10.86 |
| rs17016768 | T | C | 0.30 | -1.19E-02 | 2.00E-03 | 2.68E-09 | 35.40 |
| rs17026944 | A | G | 0.26 | 5.95E-03 | 2.00E-03 | 2.93E-03 | 8.85 |
| rs17087472 | G | A | 0.09 | 1.00E-02 | 3.00E-03 | 8.58E-04 | 11.11 |
| rs17177078 | T | C | 0.07 | -2.63E-02 | 4.00E-03 | 4.87E-11 | 43.23 |
| rs1728922 | C | A | 0.37 | 1.24E-02 | 2.00E-03 | 5.65E-10 | 38.44 |
| rs17394997 | C | T | 0.05 | 4.65E-03 | 4.00E-03 | 2.45E-01 | 1.35 |
| rs17478715 | G | C | 0.09 | -9.44E-03 | 3.00E-03 | 1.65E-03 | 9.90 |
| rs17550821 | A | G | 0.16 | -4.63E-03 | 2.00E-03 | 2.06E-02 | 5.36 |
| rs17621058 | G | A | 0.32 | 8.25E-03 | 2.00E-03 | 3.71E-05 | 17.02 |
| rs17648701 | C | T | 0.19 | -7.20E-03 | 2.00E-03 | 3.18E-04 | 12.96 |
| rs17665139 | T | C | 0.15 | -9.74E-03 | 2.00E-03 | 1.12E-06 | 23.72 |
| rs17702278 | G | C | 0.07 | 9.85E-03 | 3.00E-03 | 1.03E-03 | 10.78 |
| rs17742929 | A | G | 0.25 | -8.21E-03 | 2.00E-03 | 4.04E-05 | 16.85 |
| rs17771664 | A | G | 0.26 | 8.13E-03 | 2.00E-03 | 4.80E-05 | 16.52 |
| rs17836088 | C | G | 0.21 | -8.35E-03 | 2.00E-03 | 2.98E-05 | 17.43 |
| rs1783835 | G | A | 0.55 | 1.02E-02 | 2.00E-03 | 3.40E-07 | 26.01 |
| rs17842490 | G | A | 0.01 | 3.34E-02 | 7.00E-03 | 1.83E-06 | 22.77 |
| rs1837305 | A | G | 0.09 | 1.01E-02 | 3.00E-03 | 7.61E-04 | 11.33 |
| rs186046560 | G | C | 0.15 | -1.58E-02 | 3.00E-03 | 1.39E-07 | 27.74 |
| rs1865250 | C | T | 0.62 | -1.59E-02 | 2.00E-03 | 1.87E-15 | 63.20 |
| rs1884937 | T | A | 0.22 | -8.05E-03 | 2.00E-03 | 5.70E-05 | 16.20 |
| rs1887813 | T | C | 0.57 | -4.93E-03 | 2.00E-03 | 1.37E-02 | 6.08 |
| rs1895615 | C | T | 0.74 | 6.35E-03 | 2.00E-03 | 1.50E-03 | 10.08 |
| rs191237460 | C | T | 0.04 | 1.73E-02 | 4.00E-03 | 1.53E-05 | 18.71 |
| rs192246131 | A | G | 0.10 | -1.24E-02 | 4.00E-03 | 1.94E-03 | 9.61 |
| rs1940728 | T | G | 0.43 | 9.11E-03 | 2.00E-03 | 5.24E-06 | 20.75 |
| rs1942964 | G | T | 0.50 | -1.07E-02 | 2.00E-03 | 8.80E-08 | 28.62 |
| rs1944688 | A | G | 0.76 | 1.09E-02 | 2.00E-03 | 5.04E-08 | 29.70 |
| rs194849 | A | G | 0.50 | 1.14E-02 | 2.00E-03 | 1.20E-08 | 32.49 |
| rs194865 | A | G | 0.50 | 1.14E-02 | 2.00E-03 | 1.20E-08 | 32.49 |
| rs194868 | C | T | 0.50 | 1.15E-02 | 2.00E-03 | 8.92E-09 | 33.06 |
| rs1949640 | A | G | 0.24 | -9.84E-03 | 2.00E-03 | 8.65E-07 | 24.21 |
| rs1953184 | A | G | 0.60 | -8.82E-03 | 2.00E-03 | 1.03E-05 | 19.45 |
| rs1975611 | T | C | 0.22 | 3.77E-03 | 2.00E-03 | 5.94E-02 | 3.55 |
| rs1990635 | G | A | 0.25 | 9.82E-03 | 2.00E-03 | 9.11E-07 | 24.11 |
| rs1994190 | T | C | 0.50 | 6.54E-03 | 2.00E-03 | 1.08E-03 | 10.69 |
| rs2001846 | C | T | 0.52 | 1.00E-02 | 2.00E-03 | 5.73E-07 | 25.00 |
| rs2005616 | T | G | 0.62 | -8.44E-03 | 2.00E-03 | 2.44E-05 | 17.81 |
| rs201589 | T | C | 0.55 | -8.84E-03 | 2.00E-03 | 9.87E-06 | 19.54 |
| rs201797375 | T | TCTC | 0.03 | -3.71E-02 | 5.00E-03 | 1.17E-13 | 55.06 |
| rs202667 | A | G | 0.78 | 1.28E-02 | 2.00E-03 | 1.55E-10 | 40.96 |
| rs203162 | C | T | 0.48 | 5.99E-03 | 2.00E-03 | 2.74E-03 | 8.97 |
| rs203172 | T | C | 0.48 | 6.01E-03 | 2.00E-03 | 2.66E-03 | 9.03 |
| rs2037853 | C | A | 0.74 | -8.80E-03 | 2.00E-03 | 1.08E-05 | 19.36 |
| rs2061406 | A | G | 0.21 | -7.86E-03 | 2.00E-03 | 8.49E-05 | 15.44 |
| rs2070489 | G | A | 0.62 | 8.89E-03 | 2.00E-03 | 8.79E-06 | 19.76 |
| rs2086824 | C | A | 0.47 | -4.12E-03 | 2.00E-03 | 3.94E-02 | 4.24 |
| rs2098112 | A | G | 0.46 | 1.27E-02 | 2.00E-03 | 2.15E-10 | 40.32 |
| rs2098628 | C | T | 0.60 | -4.91E-03 | 2.00E-03 | 1.41E-02 | 6.03 |
| rs210596 | C | A | 0.32 | -1.09E-02 | 2.00E-03 | 5.04E-08 | 29.70 |
| rs210600 | A | G | 0.26 | -1.33E-02 | 2.00E-03 | 2.93E-11 | 44.22 |
| rs2106993 | G | A | 0.37 | -8.90E-03 | 2.00E-03 | 8.59E-06 | 19.80 |
| rs2108421 | T | A | 0.55 | 6.83E-03 | 2.00E-03 | 6.38E-04 | 11.66 |
| rs2132489 | T | C | 0.57 | 6.64E-03 | 2.00E-03 | 9.00E-04 | 11.02 |
| rs2132661 | G | A | 0.21 | 1.04E-02 | 2.00E-03 | 1.99E-07 | 27.04 |
| rs2142205 | A | C | 0.54 | 3.13E-03 | 2.00E-03 | 1.18E-01 | 2.45 |
| rs2159102 | A | T | 0.38 | 8.87E-03 | 2.00E-03 | 9.21E-06 | 19.67 |
| rs217322 | T | C | 0.44 | 6.36E-03 | 2.00E-03 | 1.47E-03 | 10.11 |
| rs2186799 | T | G | 0.53 | -9.55E-03 | 2.00E-03 | 1.80E-06 | 22.80 |
| rs2187927 | G | A | 0.13 | -6.75E-03 | 3.00E-03 | 2.44E-02 | 5.06 |
| rs2200159 | C | G | 0.57 | -6.69E-03 | 2.00E-03 | 8.23E-04 | 11.19 |
| rs2223708 | C | T | 0.41 | -5.37E-03 | 2.00E-03 | 7.25E-03 | 7.21 |
| rs2229677 | G | A | 0.64 | 3.77E-03 | 2.00E-03 | 5.94E-02 | 3.55 |
| rs2240310 | T | C | 0.45 | -4.71E-03 | 2.00E-03 | 1.85E-02 | 5.55 |
| rs2250522 | A | G | 0.61 | -3.43E-03 | 2.00E-03 | 8.63E-02 | 2.94 |
| rs2268890 | G | A | 0.61 | -6.49E-03 | 2.00E-03 | 1.17E-03 | 10.53 |
| rs2279829 | T | C | 0.21 | -8.90E-03 | 2.00E-03 | 8.59E-06 | 19.80 |
| rs2298214 | A | C | 0.59 | -6.89E-03 | 2.00E-03 | 5.71E-04 | 11.87 |
| rs231046 | A | G | 0.17 | -1.06E-02 | 2.00E-03 | 1.16E-07 | 28.09 |
| rs2315927 | T | C | 0.24 | -9.39E-03 | 2.00E-03 | 2.67E-06 | 22.04 |
| rs2316901 | A | G | 0.37 | 9.41E-03 | 2.00E-03 | 2.54E-06 | 22.14 |
| rs2322440 | T | G | 0.34 | 5.84E-03 | 2.00E-03 | 3.50E-03 | 8.53 |
| rs2381760 | G | A | 0.18 | -9.00E-03 | 2.00E-03 | 6.80E-06 | 20.25 |
| rs2394120 | C | A | 0.22 | -8.11E-03 | 2.00E-03 | 5.01E-05 | 16.44 |
| rs2394121 | G | T | 0.22 | -8.04E-03 | 2.00E-03 | 5.82E-05 | 16.16 |
| rs2412973 | A | C | 0.45 | 7.10E-03 | 2.00E-03 | 3.85E-04 | 12.60 |
| rs2452170 | A | G | 0.47 | 1.52E-02 | 2.00E-03 | 2.96E-14 | 57.76 |
| rs247226 | A | G | 0.27 | -8.59E-03 | 2.00E-03 | 1.75E-05 | 18.45 |
| rs247271 | A | T | 0.27 | -8.71E-03 | 2.00E-03 | 1.33E-05 | 18.97 |
| rs247272 | A | C | 0.27 | -8.70E-03 | 2.00E-03 | 1.36E-05 | 18.92 |
| rs2514218 | T | C | 0.35 | -1.42E-02 | 2.00E-03 | 1.25E-12 | 50.41 |
| rs2525569 | A | T | 0.62 | 9.24E-03 | 2.00E-03 | 3.84E-06 | 21.34 |
| rs2587507 | C | T | 0.50 | -5.01E-03 | 2.00E-03 | 1.22E-02 | 6.28 |
| rs2599510 | G | A | 0.51 | -6.41E-03 | 2.00E-03 | 1.35E-03 | 10.27 |
| rs2619364 | G | A | 0.30 | 4.03E-03 | 2.00E-03 | 4.39E-02 | 4.06 |
| rs2622586 | C | T | 0.36 | 3.64E-03 | 2.00E-03 | 6.88E-02 | 3.31 |
| rs2638282 | A | G | 0.47 | 1.52E-02 | 2.00E-03 | 2.96E-14 | 57.76 |
| rs26561 | G | A | 0.37 | -7.36E-03 | 2.00E-03 | 2.33E-04 | 13.54 |
| rs2682406 | A | T | 0.44 | 9.16E-03 | 2.00E-03 | 4.65E-06 | 20.98 |
| rs2685799 | C | T | 0.49 | 1.76E-03 | 2.00E-03 | 3.79E-01 | 0.77 |
| rs2693672 | G | C | 0.41 | -7.28E-03 | 2.00E-03 | 2.73E-04 | 13.25 |
| rs2698322 | C | G | 0.44 | 9.07E-03 | 2.00E-03 | 5.76E-06 | 20.57 |
| rs2703488 | C | T | 0.49 | 6.22E-03 | 2.00E-03 | 1.87E-03 | 9.67 |
| rs271072 | C | A | 0.63 | -3.99E-03 | 2.00E-03 | 4.60E-02 | 3.98 |
| rs2713552 | T | G | 0.61 | -8.95E-03 | 2.00E-03 | 7.64E-06 | 20.03 |
| rs2713556 | T | C | 0.61 | -9.23E-03 | 2.00E-03 | 3.93E-06 | 21.30 |
| rs2731458 | T | C | 0.43 | 4.54E-03 | 2.00E-03 | 2.32E-02 | 5.15 |
| rs27535 | C | T | 0.38 | -7.31E-03 | 2.00E-03 | 2.57E-04 | 13.36 |
| rs276683 | A | G | 0.74 | 4.90E-03 | 2.00E-03 | 1.43E-02 | 6.00 |
| rs2783129 | G | C | 0.50 | -4.91E-03 | 2.00E-03 | 1.41E-02 | 6.03 |
| rs2817865 | A | G | 0.42 | 8.88E-03 | 2.00E-03 | 9.00E-06 | 19.71 |
| rs2817866 | A | G | 0.42 | 8.89E-03 | 2.00E-03 | 8.79E-06 | 19.76 |
| rs2839402 | T | C | 0.20 | 9.71E-03 | 2.00E-03 | 1.20E-06 | 23.57 |
| rs28522500 | C | T | 0.11 | -7.18E-03 | 3.00E-03 | 1.67E-02 | 5.73 |
| rs28569001 | T | C | 0.12 | -1.34E-02 | 3.00E-03 | 7.94E-06 | 19.95 |
| rs2866372 | T | C | 0.46 | -2.74E-03 | 2.00E-03 | 1.71E-01 | 1.88 |
| rs28712821 | A | G | 0.61 | 2.78E-02 | 2.00E-03 | 6.33E-44 | 193.21 |
| rs28929474 | T | C | 0.02 | -4.89E-02 | 6.00E-03 | 3.64E-16 | 66.42 |
| rs2970982 | T | C | 0.15 | -5.60E-03 | 2.00E-03 | 5.11E-03 | 7.84 |
| rs2992070 | T | C | 0.26 | -8.27E-03 | 2.00E-03 | 3.55E-05 | 17.10 |
| rs30242 | C | T | 0.41 | 2.20E-03 | 2.00E-03 | 2.71E-01 | 1.21 |
| rs3110474 | T | G | 0.71 | -4.95E-03 | 2.00E-03 | 1.33E-02 | 6.13 |
| rs3123142 | C | T | 0.79 | -6.36E-03 | 2.00E-03 | 1.47E-03 | 10.11 |
| rs3124420 | T | G | 0.75 | -6.56E-03 | 2.00E-03 | 1.04E-03 | 10.76 |
| rs3190836 | T | A | 0.27 | 5.46E-03 | 2.00E-03 | 6.33E-03 | 7.45 |
| rs3214499 | GA | G | 0.41 | 1.89E-02 | 2.00E-03 | 3.39E-21 | 89.30 |
| rs324012 | T | C | 0.41 | 7.93E-03 | 2.00E-03 | 7.34E-05 | 15.72 |
| rs338708 | C | T | 0.68 | 7.02E-03 | 2.00E-03 | 4.48E-04 | 12.32 |
| rs339046 | C | T | 0.49 | -8.50E-03 | 2.00E-03 | 2.14E-05 | 18.06 |
| rs339900 | A | G | 0.55 | 2.92E-03 | 2.00E-03 | 1.44E-01 | 2.13 |
| rs34037363 | T | C | 0.11 | -1.05E-02 | 3.00E-03 | 4.65E-04 | 12.25 |
| rs34188744 | A | G | 0.64 | -3.06E-04 | 2.00E-03 | 8.78E-01 | 0.02 |
| rs34440469 | A | G | 0.28 | -8.32E-03 | 2.00E-03 | 3.18E-05 | 17.31 |
| rs34489326 | G | A | 0.26 | -5.01E-03 | 2.00E-03 | 1.22E-02 | 6.28 |
| rs34566402 | T | C | 0.15 | -9.02E-03 | 3.00E-03 | 2.64E-03 | 9.04 |
| rs34664008 | C | A | 0.15 | -5.33E-03 | 2.00E-03 | 7.70E-03 | 7.10 |
| rs34704785 | T | C | 0.52 | -1.02E-02 | 2.00E-03 | 3.40E-07 | 26.01 |
| rs34794623 | A | C | 0.21 | 1.05E-02 | 2.00E-03 | 1.52E-07 | 27.56 |
| rs34908147 | A | G | 0.28 | 5.35E-03 | 2.00E-03 | 7.47E-03 | 7.16 |
| rs35058767 | A | G | 0.38 | -8.30E-03 | 2.00E-03 | 3.32E-05 | 17.22 |
| rs35175834 | A | G | 0.21 | 1.05E-02 | 2.00E-03 | 1.52E-07 | 27.56 |
| rs35400171 | G | T | 0.59 | 7.37E-03 | 2.00E-03 | 2.29E-04 | 13.58 |
| rs35473814 | A | G | 0.21 | 1.04E-02 | 2.00E-03 | 1.99E-07 | 27.04 |
| rs35534970 | A | G | 0.21 | 1.01E-02 | 2.00E-03 | 4.42E-07 | 25.50 |
| rs35890289 | T | G | 0.27 | -6.39E-03 | 2.00E-03 | 1.40E-03 | 10.21 |
| rs35972889 | G | A | 0.12 | -1.36E-02 | 3.00E-03 | 5.81E-06 | 20.55 |
| rs362307 | T | C | 0.06 | -8.90E-03 | 3.00E-03 | 3.01E-03 | 8.80 |
| rs3758396 | T | C | 0.88 | 1.11E-02 | 3.00E-03 | 2.16E-04 | 13.69 |
| rs3759575 | G | A | 0.23 | 3.98E-03 | 2.00E-03 | 4.66E-02 | 3.96 |
| rs3761422 | C | T | 0.62 | 7.37E-03 | 2.00E-03 | 2.29E-04 | 13.58 |
| rs3770381 | C | A | 0.39 | -2.55E-03 | 2.00E-03 | 2.02E-01 | 1.63 |
| rs3790908 | A | G | 0.13 | -6.64E-03 | 2.00E-03 | 9.00E-04 | 11.02 |
| rs379410 | C | T | 0.09 | -1.75E-02 | 3.00E-03 | 5.43E-09 | 34.03 |
| rs3798461 | A | G | 0.26 | -5.02E-03 | 2.00E-03 | 1.21E-02 | 6.30 |
| rs3849743 | G | A | 0.44 | -2.90E-03 | 2.00E-03 | 1.47E-01 | 2.10 |
| rs3857984 | G | A | 0.41 | -7.07E-03 | 2.00E-03 | 4.08E-04 | 12.50 |
| rs3901955 | G | A | 0.12 | -9.35E-03 | 3.00E-03 | 1.83E-03 | 9.71 |
| rs3911276 | A | G | 0.39 | -8.55E-03 | 2.00E-03 | 1.91E-05 | 18.28 |
| rs400288 | C | A | 0.24 | 6.87E-03 | 2.00E-03 | 5.93E-04 | 11.80 |
| rs411354 | C | T | 0.57 | -7.28E-03 | 2.00E-03 | 2.73E-04 | 13.25 |
| rs4273169 | A | G | 0.36 | -1.34E-02 | 2.00E-03 | 2.08E-11 | 44.89 |
| rs4291495 | T | C | 0.42 | -7.85E-03 | 2.00E-03 | 8.67E-05 | 15.41 |
| rs4329921 | G | A | 0.25 | 9.80E-03 | 2.00E-03 | 9.58E-07 | 24.01 |
| rs4378971 | T | A | 0.79 | -6.70E-03 | 2.00E-03 | 8.08E-04 | 11.22 |
| rs4416687 | T | G | 0.39 | -8.38E-03 | 2.00E-03 | 2.79E-05 | 17.56 |
| rs4425611 | T | C | 0.39 | -8.40E-03 | 2.00E-03 | 2.67E-05 | 17.64 |
| rs4438553 | T | C | 0.41 | -5.48E-03 | 2.00E-03 | 6.14E-03 | 7.51 |
| rs4493211 | T | G | 0.21 | 8.18E-03 | 2.00E-03 | 4.31E-05 | 16.73 |
| rs4508098 | C | T | 0.24 | 7.62E-03 | 2.00E-03 | 1.39E-04 | 14.52 |
| rs4542420 | C | G | 0.24 | 7.22E-03 | 2.00E-03 | 3.06E-04 | 13.03 |
| rs4568472 | T | G | 0.39 | -8.41E-03 | 2.00E-03 | 2.61E-05 | 17.68 |
| rs4657405 | C | T | 0.74 | -1.11E-02 | 2.00E-03 | 2.86E-08 | 30.80 |
| rs4659382 | G | C | 0.27 | -8.31E-03 | 2.00E-03 | 3.25E-05 | 17.26 |
| rs4665962 | G | A | 0.20 | 1.30E-02 | 2.00E-03 | 8.03E-11 | 42.25 |
| rs4682718 | G | A | 0.18 | -7.39E-03 | 2.00E-03 | 2.20E-04 | 13.65 |
| rs4698921 | T | C | 0.58 | 1.00E-04 | 2.00E-03 | 9.60E-01 | 0.00 |
| rs4735044 | C | T | 0.24 | -1.16E-02 | 2.00E-03 | 6.63E-09 | 33.64 |
| rs4762557 | C | T | 0.58 | 6.16E-03 | 2.00E-03 | 2.07E-03 | 9.49 |
| rs4786999 | G | C | 0.48 | -7.13E-03 | 2.00E-03 | 3.64E-04 | 12.71 |
| rs4788084 | T | C | 0.35 | -1.54E-02 | 2.00E-03 | 1.36E-14 | 59.29 |
| rs4810972 | A | G | 0.56 | -4.81E-03 | 2.00E-03 | 1.62E-02 | 5.78 |
| rs4821815 | A | G | 0.35 | -5.36E-03 | 2.00E-03 | 7.36E-03 | 7.18 |
| rs4833416 | A | G | 0.67 | -7.23E-03 | 2.00E-03 | 3.00E-04 | 13.07 |
| rs4861023 | G | A | 0.23 | 1.13E-02 | 2.00E-03 | 1.60E-08 | 31.92 |
| rs4861024 | G | A | 0.23 | 1.15E-02 | 2.00E-03 | 8.92E-09 | 33.06 |
| rs4879286 | T | C | 0.76 | -6.35E-03 | 2.00E-03 | 1.50E-03 | 10.08 |
| rs4879287 | G | A | 0.76 | -6.37E-03 | 2.00E-03 | 1.45E-03 | 10.14 |
| rs4879465 | A | G | 0.15 | 6.93E-03 | 3.00E-03 | 2.09E-02 | 5.34 |
| rs4912532 | T | G | 0.53 | 8.85E-03 | 2.00E-03 | 9.64E-06 | 19.58 |
| rs4946198 | T | C | 0.73 | -5.32E-03 | 2.00E-03 | 7.81E-03 | 7.08 |
| rs4984900 | A | G | 0.25 | 6.48E-03 | 2.00E-03 | 1.20E-03 | 10.50 |
| rs500321 | T | A | 0.72 | -1.02E-02 | 2.00E-03 | 3.40E-07 | 26.01 |
| rs504764 | T | C | 0.34 | 1.60E-02 | 2.00E-03 | 1.24E-15 | 64.00 |
| rs518969 | C | A | 0.26 | -5.51E-03 | 2.00E-03 | 5.87E-03 | 7.59 |
| rs534840 | T | A | 0.45 | 5.70E-03 | 2.00E-03 | 4.37E-03 | 8.12 |
| rs541455835 | T | TA | 0.24 | -2.47E-02 | 2.00E-03 | 4.87E-35 | 152.52 |
| rs549284860 | C | T | 0.21 | 1.33E-02 | 3.00E-03 | 9.28E-06 | 19.65 |
| rs55677435 | G | T | 0.15 | -9.38E-03 | 3.00E-03 | 1.77E-03 | 9.78 |
| rs55722796 | T | C | 0.75 | 8.18E-03 | 2.00E-03 | 4.31E-05 | 16.73 |
| rs55755441 | T | C | 0.17 | 6.50E-03 | 2.00E-03 | 1.15E-03 | 10.56 |
| rs55833463 | T | G | 0.18 | 9.77E-03 | 2.00E-03 | 1.03E-06 | 23.86 |
| rs55918644 | G | A | 0.52 | 6.72E-03 | 2.00E-03 | 7.79E-04 | 11.29 |
| rs55962391 | C | T | 0.14 | -8.38E-03 | 3.00E-03 | 5.22E-03 | 7.80 |
| rs56095217 | A | G | 0.27 | -8.69E-03 | 2.00E-03 | 1.39E-05 | 18.88 |
| rs56172794 | T | C | 0.53 | 8.83E-03 | 2.00E-03 | 1.01E-05 | 19.49 |
| rs56226746 | A | G | 0.32 | -1.40E-02 | 2.00E-03 | 2.56E-12 | 49.00 |
| rs56300626 | C | T | 0.09 | 1.00E-02 | 3.00E-03 | 8.58E-04 | 11.11 |
| rs56915106 | A | G | 0.19 | -1.13E-02 | 2.00E-03 | 1.60E-08 | 31.92 |
| rs57010886 | G | T | 0.12 | -7.04E-03 | 3.00E-03 | 1.89E-02 | 5.51 |
| rs57024418 | G | A | 0.32 | 8.27E-03 | 2.00E-03 | 3.55E-05 | 17.10 |
| rs57281063 | A | G | 0.39 | 1.00E-02 | 2.00E-03 | 5.73E-07 | 25.00 |
| rs57563458 | A | G | 0.42 | -7.77E-03 | 2.00E-03 | 1.02E-04 | 15.09 |
| rs57835221 | A | G | 0.27 | -8.75E-03 | 2.00E-03 | 1.21E-05 | 19.14 |
| rs5794864 | A | AT | 0.61 | -1.28E-02 | 2.00E-03 | 1.55E-10 | 40.96 |
| rs58150027 | T | G | 0.32 | -4.00E-03 | 2.00E-03 | 4.55E-02 | 4.00 |
| rs58189451 | A | G | 0.35 | 4.94E-03 | 2.00E-03 | 1.35E-02 | 6.10 |
| rs584768 | A | G | 0.47 | 1.53E-02 | 2.00E-03 | 2.01E-14 | 58.52 |
| rs589030 | C | G | 0.37 | -5.04E-03 | 2.00E-03 | 1.17E-02 | 6.35 |
| rs59478157 | G | A | 0.03 | -1.27E-02 | 5.00E-03 | 1.11E-02 | 6.45 |
| rs596364 | C | T | 0.75 | -6.50E-03 | 2.00E-03 | 1.15E-03 | 10.56 |
| rs597069 | T | G | 0.16 | -1.07E-02 | 3.00E-03 | 3.62E-04 | 12.72 |
| rs6001620 | T | C | 0.28 | 3.73E-03 | 2.00E-03 | 6.22E-02 | 3.48 |
| rs60232290 | C | T | 0.38 | 3.66E-03 | 2.00E-03 | 6.72E-02 | 3.35 |
| rs6059729 | G | A | 0.10 | 1.11E-02 | 3.00E-03 | 2.16E-04 | 13.69 |
| rs60632986 | T | G | 0.46 | 4.83E-03 | 2.00E-03 | 1.57E-02 | 5.83 |
| rs6068280 | G | A | 0.66 | -3.34E-03 | 2.00E-03 | 9.49E-02 | 2.79 |
| rs6130096 | T | C | 0.35 | 7.07E-03 | 2.00E-03 | 4.08E-04 | 12.50 |
| rs61408162 | T | C | 0.32 | -6.75E-03 | 2.00E-03 | 7.38E-04 | 11.39 |
| rs61792373 | G | C | 0.52 | 6.60E-03 | 2.00E-03 | 9.67E-04 | 10.89 |
| rs619920 | G | C | 0.26 | -4.93E-03 | 2.00E-03 | 1.37E-02 | 6.08 |
| rs62015178 | A | G | 0.01 | 1.92E-02 | 6.00E-03 | 1.37E-03 | 10.24 |
| rs62034321 | T | C | 0.35 | -1.54E-02 | 2.00E-03 | 1.36E-14 | 59.29 |
| rs62034323 | T | C | 0.35 | -1.54E-02 | 2.00E-03 | 1.36E-14 | 59.29 |
| rs62041531 | G | A | 0.23 | -1.13E-02 | 2.00E-03 | 1.60E-08 | 31.92 |
| rs62048402 | A | G | 0.43 | -1.16E-02 | 2.00E-03 | 6.63E-09 | 33.64 |
| rs62064660 | A | G | 0.24 | -2.35E-02 | 2.00E-03 | 7.06E-32 | 138.06 |
| rs62100772 | A | G | 0.42 | -6.74E-03 | 2.00E-03 | 7.52E-04 | 11.36 |
| rs62100776 | T | A | 0.45 | -6.51E-03 | 2.00E-03 | 1.13E-03 | 10.60 |
| rs62107261 | C | T | 0.04 | -1.81E-02 | 4.00E-03 | 6.04E-06 | 20.48 |
| rs62143863 | T | C | 0.34 | -3.21E-03 | 2.00E-03 | 1.08E-01 | 2.58 |
| rs62171925 | G | T | 0.32 | 5.20E-03 | 2.00E-03 | 9.32E-03 | 6.76 |
| rs62244884 | A | G | 0.44 | 7.64E-03 | 2.00E-03 | 1.33E-04 | 14.59 |
| rs62325468 | C | G | 0.04 | -3.16E-02 | 5.00E-03 | 2.62E-10 | 39.94 |
| rs62366921 | A | G | 0.19 | -7.92E-03 | 2.00E-03 | 7.49E-05 | 15.68 |
| rs62369151 | C | T | 0.39 | -9.43E-03 | 2.00E-03 | 2.42E-06 | 22.23 |
| rs62388807 | T | C | 0.50 | -9.92E-03 | 2.00E-03 | 7.05E-07 | 24.60 |
| rs62403684 | G | A | 0.33 | 9.84E-03 | 2.00E-03 | 8.65E-07 | 24.21 |
| rs6449597 | A | G | 0.27 | -8.74E-03 | 2.00E-03 | 1.24E-05 | 19.10 |
| rs6482725 | G | C | 0.13 | -9.81E-03 | 3.00E-03 | 1.08E-03 | 10.69 |
| rs6485794 | G | C | 0.43 | 1.45E-02 | 2.00E-03 | 4.17E-13 | 52.56 |
| rs6493274 | C | A | 0.21 | 1.04E-02 | 2.00E-03 | 1.99E-07 | 27.04 |
| rs6498447 | C | T | 0.65 | -7.17E-03 | 2.00E-03 | 3.37E-04 | 12.85 |
| rs6502619 | G | C | 0.56 | 7.97E-03 | 2.00E-03 | 6.75E-05 | 15.88 |
| rs6599691 | G | T | 0.16 | -8.84E-03 | 2.00E-03 | 9.87E-06 | 19.54 |
| rs66539338 | T | C | 0.16 | -1.15E-02 | 2.00E-03 | 8.92E-09 | 33.06 |
| rs6664662 | C | T | 0.34 | 4.23E-03 | 2.00E-03 | 3.44E-02 | 4.47 |
| rs66720517 | T | C | 0.15 | -6.80E-03 | 2.00E-03 | 6.74E-04 | 11.56 |
| rs6679878 | G | A | 0.72 | -3.73E-03 | 2.00E-03 | 6.22E-02 | 3.48 |
| rs6681970 | G | A | 0.28 | -1.05E-02 | 2.00E-03 | 1.52E-07 | 27.56 |
| rs6683030 | T | C | 0.50 | 4.37E-03 | 2.00E-03 | 2.89E-02 | 4.77 |
| rs6696267 | C | A | 0.56 | -5.32E-03 | 2.00E-03 | 7.81E-03 | 7.08 |
| rs6696784 | A | G | 0.60 | 5.18E-03 | 2.00E-03 | 9.60E-03 | 6.71 |
| rs6700838 | T | C | 0.61 | 4.47E-03 | 2.00E-03 | 2.54E-02 | 5.00 |
| rs6723680 | G | A | 0.25 | 4.85E-03 | 2.00E-03 | 1.53E-02 | 5.88 |
| rs672643 | G | C | 0.31 | -3.41E-03 | 2.00E-03 | 8.82E-02 | 2.91 |
| rs6739804 | C | T | 0.64 | -1.36E-02 | 2.00E-03 | 1.05E-11 | 46.24 |
| rs676388 | C | T | 0.47 | 1.54E-02 | 2.00E-03 | 1.36E-14 | 59.29 |
| rs6770091 | A | G | 0.46 | 4.98E-03 | 2.00E-03 | 1.28E-02 | 6.20 |
| rs6776912 | G | A | 0.21 | -4.78E-03 | 2.00E-03 | 1.68E-02 | 5.71 |
| rs6785354 | T | C | 0.76 | 4.06E-03 | 2.00E-03 | 4.24E-02 | 4.12 |
| rs67994637 | A | G | 0.25 | 9.83E-03 | 2.00E-03 | 8.88E-07 | 24.16 |
| rs6802087 | T | C | 0.53 | 8.81E-03 | 2.00E-03 | 1.06E-05 | 19.40 |
| rs6813691 | T | C | 0.15 | -5.83E-03 | 2.00E-03 | 3.56E-03 | 8.50 |
| rs6817004 | T | C | 0.75 | -3.33E-03 | 2.00E-03 | 9.59E-02 | 2.77 |
| rs6821766 | C | G | 0.23 | 1.13E-02 | 2.00E-03 | 1.60E-08 | 31.92 |
| rs6828213 | G | T | 0.85 | 9.32E-03 | 2.00E-03 | 3.16E-06 | 21.72 |
| rs6828896 | C | A | 0.28 | -1.02E-02 | 2.00E-03 | 3.40E-07 | 26.01 |
| rs6830501 | A | G | 0.47 | -6.20E-03 | 2.00E-03 | 1.94E-03 | 9.61 |
| rs6830774 | C | T | 0.47 | -6.23E-03 | 2.00E-03 | 1.84E-03 | 9.70 |
| rs6843300 | C | T | 0.80 | 8.50E-03 | 2.00E-03 | 2.14E-05 | 18.06 |
| rs6861634 | C | T | 0.44 | -5.26E-03 | 2.00E-03 | 8.54E-03 | 6.92 |
| rs690893 | A | C | 0.66 | -6.36E-03 | 2.00E-03 | 1.47E-03 | 10.11 |
| rs6921970 | T | C | 0.26 | -4.96E-03 | 2.00E-03 | 1.31E-02 | 6.15 |
| rs6924111 | A | T | 0.13 | -3.50E-03 | 2.00E-03 | 8.01E-02 | 3.06 |
| rs6941096 | C | A | 0.39 | -8.55E-03 | 2.00E-03 | 1.91E-05 | 18.28 |
| rs6941722 | A | G | 0.39 | -8.58E-03 | 2.00E-03 | 1.79E-05 | 18.40 |
| rs6963882 | G | C | 0.29 | -1.01E-02 | 2.00E-03 | 4.42E-07 | 25.50 |
| rs6977921 | G | A | 0.43 | 9.52E-03 | 2.00E-03 | 1.94E-06 | 22.66 |
| rs6984305 | T | A | 0.91 | 4.93E-03 | 3.00E-03 | 1.00E-01 | 2.70 |
| rs7004498 | A | G | 0.20 | 4.47E-03 | 2.00E-03 | 2.54E-02 | 5.00 |
| rs7019416 | A | G | 0.53 | 6.18E-03 | 2.00E-03 | 2.00E-03 | 9.55 |
| rs7035059 | C | G | 0.34 | -2.94E-03 | 2.00E-03 | 1.42E-01 | 2.16 |
| rs7044210 | C | A | 0.12 | -9.15E-03 | 3.00E-03 | 2.29E-03 | 9.30 |
| rs704491 | C | T | 0.09 | -7.69E-03 | 3.00E-03 | 1.04E-02 | 6.57 |
| rs7073419 | T | C | 0.16 | -1.16E-02 | 2.00E-03 | 6.63E-09 | 33.64 |
| rs7101911 | T | G | 0.06 | 2.01E-02 | 3.00E-03 | 2.08E-11 | 44.89 |
| rs7121038 | T | C | 0.57 | -1.45E-02 | 2.00E-03 | 4.17E-13 | 52.56 |
| rs7131926 | A | T | 0.47 | 6.68E-03 | 2.00E-03 | 8.38E-04 | 11.16 |
| rs7132908 | A | G | 0.36 | -9.34E-03 | 2.00E-03 | 3.01E-06 | 21.81 |
| rs713598 | G | C | 0.42 | -4.67E-03 | 2.00E-03 | 1.95E-02 | 5.45 |
| rs71432275 | G | GAC | 0.64 | -1.28E-02 | 2.00E-03 | 1.55E-10 | 40.96 |
| rs71438074 | C | T | 0.08 | -1.42E-02 | 4.00E-03 | 3.85E-04 | 12.60 |
| rs71541330 | A | C | 0.18 | -7.14E-03 | 2.00E-03 | 3.57E-04 | 12.74 |
| rs71573392 | T | G | 0.15 | -9.24E-03 | 3.00E-03 | 2.07E-03 | 9.49 |
| rs7164309 | G | A | 0.36 | -9.71E-03 | 2.00E-03 | 1.20E-06 | 23.57 |
| rs7166534 | C | T | 0.21 | 1.03E-02 | 2.00E-03 | 2.60E-07 | 26.52 |
| rs7175414 | A | G | 0.21 | 1.03E-02 | 2.00E-03 | 2.60E-07 | 26.52 |
| rs7192896 | A | C | 0.25 | 9.85E-03 | 2.00E-03 | 8.44E-07 | 24.26 |
| rs7193557 | G | A | 0.25 | 9.85E-03 | 2.00E-03 | 8.44E-07 | 24.26 |
| rs721862 | A | G | 0.47 | 8.84E-03 | 2.00E-03 | 9.87E-06 | 19.54 |
| rs7237124 | C | T | 0.07 | -7.29E-03 | 3.00E-03 | 1.51E-02 | 5.90 |
| rs7239712 | G | A | 0.35 | -8.76E-03 | 2.00E-03 | 1.19E-05 | 19.18 |
| rs7243008 | T | A | 0.45 | -6.51E-03 | 2.00E-03 | 1.13E-03 | 10.60 |
| rs7244046 | A | G | 0.36 | 7.18E-03 | 2.00E-03 | 3.31E-04 | 12.89 |
| rs7251466 | C | G | 0.10 | 1.02E-02 | 3.00E-03 | 6.74E-04 | 11.56 |
| rs726433 | T | C | 0.09 | 1.01E-02 | 3.00E-03 | 7.61E-04 | 11.33 |
| rs72715245 | T | C | 0.24 | -8.97E-03 | 2.00E-03 | 7.29E-06 | 20.12 |
| rs72733137 | C | A | 0.18 | 9.46E-03 | 2.00E-03 | 2.25E-06 | 22.37 |
| rs72737614 | G | A | 0.09 | 9.45E-03 | 3.00E-03 | 1.63E-03 | 9.92 |
| rs72737615 | A | G | 0.09 | 9.45E-03 | 3.00E-03 | 1.63E-03 | 9.92 |
| rs72737616 | T | C | 0.09 | 9.46E-03 | 3.00E-03 | 1.61E-03 | 9.94 |
| rs72768626 | G | A | 0.07 | -2.58E-02 | 4.00E-03 | 1.12E-10 | 41.60 |
| rs72829908 | G | A | 0.16 | -1.12E-02 | 2.00E-03 | 2.14E-08 | 31.36 |
| rs72843198 | T | C | 0.04 | -1.36E-02 | 4.00E-03 | 6.74E-04 | 11.56 |
| rs72856551 | T | G | 0.24 | -9.33E-03 | 2.00E-03 | 3.09E-06 | 21.76 |
| rs72859280 | T | G | 0.05 | 2.34E-02 | 5.00E-03 | 2.87E-06 | 21.90 |
| rs72891133 | T | C | 0.16 | 6.65E-03 | 2.00E-03 | 8.84E-04 | 11.06 |
| rs72898831 | G | A | 0.13 | -1.15E-02 | 2.00E-03 | 8.92E-09 | 33.06 |
| rs72991092 | C | G | 0.39 | -8.57E-03 | 2.00E-03 | 1.83E-05 | 18.36 |
| rs7306606 | T | C | 0.55 | 2.93E-03 | 2.00E-03 | 1.43E-01 | 2.15 |
| rs73068342 | G | A | 0.28 | -5.76E-03 | 2.00E-03 | 3.98E-03 | 8.29 |
| rs7307912 | T | C | 0.25 | -1.07E-02 | 2.00E-03 | 8.80E-08 | 28.62 |
| rs73080312 | T | C | 0.10 | -7.91E-03 | 3.00E-03 | 8.37E-03 | 6.95 |
| rs73115953 | T | G | 0.19 | 4.53E-03 | 2.00E-03 | 2.35E-02 | 5.13 |
| rs73139125 | G | A | 0.26 | 5.98E-03 | 2.00E-03 | 2.79E-03 | 8.94 |
| rs7314011 | T | G | 0.59 | 6.01E-03 | 2.00E-03 | 2.66E-03 | 9.03 |
| rs73193904 | C | G | 0.14 | -6.06E-03 | 2.00E-03 | 2.45E-03 | 9.18 |
| rs732770 | A | G | 0.34 | 9.56E-03 | 2.00E-03 | 1.75E-06 | 22.85 |
| rs7332053 | T | G | 0.17 | -3.55E-03 | 2.00E-03 | 7.59E-02 | 3.15 |
| rs7447792 | C | T | 0.73 | 7.83E-03 | 2.00E-03 | 9.04E-05 | 15.33 |
| rs7468983 | T | C | 0.09 | 1.01E-02 | 3.00E-03 | 7.61E-04 | 11.33 |
| rs7503334 | T | C | 0.56 | 7.96E-03 | 2.00E-03 | 6.89E-05 | 15.84 |
| rs7503866 | G | A | 0.40 | 7.43E-03 | 2.00E-03 | 2.03E-04 | 13.80 |
| rs75214378 | A | G | 0.03 | 1.21E-02 | 5.00E-03 | 1.55E-02 | 5.86 |
| rs7539883 | A | G | 0.74 | -1.12E-02 | 2.00E-03 | 2.14E-08 | 31.36 |
| rs7542102 | C | T | 0.27 | 2.59E-04 | 2.00E-03 | 8.97E-01 | 0.02 |
| rs7542510 | T | C | 0.21 | 6.12E-03 | 2.00E-03 | 2.21E-03 | 9.36 |
| rs7562032 | A | G | 0.44 | 7.21E-03 | 2.00E-03 | 3.12E-04 | 13.00 |
| rs7575544 | C | T | 0.27 | 7.40E-03 | 2.00E-03 | 2.16E-04 | 13.69 |
| rs7580759 | T | C | 0.38 | 1.23E-02 | 2.00E-03 | 7.75E-10 | 37.82 |
| rs7581524 | T | C | 0.26 | 1.04E-02 | 2.00E-03 | 1.99E-07 | 27.04 |
| rs7584354 | T | C | 0.40 | 6.54E-03 | 2.00E-03 | 1.08E-03 | 10.69 |
| rs7584386 | G | C | 0.40 | 6.47E-03 | 2.00E-03 | 1.22E-03 | 10.47 |
| rs76104618 | A | G | 0.08 | -8.95E-03 | 3.00E-03 | 2.85E-03 | 8.90 |
| rs76192127 | C | T | 0.13 | 2.61E-03 | 2.00E-03 | 1.92E-01 | 1.70 |
| rs76202302 | T | C | 0.04 | 7.62E-03 | 4.00E-03 | 5.68E-02 | 3.63 |
| rs76476582 | T | C | 0.04 | -1.58E-02 | 4.00E-03 | 7.82E-05 | 15.60 |
| rs7656573 | A | T | 0.52 | 6.56E-03 | 2.00E-03 | 1.04E-03 | 10.76 |
| rs7657201 | A | T | 0.52 | 6.60E-03 | 2.00E-03 | 9.67E-04 | 10.89 |
| rs76701 | T | C | 0.31 | 7.76E-03 | 2.00E-03 | 1.04E-04 | 15.05 |
| rs7678535 | G | A | 0.52 | 6.59E-03 | 2.00E-03 | 9.84E-04 | 10.86 |
| rs76822519 | A | C | 0.05 | 7.43E-03 | 4.00E-03 | 6.32E-02 | 3.45 |
| rs7690809 | G | A | 0.70 | 5.09E-03 | 2.00E-03 | 1.09E-02 | 6.48 |
| rs7703288 | G | C | 0.35 | 5.88E-03 | 2.00E-03 | 3.28E-03 | 8.64 |
| rs77131596 | C | T | 0.06 | 2.07E-02 | 3.00E-03 | 5.20E-12 | 47.61 |
| rs7720784 | C | G | 0.39 | 5.98E-03 | 2.00E-03 | 2.79E-03 | 8.94 |
| rs77232328 | T | C | 0.05 | 2.26E-02 | 4.00E-03 | 1.60E-08 | 31.92 |
| rs7762246 | C | T | 0.16 | 6.63E-03 | 2.00E-03 | 9.16E-04 | 10.99 |
| rs7824565 | G | A | 0.21 | -9.08E-03 | 2.00E-03 | 5.63E-06 | 20.61 |
| rs7838301 | G | A | 0.15 | 3.45E-03 | 2.00E-03 | 8.45E-02 | 2.98 |
| rs7842666 | A | G | 0.53 | 6.83E-03 | 2.00E-03 | 6.38E-04 | 11.66 |
| rs7857323 | T | A | 0.31 | -7.07E-03 | 2.00E-03 | 4.08E-04 | 12.50 |
| rs7857936 | C | T | 0.37 | 2.21E-03 | 2.00E-03 | 2.69E-01 | 1.22 |
| rs78815801 | C | T | 0.11 | 1.58E-02 | 3.00E-03 | 1.39E-07 | 27.74 |
| rs78822708 | C | G | 0.47 | 4.14E-03 | 2.00E-03 | 3.85E-02 | 4.28 |
| rs79273563 | T | C | 0.05 | -1.28E-02 | 4.00E-03 | 1.37E-03 | 10.24 |
| rs7933896 | T | C | 0.43 | 1.55E-02 | 2.00E-03 | 9.19E-15 | 60.06 |
| rs7950528 | C | T | 0.37 | 9.45E-03 | 2.00E-03 | 2.30E-06 | 22.33 |
| rs79541554 | T | C | 0.08 | 8.00E-03 | 3.00E-03 | 7.66E-03 | 7.11 |
| rs7961349 | T | A | 0.50 | 5.13E-03 | 2.00E-03 | 1.03E-02 | 6.58 |
| rs7962117 | G | A | 0.15 | -1.97E-03 | 2.00E-03 | 3.25E-01 | 0.97 |
| rs799165 | A | T | 0.12 | 1.56E-02 | 3.00E-03 | 1.99E-07 | 27.04 |
| rs80127733 | G | T | 0.06 | 1.98E-02 | 3.00E-03 | 4.11E-11 | 43.56 |
| rs8025614 | A | G | 0.54 | -4.84E-03 | 2.00E-03 | 1.55E-02 | 5.86 |
| rs8032333 | G | A | 0.21 | 1.05E-02 | 2.00E-03 | 1.52E-07 | 27.56 |
| rs8040191 | T | C | 0.21 | 1.03E-02 | 2.00E-03 | 2.60E-07 | 26.52 |
| rs8047349 | G | C | 0.71 | 9.23E-03 | 2.00E-03 | 3.93E-06 | 21.30 |
| rs8065563 | A | G | 0.56 | 7.97E-03 | 2.00E-03 | 6.75E-05 | 15.88 |
| rs8100343 | T | C | 0.21 | 3.40E-03 | 2.00E-03 | 8.91E-02 | 2.89 |
| rs8114499 | C | T | 0.15 | 9.68E-03 | 3.00E-03 | 1.25E-03 | 10.41 |
| rs833820 | C | A | 0.43 | -4.22E-03 | 2.00E-03 | 3.49E-02 | 4.45 |
| rs8519 | G | A | 0.66 | 7.99E-03 | 2.00E-03 | 6.47E-05 | 15.96 |
| rs874758 | G | A | 0.81 | -5.53E-03 | 2.00E-03 | 5.69E-03 | 7.65 |
| rs888071 | C | A | 0.25 | -5.03E-03 | 2.00E-03 | 1.19E-02 | 6.33 |
| rs8891 | C | T | 0.45 | 7.37E-03 | 2.00E-03 | 2.29E-04 | 13.58 |
| rs919853 | C | T | 0.25 | 9.23E-03 | 2.00E-03 | 3.93E-06 | 21.30 |
| rs9330453 | A | G | 0.46 | -5.38E-03 | 2.00E-03 | 7.15E-03 | 7.24 |
| rs9343314 | A | G | 0.26 | -4.94E-03 | 2.00E-03 | 1.35E-02 | 6.10 |
| rs9343317 | A | G | 0.26 | -4.96E-03 | 2.00E-03 | 1.31E-02 | 6.15 |
| rs9343318 | G | A | 0.26 | -5.00E-03 | 2.00E-03 | 1.24E-02 | 6.25 |
| rs9349702 | C | T | 0.65 | -4.87E-03 | 2.00E-03 | 1.49E-02 | 5.93 |
| rs9365750 | A | G | 0.24 | -5.92E-03 | 2.00E-03 | 3.08E-03 | 8.76 |
| rs936891 | G | A | 0.69 | -7.23E-03 | 2.00E-03 | 3.00E-04 | 13.07 |
| rs9372734 | T | C | 0.51 | 9.59E-03 | 2.00E-03 | 1.63E-06 | 22.99 |
| rs9376591 | C | T | 0.39 | -8.41E-03 | 2.00E-03 | 2.61E-05 | 17.68 |
| rs9389886 | T | C | 0.39 | -8.65E-03 | 2.00E-03 | 1.53E-05 | 18.71 |
| rs9389889 | T | A | 0.39 | -8.44E-03 | 2.00E-03 | 2.44E-05 | 17.81 |
| rs9482094 | G | A | 0.61 | -8.66E-03 | 2.00E-03 | 1.49E-05 | 18.75 |
| rs9536468 | A | G | 0.15 | -6.21E-03 | 2.00E-03 | 1.90E-03 | 9.64 |
| rs9537938 | A | G | 0.73 | 4.79E-03 | 2.00E-03 | 1.66E-02 | 5.74 |
| rs9558931 | T | C | 0.38 | -6.94E-03 | 2.00E-03 | 5.20E-04 | 12.04 |
| rs9571763 | T | C | 0.60 | -8.78E-03 | 2.00E-03 | 1.13E-05 | 19.27 |
| rs9607805 | T | C | 0.70 | 8.76E-03 | 2.00E-03 | 1.19E-05 | 19.18 |
| rs9610866 | A | G | 0.36 | -4.77E-03 | 2.00E-03 | 1.71E-02 | 5.69 |
| rs9616812 | T | C | 0.49 | -2.94E-03 | 2.00E-03 | 1.42E-01 | 2.16 |
| rs962961 | T | C | 0.34 | -1.05E-02 | 2.00E-03 | 1.52E-07 | 27.56 |
| rs9632698 | T | A | 0.14 | 1.01E-02 | 2.00E-03 | 4.42E-07 | 25.50 |
| rs9653892 | A | T | 0.47 | 4.19E-03 | 2.00E-03 | 3.62E-02 | 4.39 |
| rs968050 | T | C | 0.51 | 9.59E-03 | 2.00E-03 | 1.63E-06 | 22.99 |
| rs9694306 | C | T | 0.30 | -9.53E-03 | 2.00E-03 | 1.89E-06 | 22.71 |
| rs9747342 | T | C | 0.25 | -8.21E-03 | 2.00E-03 | 4.04E-05 | 16.85 |
| rs9783783 | C | G | 0.50 | -1.02E-02 | 2.00E-03 | 3.40E-07 | 26.01 |
| rs9787623 | A | G | 0.15 | -9.44E-03 | 2.00E-03 | 2.36E-06 | 22.28 |
| rs980615 | A | G | 0.56 | 1.34E-03 | 2.00E-03 | 5.03E-01 | 0.45 |
| rs9812753 | G | C | 0.57 | -5.68E-03 | 2.00E-03 | 4.51E-03 | 8.07 |
| rs9841673 | T | A | 0.26 | 8.39E-03 | 2.00E-03 | 2.73E-05 | 17.60 |
| rs9858200 | A | G | 0.24 | -8.07E-03 | 2.00E-03 | 5.46E-05 | 16.28 |
| rs9859766 | G | A | 0.05 | 1.48E-02 | 4.00E-03 | 2.16E-04 | 13.69 |
| rs9859961 | A | T | 0.59 | -5.46E-03 | 2.00E-03 | 6.33E-03 | 7.45 |
| rs9862527 | A | G | 0.49 | 5.64E-03 | 2.00E-03 | 4.80E-03 | 7.95 |
| rs9883163 | T | C | 0.05 | 1.50E-02 | 4.00E-03 | 1.77E-04 | 14.06 |
| rs9918679 | T | G | 0.37 | -2.12E-03 | 2.00E-03 | 2.89E-01 | 1.12 |
| rs9925015 | C | T | 0.17 | 6.17E-03 | 2.00E-03 | 2.04E-03 | 9.52 |
| rs993560 | G | A | 0.33 | -6.33E-03 | 2.00E-03 | 1.55E-03 | 10.02 |
| rs9943780 | T | C | 0.24 | 7.99E-03 | 2.00E-03 | 6.47E-05 | 15.96 |
| rs9950738 | G | C | 0.31 | 7.97E-03 | 2.00E-03 | 6.75E-05 | 15.88 |
| rs9986203 | T | C | 0.60 | -7.00E-03 | 2.00E-03 | 4.65E-04 | 12.25 |

Abbreviations: SNP - single nucleotide polymorphisms, EAF - effect allele frequency, SE - standard error.

Source genome-wide association study is Saunders et al.^13^

Multi-allelic instruments were included given that all datasets clearly report multiple alleles allowing comparison.

Instruments were selected at genome-wide significance in the trans-ancestry analyses, but then ancestry-specific betas and standard errors were used.

As a result, some instruments had higher p values and lower F statistics. Post hoc choice of instruments, genetic models or data based on measured F-statistics can exacerbate bias.

In particular, the commonly cited rule of thumb that F > 10 avoids bias in IV analysis is misleading.^11^

#### Table S7: Comparing those with and without lifestyle survey data in Million Veteran Program.

|  | **Participants enrolled N=797,380** | **Participants with lifestyle data N=317,075*** |
| --- | --- | --- |
| **Age, *mean years ±S.D.*** | 66.43±11.73 | 58.13±14.93 |
| **Females, *N(%)*** | 7.39 | 10.41 |
| **Ethnicity, *Latin American, N(%)*** | 19945(6.29) | 45571(9.49) |

Abbreviations: N - number, S.D. - standard deviation.

*Returned at least some, not necessarily complete, survey data. N therefore differs from analyses requiring complete survey covariate data.

#### Table S8: Sociodemographic and clinical characteristics at lifestyle survey by drinking status, according to ancestry group, in Million Veteran Program.

|  | **EUROPEAN ANCESTRY** | | | **AFRICAN AMERICAN ANCESTRY** | | | **ADMIXED AMERICAN ANCESTRY** | | |
| --- | --- | --- | --- | --- | --- | --- | --- | --- | --- |
|  | **Never drinkers,** N=6496(4.3%) | **Former drinkers,** N= 62,395(41.0%) | **Current drinkers,** N=83,235(54.7%) | **Never drinkers,** N=815(4.7%) | **Former drinkers,** N=8549(49.1%) | **Current drinkers,** N=8055(46.2%) | **Never drinkers,** N=229(3.6%) | **Former drinkers,** N= 2984(46.5%) | **Current drinkers,** N=3202(49.9%) |
| **Age**, *mean years*±S.D. | 71.6±10.0 | 67.8±10.3 | 67.7±11.1 | 65.3±10.2 | 64.0±9.2 | 61.5±9.9 | 66.6±12.0) | 64.5±10.9 | 61.2±12.5 |
| **Sex**, *N(%) female* | 416(69.7) | 3210(52.9) | 4343(48.5) | 126(15.5) | 619(7.2) | 826(10.3) | 25(10.9) | 130(4.5) | 205(6.4) |
| **Smoking**,N(%) *daily* | 836(12.9) | 10,022(16.1) | 10,928(13.1) | 109(13.4) | 1612(18.9) | 2000(24.8) | 23(10.0) | 359(12.0) | 389(12.1) |
| **Smoking**,N(%) *never* | 5378(82.8) | 48,965(78.5) | 65,994(79.3) | 646(79.3) | 5935(69.4) | 4667(57.9) | 195(85.2) | 2363(79.2) | 2363(73.8) |
| **Income**, *N(%) <$10,000^2^* | 291(4.5) | 3071(4.9) | 2326(2.8)) | 87(10.7) | 1033(12.1) | 856(10.6) | 23(10.0) | 233(7.8) | 155(4.8) |
| *$10,000-$19,999* | 1197(18.4) | 11066(17.7) | 8966(10.8) | 143(17.5) | 1865(21.8) | 1528(19.0) | 48(21.0) | 574(19.2) | 424(13.2) |
| *$20,000-$29,999* | 1266(19.5) | 11182(17.9) | 10932(13.1) | 104(12.8) | 1222(14.3) | 1043(12.9) | 43(18.8) | 479(16.1) | 401(12.5) |
| *$30,000-$39,999* | 1067(16.4) | 9035(14.5) | 11122(13.4) | 101(12.3) | 1104(12.9) | 953(11.8) | 31(13.5) | 417(14.0) | 477(14.9) |
| *$40,000-$49,999* | 730(11.2) | 6860(11.0) | 9433(11.3) | 88(10.8) | 788(9.2) | 740(9.2) | 15(16.6) | 334(11.2) | 382(11.9) |
| *$50,000-$59,999* | 445(6.9) | 5074(8.1) | 8232(9.9) | 69(8.5) | 612(7.2) | 634(7.9) | 22(9.6) | 264(8.8) | 302(9.4) |
| *$60,000-$69,999* | 447(6.9) | 4869(7.8) | 8473(10.2) | 70(8.6) | 598(7.0) | 671(8.3) | 14(6.1) | 231(7.7) | 315(9.8) |
| *$70,000-$74,999* | 273(4.2) | 3608(5.8) | 7823(9.4) | 43(5.3) | 399(4.7) | 508(6.3) | 8(3.5) | 139(4.7) | 267(8.3) |
| *$75,000-$99,999* | 163(2.5) | 2139(3.4) | 6087(7.3) | 23(2.8) | 216(2.5) | 373(4.6) | 8(3.5) | 67(2.2) | 188(5.9) |
| *Over $150,000^3^* | 58(0.9) | 600(1.0) | 2523(3.0) | 6(0.7) | 48(0.6) | 135(1.7) | 3(1.3) | 18(0.6) | 51(1.6) |
| **Income, N(%),** prefer not to answer | 559(8.6) | 4891(7.8) | 7318(8.8) | 81(0.7) | 664(0.6) | 614(7.6) | 14(6.1) | 228(7.6) | 240(7.5) |
| **Education**, N(%) *less than high school^4^* | 514(7.9) | 3242(5.2) | 1864(2.2) | 51(6.3) | 523(6.1) | 294(3.6) | 23(10.0) | 214(7.2) | 138(1.7) |
| *High school diploma/GED* | 1998(30.8) | 17695(28.4) | 16383(19.7) | 216(26.5) | 2375(27.8) | 1858(23.1) | 61(26.7) | 822(27.5) | 636(19.9) |
| *Some college credit, but no degree* | 1954(30.1) | 20555(32.9) | 25207(30.3) | 249(30.6) | 3144(36.8) | 2951(36.6) | 77(33.6) | 1059(35.5) | 1146(35.8) |
| *Associates degree* | 684(10.5) | 7494(12.0) | 10433(12.5) | 112(13.7) | 1183(13.8) | 1154(14.3) | 24(33.6) | 436(14.6) | 553(17.3) |
| *Bachelor's degree* | 823(12.7) | 8554(13.7) | 17352(20.8) | 118(14.5) | 820(9.6) | 1098(13.6) | 23(10.5) | 296(9.9) | 466(14.6) |
| *Master's degree* | 355(5.05) | 3421(5.5) | 8790(10.6) | 47(5.8) | 385(4.5) | 567(7.0) | 16(10.0) | 121(4.1) | 207(6.5) |
| **Education**, N(%) *professional/doctoral degree* | 168(2.6) | 1434(2.6) | 3206(3.9) | 22(2.7) | 119(1.4) | 133(1.7) | 5(7.0) | 36(7.0) | 56(1.7) |
| **Dementia cases**, N(%) | 450(6.9) | 3130(5.0) | 2762(3.3) | 43(5.3) | 431(5.0) | 221(2.7) | 18(7.9) | 155(5.2) | 90(2.8) |
| **Opioid dependence,** N(%) | 173(2.7) | 2624(4.2) | 1345(1.6) | 34(4.2) | 7792(8.9) | 7596(5.7) | 10(4.4) | 169(5.7) | 86(2.7) |
| **Alcohol Use Disorder,** N(%) | 327(5.0) | 12,553(20.1) | 16877(20.3) | 88(10.8) | 2978(34.8) | 3314(41.1) | 32(14.0) | 813(27.2) | 1048(32.7) |
| **Cannabis Use Disorder,** N(%) | 125(1.9) | 2786(4.4) | 2146(2.6) | 34(4.2) | 1059(12.4) | 976(12.1) | 9(3.9) | 218(7.3) | 183(5.7) |
| **Died during follow up,** N(%) | 1500(23.1) | 11,177(17.9) | 10,169(12.2) | 116(12.2) | 1206(14.1) | 744(9.2) | 27(11.8) | 353(11.8) | 203(6.3) |

Abbreviations: N - number, S.D. - standard deviation.

#### Table S9: Characteristics by beverage preference, in Million Veteran Program.

|  | **Wine drinkers** | **Beer drinkers** | **Liquor drinkers** |
| --- | --- | --- | --- |
|  | N=5016(24.20%) | N=9552(46.08) | N=5762(27.80) |
| **Age,** *years***±S.D.** | 72±9.44 | 65.56±9.99 | 71±10.01 |
| **Total ethanol daily,** *grams* | 27.69±16.29 | 38.01±25.22 | 36.77±23.26 |
| **Dementia cases,** N(%) | 190(3.79) | 209(2.19) | 219(3.80) |
| **Education, N(%)** *less than high school* | 60(1.20) | 245(2.56) | 90(1.56) |
| **Education, N(%)** *professional or doctorate degree* | 408(8.13) | 209(2.19) | 318(5.52) |
| **Income, N(%)** *<$10,000* | 52(1.04) | 391(4.09) | 118(2.05) |
| **Income, N(%)** *>$150,000* | 348(6.94) | 190(1.99) | 253(4.39) |
| **Association incident dementia, HR[95% CI]*** | 1.17[0.95-1.46] | 1.20[0.98-1.47] | REF |

*Estimates generated from Cox proportional hazards model with incident all-cause dementia as the outcome, and spirit drinking frequency as reference group.

Models were controlled for: total ethanol daily, age, sex, education, income, smoking, post-traumatic stress disorder, depression, head injury, substance use.

Abbreviations: S.D – standard deviation, N – number, HR – hazard ratio, CI – confidence interval, REF – reference group.

#### Table S10: Aalen-Johansen estimates, Million Veteran Program.

|  | **State** | **Total N** | **Event N** | **Mean time in state (days)** |
| --- | --- | --- | --- | --- |
| **Alcohol consumption, drinks weekly** |  |  |  |  |
| Non-drinker | censored | 76,636 | 0 | 2656.59 |
| <7 | censored | 109,495 | 0 | 2804.21 |
| 7- <14 | censored | 12,526 | 0 | 2835.58 |
| 14- <22 | censored | 3214 | 0 | 2774.25 |
| 22- <40 | censored | 1136 | 0 | 2766.81 |
| >40 | censored | 1358 | 0 | 2664.65 |
| Non-drinker | death | 76,636 | 12,746 | 468.64 |
| <7 | death | 109,495 | 13,416 | 365.78 |
| 7- <14 | death | 12,526 | 1479 | 360.32 |
| 14- <22 | death | 3214 | 446 | 413.21 |
| 22- <40 | death | 1136 | 168 | 428.25 |
| >40 | death | 1358 | 235 | 504.35 |
| Non-drinker | dementia | 76,636 | 4193 | 151.78 |
| <7 | dementia | 109,495 | 3991 | 107.01 |
| 7- <14 | dementia | 12,526 | 340 | 81.10 |
| 14- <22 | dementia | 3214 | 97 | 89.55 |
| 22- <40 | dementia | 1136 | 32 | 81.94 |
| >40 | dementia | 1358 | 50 | 108.01 |
| **Alcohol Use Disorder** |  |  |  |  |
| Control | censored | 159,389 | 0 | 2746.54 |
| Case | censored | 44,976 | 0 | 2747.80 |
| Control | death | 159,389 | 22,100 | 409.12 |
| Case | death | 44,976 | 6390 | 402.67 |
| Control | dementia | 159,389 | 6673 | 121.34 |
| Case | dementia | 44,976 | 2030 | 126.53 |

Values show how long on average an individual moving through the study spends in each potential state: censored, death or dementia, according to their alcohol intake.

#### Table S11: Competing risk regression compared to Cox proportional hazards regression estimates, Million Veteran Program.

|  |  | **Cox regression** | | | **Competing risk regression** | | |
| --- | --- | --- | --- | --- | --- | --- | --- |
| **Alcohol intake** | *N* | *HR* | *95% CI* | *P value* | *HR* | *95% CI* | *P value* |
| Nondrinker | 76636 | 1.37 | 1.31-1.43 | <2E-16 | 1.34 | 1.30-1.38 | <2E-16 |
| <7 | 109495 | REF | REF | REF | REF | REF | REF |
| 7- <14 | 12526 | 0.94 | 0.84-1.06 | 0.31 | 0.94 | 0.83-1.05 | 0.25 |
| 14- <22 | 3214 | 1.11 | 0.91-1.36 | 0.31 | 1.09 | 0.89-1.29 | 0.39 |
| 22- <40 | 1136 | 1.14 | 0.80-1.62 | 0.46 | 1.1 | 0.75-1.45 | 0.6 |
| >40 | 1358 | 1.61 | 1.22-2.14 | 9.12E-04 | 1.51 | 1.23-1.79 | 4.70E-03 |
| Alcohol Use Disorder | 41399 | 1.51 | 1.42-1.60 | <2E-16 | 1.47 | 1.41-1.53 | <2E-16 |

Abbreviations: N – number, HR - hazard ratio, CI - confidence interval, REF - reference group.

#### Table S12: Dietary and serum/blood thiamine according to alcohol intake, Million Veteran Program.

|  | **Dietary thiamine w/o supplementation, N=135,144** | **Dietary thiamine with supplementation, N=135,144** | **Low serum or whole blood thiamine recorded in EHR, N=7723** |
| --- | --- | --- | --- |
| **Non drinker** | 0.98±0.47 | 9.15±17.9 | 185(4.64) |
| **Drinks per week** |  |  |  |
| <7 | 0.99±0.43 | 9.11±17.9 | 260(5.49) |
| 7- <14 | 0.98±0.42 | 9.66±18.4 | 60(7.27) |
| 14- <22 | 0.95±0.40 | 9.33±18.9 | 24(6.96) |
| 22- <40 | 0.97±0.42 | 10.01±18.9 | 12(7.10) |
| >40 | 0.99±0.44 | 9.46±18.3 | 12(4.46) |
| **No history of Alcohol Use Disorder** | 0.98±0.44 | 9.03±17.8 | 273(4.66) |
| **Alcohol Use Disorder** | 0.98±0.46 | 9.96±18.75 | 285(6.27) |
| **Association Alcohol Use Disorder controlling for thiamine** | 1.51[1.40-1.62] | 1.50[1.39-1.61] | 1.32[1.15-1.53] |

Abbreviations: w/o - without, N - number, EHR - electronic health record.

#### Table S13: Alcohol dementia associations in Million Veteran Program, by ancestry group.

|  | **AVERAGE OF 9 YEARS PRIOR TO DIAGNOSIS** | | | | | | | | | | | |  | **AVERAGE OF 4 YEARS PRIOR TO DIAGNOSIS** | | | | | | | |
| --- | --- | --- | --- | --- | --- | --- | --- | --- | --- | --- | --- | --- | --- | --- | --- | --- | --- | --- | --- | --- | --- |
|  | **European ancestry** | | | | **African ancestry** | | | | **Latin American ancestry** | | | |  | **European ancestry** | | | | **African ancestry** | | | |
| **Alcohol phenotype** | N | HR | 95% CI | P value | N | HR | 95% CI | P value | N | HR | 95% CI | P value | **Alcohol phenotype** | N | HR | 95% CI | P value | N | HR | 95% CI | P value |
| **Drinks per week** |  |  |  |  |  |  |  |  |  |  |  |  | **Drinks per week** |  |  |  |  |  |  |  |  |
| Non drinker | 76636 | 1.37 | 1.31-1.43 | <2E-16 | 14115 | 1.2 | 1.07-1.35 | 1.95E-03 | 4166 | 1.39 | 1.12-1.73 | 2.74E-03 | Never drinker | 5645 | 1.39 | 1.25-1.56 | 5.34E-09 | 674 | 1.23 | 0.85-1.78 | 2.70E-01 |
|  |  |  |  |  |  |  |  |  |  |  |  |  | Former drinker | 42623 | 1.27 | 1.19-1.35 | 8.54E-14 | 5101 | 1.19 | 0.97-1.45 | 0.09 |
| <7 | 109495 | REF | REF | REF | 14543 | REF | REF | REF | 5331 | REF | REF | REF | <7 | 59143 | REF | REF | REF | 6340 | REF | REF | REF |
| 7- <14 | 12526 | 0.94 | 0.84-1.06 | 0.31 | 2308 | 0.92 | 0.71-1.20 | 0.54 | 655 | 1.3 | 0.84-2.02 | 0.23 | 7- <14 | 12368 | 0.87 | 0.78-0.97 | 8.96E-03 | 1312 | 1.07 | 0.75-1.53 | 0.7 |
| 14- <22 | 3214 | 1.11 | 0.91-1.36 | 0.31 | 601 | 1.13 | 0.78-1.80 | 0.6 | 231 | 1.04 | 0.46-2.34 | 0.92 | 14- <22 | 7704 | 0.83 | 0.72-0.96 | 0.01 | 630 | 0.64 | 0.35-1.15 | 0.13 |
| 22- <40 | 1136 | 1.14 | 0.80-1.62 | 0.46 | 229 | 1.07 | 0.51-2.28 | 0.85 | 114 | 1.53 | 0.47-4.96 | 0.48 | 22- <40 | 2701 | 1.04 | 0.82-1.31 | 7.60E-01 | 318 | 1.74 | 1.00-3.02 | 0.05 |
| >40 | 1358 | 1.61 | 1.22-2.14 | 9.12E-04 | 366 | 1.33 | 0.78-2.28 | 0.29 | 112 | 1.76 | 0.71-4.40 | 0.22 | >40 | 941 | 0.97 | 0.64-1.47 | 0.9 | 125 | 2.49 | 1.21-5.14 | 0.01 |
| **Lifetime diagnosis AUD** | 41,399 | 1.51 | 1.42-1.60 | <2E-16 | 11,392 | 1.44 | 1.25-1.67 | 9.44E-07 | 3205 | 1.58 | 1.24-2.01 | 2.05E-04 |  |  |  |  |  |  |  |  |  |

Abbreviations: N - number, HR - hazard ratio, CI - confidence interval, AUD - alcohol use disorder, REF - reference group.

#### Table S14: Alcohol associations with dementia subtypes, in Million Veteran Program.

|  | **All-cause dementia** | | | | **Alzheimer's disease** | | | | **Vascular dementia** | | | |
| --- | --- | --- | --- | --- | --- | --- | --- | --- | --- | --- | --- | --- |
|  | **N=8703** | | | | **N=2038** | | | | **N=2271** | | | |
| **Alcohol phenotype** | *N* | *HR* | *95% CI* | *P value* | *N* | *HR* | *95% CI* | *P value* | *N* | *HR* | *95% CI* | *P value* |
| **Drinks per week** |  |  |  |  |  |  |  |  |  |  |  |  |
| Non drinker | 76,636 | 1.37 | 1.31-1.43 | <2E-16 | 73,424 | 1.42 | 1.30-1.56 | 4.84E-14 | 73,354 | 1.51 | 1.39-1.65 | <2E-16 |
| <7 | 109,495 | REF | REF | REF | 106,459 | REF | REF | REF | 106,296 | REF | REF | REF |
| 7- <14 | 12,526 | 0.94 | 0.84-1.06 | 0.31 | 12,265 | 1.04 | 0.83-1.31 | 0.73 | 12,250 | 0.86 | 0.68-1.08 | 0.2 |
| 14- <22 | 3214 | 1.11 | 0.91-1.36 | 0.31 | 3130 | 0.79 | 0.46-1.37 | 0.41 | 3135 | 1.12 | 0.75-1.67 | 0.59 |
| 22- <40 | 1136 | 1.14 | 0.80-1.62 | 0.46 | 1108 | 0.85 | 0.32-2.29 | 0.75 | 1111 | 0.98 | 0.46-2.06 | 0.96 |
| >40 | 1358 | 1.61 | 1.22-2.14 | 9.12E-04 | 1314 | 1.33 | 0.59-2.98 | 0.49 | 1319 | 1.65 | 0.95-2.88 | 0.08 |
| **LIFETIME AUD** | 41,399 | 1.51 | 1.42-1.60 | <2E-16 | 43,492 | 1.27 | 1.12-1.44 | 2.33E-04 | 43,559 | 1.43 | 1.28-1.60 | 1.27E-10 |

Drinks per week derived from first recorded AUDIT-C in electronic health record. Abbreviations: N - number, HR - hazard ratio, CI - confidence interval, AUD - alcohol use disorder, REF - reference group.

#### Table S15: Association between alcohol binging frequency and all-cause incident dementia, in Million Veteran Program.

| **Alcohol binging frequency** | **N** | **HR** | **95% CI** | **p** |
| --- | --- | --- | --- | --- |
| Never | 150,129 | REF | REF | REF |
| Less than monthly | 14,045 | 0.97 | 0.87-1.08 | 0.55 |
| Monthly | 5237 | 0.87 | 0.72-1.07 | 0.17 |
| Weekly | 5023 | 1.04 | 0.87-1.26 | 0.66 |
| Daily or almost daily | 4641 | 1.31 | 1.05-1.64 | 0.02 |

An alcohol binge was defined as >6 drinks on one occasion, at the first recorded AUDIT-C on record. Models controlled for total proxy drinks per week in addition to covariates. Abbreviations: N - number, HR - hazard ratio, CI - confidence interval, REF - reference group.

#### Table S16: Genome-wide significant SNPs for all-cause dementia in Million Veteran Program, by ancestry.

| **SNP** | **Chromosome** | **Position** | **Closest gene(s)** | **Other allele** | **Effect allele** | **EAF** | **N** | **OR** | **Beta** | **SE** | **LCI** | **UCI** | **P value** | **Ancestry** |
| --- | --- | --- | --- | --- | --- | --- | --- | --- | --- | --- | --- | --- | --- | --- |
| rs3851179 | 11 | 85868640 | PICALM | T | C | 0.64 | 445,784 | 1.06 | 0.03 | 0.01 | 1.04 | 1.08 | 1.70E-10 | EUR |
| rs429358 | 19 | 45411941 | APOE | T | C | 0.14 | 445,784 | 1.69 | 0.52 | 0.12 | 1.65 | 1.73 | 1.14E-314 | EUR |
| rs111371860 | 19 | 45345787 | BCAM, NECTIN2 | A | T | 0.07 | 445,784 | 0.87 | -0.06 | 0.02 | 0.84 | 0.91 | 8.60E-11 | EUR |
| rs744373 | 2 | 127894615 | BIN1, NIFKP9 | A | G | 0.29 | 445,784 | 1.07 | 0.03 | 0.01 | 1.05 | 1.09 | 5.19E-11 | EUR |
| rs429358 | 19 | 45411941 | APOE | T | C | 0.22 | 112,506 | 1.36 | 0.31 | 0.02 | 1.3 | 1.43 | 5.38E-41 | AFR |

Abbreviations: SNP - single nucleotide polymorphism, EAF - effect allele frequency, OR - odds ratio, SE - standard error, LCI - lower confidence interval, UCI - upper confidence interval.

#### Table S17: Heritabilities for key phenotypes.

| **Phenotype** | **Observed h2** | **Liability h2** | **Lambda GC** | **Mean Chi^2** | **Intercept** | **Ratio** |
| --- | --- | --- | --- | --- | --- | --- |
| All-cause dementia | 0.0075(0.0013) | 0.025(0.04) | 1.07 | 1.09 | 1.02(0.009) | 0.21(0.11) |
| Proxy Alzheimer's disease | 0.01(0.001) | 0.031(0.006) | 1.2 | 1.27 | 1.07(0.01) | 0.25(0.04) |
| Alcohol Use Disorder | 0.09(0.004) | 0.13(0.005) | 1.53 | 1.71 | 1.04(0.01) | 0.05(0.02) |
| Problematic Alcohol Use | 0.05(0.003) |  | 1.23 | 1.27 | 1.02(0.008) | 0.08(0.03) |
| Drinks per week | 0.05(0.002) |  | 2.49 | 3.2 | 1.12(0.01) | 0.13(0.01) |

Abbreviations: h2 - heritability, GC - genomic control.

#### Table S18: Mendelian randomization estimates including robust methods, by ancestry.

|  |  | **European ancestry** | | | | | | | **African ancestry** | | | | | | |
| --- | --- | --- | --- | --- | --- | --- | --- | --- | --- | --- | --- | --- | --- | --- | --- |
| **Alcohol phenotype** | **MR method** | **SNPs** | **Beta** | **LCI** | **UCI** | **p** | **IVW heterogeneity** | **Pleiotropy** | **SNPs** | **Beta** | **LCI** | **UCI** | **p** | **IVW heterogeneity** | **Pleiotropy** |
| Alcohol use disorder | MR Egger | 66 | -0.12 | -0.43 | 0.18 | 4.28E-01 |  |  | 4 | 0.15 | -1.12 | 1.43 | 0.83 |  |  |
|  | Weighted median | 66 | -0.05 | -0.26 | 0.16 | 6.51E-01 | 124.61(1.25E-05) | 0.01 | 4 | 0.34 | -0.18 | 0.86 | 0.2 | 0.69(0.87) | 0.79 |
|  | Inverse variance weighted | 66 | 0.21 | 0.04 | 0.37 | 1.36E-02 |  |  | 4 | 0.34 | -0.12 | 0.81 | 0.15 |  |  |
|  | Weighted mode | 66 | -0.05 | -0.27 | 0.17 | 6.34E-01 |  |  | 4 | 0.2 | -0.4 | 0.81 | 0.56 |  |  |
|  | MR PRESSO | 66 | 0.16 | 0.08 | 0.24 | 3.57E-02 |  |  |  |  |  |  |  |  |  |
| Problematic alcohol use | MR Egger | 80 | -0.06 | -0.37 | 0.25 | 7.21E-01 |  |  |  |  |  |  |  |  |  |
|  | Weighted median | 80 | -0.04 | -0.25 | 0.18 | 7.47E-01 | 132.68(1.45E-04) | 0.03 |  |  |  |  |  |  |  |
|  | Inverse variance weighted | 80 | 0.25 | 0.08 | 0.42 | 3.87E-03 |  |  |  |  |  |  |  |  |  |
|  | Weighted mode | 80 | -0.06 | -0.28 | 0.15 | 5.63E-01 |  |  |  |  |  |  |  |  |  |
|  | MR PRESSO | 80 | 0.23 | 0.14 | 0.31 | 7.57E-03 |  |  |  |  |  |  |  |  |  |
| Drinks per week | MR Egger | 641 | 0.49 | 0.30 | 0.69 | 8.85E-07 |  |  |  |  |  |  |  |  |  |
|  | Weighted median | 641 | -0.05 | -0.22 | 0.13 | 6.02E-01 | 921.30(1.74E-12) | 2.73E-05 |  |  |  |  |  |  |  |
|  | Inverse variance weighted | 641 | 0.14 | 0.03 | 0.24 | 1.01E-02 |  |  |  |  |  |  |  |  |  |
|  | Weighted mode | 641 | 0.14 | -3.37 | 3.65 | 9.37E-01 |  |  |  |  |  |  |  |  |  |
|  | MR PRESSO | 641 | 0.09 | 0.04 | 0.14 | 8.00E-02 |  |  |  |  |  |  |  |  |  |

Abbreviations: MR - Mendelian randomization, SNPs - single nucleotide polymorphisms, LCI - lower confidence interval, UCI - upper confidence interval, LCI - lower confidence interval.

#### Table S19: Mendelian randomization estimates corrected for sample overlap using MRLap.

| **Alcohol phenotype** | **Standard IVW*** | | **MRlap 10000kb prune** | | | |
| --- | --- | --- | --- | --- | --- | --- |
|  | *SNPs* | *Beta, SE (p)* | *SNPs* | *Observed Beta, SE (p)* | *Corrected Beta, SE (p)* | *Difference (p)* |
| **Alcohol Use Disorder** | 66 | 0.21, 0.08, (1.36E-02) | 49 | 0.07, 0.03 (6.56E-03) | 0.09, 0.03 (0.01) | -1.63(0.10) |
| **Problematic Alcohol Use** | 80 | 0.25, 0.09(3.87E-03) | 54 | 0.08, 0.03(6.63E-03) | 0.09, 0.04(0.01) | -1.81(0.07) |

*Includes proxy SNPs.

Abbreviations: MR - Mendelian randomization, IVW - inverse variance weighted, kb - kilobase, SNPs - single nucleotide polymorphisms, SE - standard error.

#### Table S20: Reverse Mendelian randomization estimates.

|  | **Alcohol Use Disorder** | | | | | | | **Problematic Alcohol Use** | | | | | | | **Drinks per week** | | | | | | |
| --- | --- | --- | --- | --- | --- | --- | --- | --- | --- | --- | --- | --- | --- | --- | --- | --- | --- | --- | --- | --- | --- |
|  | **p<5x10E^-8^ THRESHOLD** | | | | | | | | | | | | | | | | | | | | |
|  | SNPs | Beta | LCI | UCI | p value | Heterogeneity | Pleiotropy | SNPs | Beta | LCI | UCI | p value | Heterogeneity | Pleiotropy | SNPs | Beta | LCI | UCI | p value | Heterogeneity | Pleiotropy |
| **MR method** |  |  |  |  |  |  |  |  |  |  |  |  |  |  |  |  |  |  |  |  |  |
| **MR Egger** | 4 | 1.68E-02 | -4.11E-03 | 3.78E-02 | 2.56E-01 |  |  | 4 | 3.45E-03 | -1.63E-02 | 2.32E-02 | 7.65E-01 |  |  | 4 | -1.62E-04 | -1.06E-02 | 1.02E-02 | 9.78E-01 |  |  |
| **Weighted median** | 4 | 5.90E-03 | -8.15E-03 | 2.00E-02 | 4.10E-01 |  |  | 4 | -4.21E-03 | -2.07E-02 | 1.23E-02 | 6.16E-01 |  |  | 4 | -4.06E-03 | -1.24E-02 | 4.31E-03 | 3.42E-01 |  |  |
| **Inverse variance weighted** | 4 | 3.68E-03 | -1.75E-02 | 2.49E-02 | 7.34E-01 | 8.78(0.03) | 0.03 | 4 | -5.27E-03 | -1.99E-02 | 9.34E-03 | 4.80E-01 | 1.67(0.64) | 0.33 | 4 | -4.41E-03 | -1.17E-02 | 2.87E-03 | 2.35E-01 | 1.30(0.73) | 0.38 |
| **Weighted mode** | 4 | 6.96E-03 | -5.79E-03 | 1.97E-02 | 3.63E-01 |  |  | 4 | -2.85E-03 | -1.85E-02 | 1.28E-02 | 7.44E-01 |  |  | 4 | -3.76E-03 | -1.15E-02 | 3.95E-03 | 4.10E-01 |  |  |
|  | **p<5x10E^-5^ THRESHOLD** | | | | | | | | | | | | | | | | | | | | |
| **MR Egger** | 99 | -9.44E-03 | -2.80E-02 | 9.14E-03 | 3.22E-01 |  |  | 103 | -3.68E-03 | -2.28E-02 | 1.54E-02 | 7.07E-01 |  |  | 103 | -6.85E-03 | -2.31E-02 | 9.41E-03 | 4.11E-01 |  |  |
| **Weighted median** | 99 | -5.70E-03 | -2.01E-02 | 8.73E-03 | 4.39E-01 |  |  | 103 | 2.59E-03 | -1.37E-02 | 1.89E-02 | 7.55E-01 |  |  | 103 | 3.42E-03 | -5.15E-03 | 1.20E-02 | 4.34E-01 |  |  |
| **Inverse variance weighted** | 99 | 9.11E-03 | -4.28E-03 | 2.25E-02 | 1.82E-01 | 210.32(3.49E-10) | 7.45E-03 | 103 | 5.37E-03 | -7.48E-03 | 1.82E-02 | 4.13E-01 | 168.25(4.02E-05) | 0.21 | 103 | 4.60E-03 | -6.90E-03 | 1.61E-02 | 4.33E-01 | 466.95(4.32E-48) | 0.06 |
| **Weighted mode** | 99 | -5.59E-03 | -1.90E-02 | 7.77E-03 | 4.14E-01 |  |  | 103 | 1.97E-03 | -1.31E-02 | 1.71E-02 | 7.99E-01 |  |  | 103 | 3.84E-03 | -3.58E-03 | 1.13E-02 | 3.13E-01 |  |  |

Estimated causal effect of all-cause dementia on alcohol use. Abbreviations: SNPs - single nucleotide polymorphisms, LCI - lower confidence interval, UCI - upper confidence interval, MR - Mendelian randomization.

#### Table S21: Earliest recorded alcohol intake according to ADH1B genotype in Million Veteran Program and UK Biobank.

|  | **Million Veteran Program*** | | | **UK Biobank** | | |
| --- | --- | --- | --- | --- | --- | --- |
|  | ***ADH1B* (rs1229984)** | | | ***ADH1B* (rs1229984)** | | |
| **Drinking behaviour** | **TT** | **TC** | **CC** | **TT** | **TC** | **CC** |
|  | N(%) | N(%) | N(%) | N(%) | N(%) | N(%) |
| **Non drinker** | 194(36.7) | 5362(38.0) | 70,885(37.5) |  |  |  |
| **Never drinker** |  |  |  | 224(23.2) | 1237(7.9) | 12,346(4.2) |
| **Former drinker** |  |  |  | 45(4.7) | 703(4.5) | 11,749(4.0) |
| **Drinks per week** |  |  |  |  |  |  |
| ***<7*** | 321(60.7) | 8100(57.4) | 100,767(53.3) | 486(50.3) | 7603(48.3) | 116,753(40.0) |
| ***7- <14*** | 11(2.1) | 480(3.4) | 12,012(6.4) | 145(15.0) | 3753(23.8) | 78,465(26.9) |
| ***14- <22*** | 1(0.2) | 95(0.7) | 3113(1.6) | 46(4.8) | 1517(9.6) | 40,767(14.0) |
| ***22- <40*** | 1(0.2) | 36(0.3) | 1099(0.6) | 18(1.9) | 818(5.2) | 25,688(8.9) |
| ***>40*** | 1(0.2) | 31(0.2) | 1325(0.7) | 3(0.3) | 106(0.7) | 6310(2.2) |

*First recorded alcohol intake.

#### Table S22: Multivariable Mendelian randomization estimates.

|  |  | **MVIVW** | | |
| --- | --- | --- | --- | --- |
|  | **SNPs** | **Beta** | **95%CI** | **p value** |
| **Alcohol Use Disorder** |  |  |  |  |
| Smoking | 58 | 0.19 | 0.01, 0.37 | 3.80E-02 |
| Income | 66 | 0.13 | [-0.07, 0.32] | 2.02E-01 |
| Global brain volume | 58 | 0.21 | 0.00, 0.41 | 4.60E-02 |
| Hippocampal volume | 58 | 0.20 | 0.02, 0.38 | 3.40E-02 |
| White matter hyperintensity volume | 58 | 0.18 | 0.00,0.36 | 5.00E-02 |
| Post-traumatic stress disorder symptoms | 65 | 0.16 | -0.03,0.35 | 1.00E-01 |
| Cannabis frequency | 64 | 0.27 | 0.08,0.47 | 7.00E-03 |
| **Problematic Alcohol Use** |  |  |  |  |
| Post-traumatic stress disorder symptoms | 79 | 0.20 | 0.02,0.39 | 3.00E-02 |
| Cannabis frequency | 79 | 0.36 | 0.16,0.55 | <0.0001 |
| **Drinks per week** |  |  |  |  |
| Smoking | 668 | 0.14 | 0.03, 0.24 | 9.00E-03 |
| Income | 666 | 0.14 | 0.04, 0.245 | 9.00E-03 |
| Global brain volume | 664 | 0.12 | 0.02, 0.22 | 2.50E-02 |
| Hippocampal volume | 664 | 0.13 | 0.02, 0.23 | 1.80E-02 |
| White matter hyperintensity volume | 664 | 0.12 | 0.02, 0.23 | 1.80E-02 |
| Post-traumatic stress disorder symptoms | 659 | 0.17 | 0.07,0.27 | 0.001 |
| Cannabis frequency | 641 | 0.17 | 0.06,0.28 | 0.003 |

Abbreviations: MVIVW - multivariable inverse variance weighted, SNPs - single nucleotide polymorphisms, CI - confidence interval.

#### Table S23: Mendelian randomization estimates for alcohol and differing dementia phenotypes.

|  |  | **Proxy dementia** | | | | | | | **Clinically diagnosed Alzheimer's disease** | | | | | | |
| --- | --- | --- | --- | --- | --- | --- | --- | --- | --- | --- | --- | --- | --- | --- | --- |
| **Alcohol phenotype** | **MR method** | **SNPs** | **Beta** | **LCI** | **UCI** | **p** | **IVW heterogeneity** | **Pleiotropy** | **SNPs** | **Beta** | **LCI** | **UCI** | **p** | **IVW heterogeneity** | **Pleiotropy** |
| Alcohol use disorder | MR Egger | 64 | 0.01 | -0.06 | 0.09 | 7.51E-01 |  |  | 51 | -0.08 | -0.51 | 0.35 | 7.23E-01 |  |  |
|  | Weighted median | 64 | 0.03 | -0.01 | 0.07 | 1.99E-01 | 151.43(3.04E-09) | 7.79E-04 | 51 | -0.08 | -0.40 | 0.23 | 5.99E-01 | 84.68(1.58E-03) | 0.17 |
|  | Inverse variance weighted | 64 | 0.02 | -0.01 | 0.06 | 2.26E-01 |  |  | 51 | 0.17 | -0.09 | 0.43 | 1.95E-01 |  |  |
|  | Weighted mode | 64 | 0.03 | -0.02 | 0.07 | 2.20E-01 |  |  | 51 | -0.13 | -0.44 | 0.18 | 4.14E-01 |  |  |
| Problematic alcohol use | MR Egger | 67 | 0.02 | -0.06 | 0.09 | 6.93E-01 |  |  | 75 | -0.03 | -0.44 | 0.38 | 8.89E-01 |  |  |
|  | Weighted median | 67 | 0.04 | -0.01 | 0.08 | 1.43E-01 | 162.93(1.55E-08) | 6.39E-04 | 75 | -0.09 | -0.44 | 0.26 | 6.24E-01 | 104.36(0.01) | 0.1 |
|  | Inverse variance weighted | 67 | 0.04 | 0.00 | 0.07 | 5.98E-02 |  |  | 75 | 0.25 | 0.01 | 0.49 | 4.35E-02 |  |  |
|  | Weighted mode | 67 | 0.03 | -0.02 | 0.09 | 2.09E-01 |  |  | 75 | -0.13 | -0.46 | 0.20 | 4.50E-01 |  |  |
| Drinks per week (trans-ethnic SNPs) | MR Egger | 774 | 0.06 | 0.02 | 0.10 | 2.26E-03 |  |  | 770 | 0.25 | -0.02 | 0.52 | 6.78E-02 |  |  |
|  | Weighted median | 774 | 0.04 | 0.00 | 0.07 | 5.65E-02 | 1174.68(4.02E-19) | 1.60E-01 | 770 | -0.08 | -0.35 | 0.18 | 5.38E-01 | 991.60(8.85E-08) | 7.60E-01 |
|  | Inverse variance weighted | 774 | 0.04 | 0.02 | 0.05 | 1.26E-04 |  |  | 770 | 0.22 | 0.08 | 0.36 | 2.54E-03 |  |  |
|  | Weighted mode | 774 | 0.04 | -0.27 | 0.35 | 8.00E-01 |  |  | 770 | -0.22 | -1.26 | 0.82 | 6.74E-01 |  |  |

Abbreviations: MR - Mendelian randomization, SNPs - single nucleotide polymorphisms, LCI - lower confidence interval, UCI - upper confidence interval, IVW - inverse variance weighted.

#### Table S24: Localized average causal effect estimates for alcohol intake on dementia, for five alcohol strata, including nondrinkers.

| **Stratum** | **Mean alcohol exposure, DPW** | **Beta** | **LCI** | **UCI** | **P value** |
| --- | --- | --- | --- | --- | --- |
| 1 | 0.06 | -8.83E-03 | -3.12E+00 | 3.10E+00 | 9.96E-01 |
| 2 | 0.38 | 3.50E-02 | -5.83E-01 | 6.53E-01 | 9.11E-01 |
| 3 | 1.34 | 6.71E-02 | -1.29E-01 | 2.63E-01 | 5.03E-01 |
| 4 | 3.86 | 4.25E-02 | -4.63E-02 | 1.31E-01 | 3.48E-01 |
| 5 | 11.66 | 8.76E-02 | 3.90E-02 | 1.36E-01 | 4.12E-04 |

Estimates generated using the doubly-ranked nonlinear Mendelian randomization method, in European ancestry unrelated participants of Million Veteran Program (N= 313,873). Adjusted for: age, age^2^, sex, and top ten principal ancestry components. Abbreviations: DPW – drinks per week, LCI – lower confidence interval, UCI – upper confidence interval.

#### Table S25: Localized average causal effect estimates for alcohol intake on dementia, for five alcohol strata, excluding nondrinkers.

| **Stratum** | **Mean alcohol exposure, DPW** | **Beta** | **LCI** | **UCI** | **P value** |
| --- | --- | --- | --- | --- | --- |
| 1 | 0.86 | 1.03 | -0.89 | 2.95 | 0.29 |
| 2 | 1.67 | 0.55 | 0.16 | 0.94 | 5.96E-03 |
| 3 | 3.51 | 0.21 | 0.04 | 0.39 | 0.02 |
| 4 | 7.10 | 0.03 | -0.07 | 0.13 | 0.56 |
| 5 | 16.09 | 0.08 | 0.01 | 0.15 | 0.02 |

Estimates generated using the doubly-ranked nonlinear Mendelian randomization method, in European ancestry unrelated participants of Million Veteran Program, excluding nondrinkers. Adjusted for: age, age^2^, sex, and top ten principal ancestry components. Abbreviations: DPW – drinks per week, LCI – lower confidence interval, UCI – upper confidence interval.

#### Table S26: Negative control - age.

| **Stratum** | **Mean alcohol exposure, DPW** | **Beta** | **LCI** | **UCI** | **P value** |
| --- | --- | --- | --- | --- | --- |
| 1 | 0.06 | 0.85 | -8.61 | 10.31 | 0.86 |
| 2 | 0.38 | -0.72 | -2.50 | 1.06 | 0.43 |
| 3 | 1.34 | 0.10 | -0.42 | 0.62 | 0.71 |
| 4 | 3.86 | 0.17 | -0.06 | 0.39 | 0.14 |
| 5 | 11.66 | 0.05 | -0.06 | 0.16 | 0.37 |

Estimates generated using the doubly-ranked nonlinear Mendelian randomization method, in European ancestry unrelated participants of Million Veteran Program. Associations were generated using linear regression, controlling for sex and top ten ancestry principal components. Abbreviations: DPW – drinks per week, LCI – lower confidence interval, UCI – upper confidence interval. N=313,873 (16,932 dementia cases) were included in the analyses.

#### Table S27: Negative control - sex.

| **Stratum** | **Mean alcohol exposure, DPW** | **Beta** | **LCI** | **UCI** | **P value** |
| --- | --- | --- | --- | --- | --- |
| 1 | 0.06 | 4.64 | -14.35 | 23.62 | 0.63 |
| 2 | 0.38 | -0.10 | -0.70 | 0.50 | 0.74 |
| 3 | 1.34 | -1.27E-03 | -0.17 | 0.17 | 0.99 |
| 4 | 3.86 | -0.02 | -0.10 | 0.06 | 0.61 |
| 5 | 11.66 | 0.02 | -0.03 | 0.07 | 0.40 |

Estimates generated using the doubly-ranked nonlinear Mendelian randomization method, in European ancestry unrelated participants of Million Veteran Program (N= 313,873). Associations were generated using logistic regression, controlling for age, age^2^ and top ten ancestry principal components. Abbreviations: DPW – drinks per week, LCI – lower confidence interval, UCI – upper confidence interval. N=313,873 (16,932 dementia cases) were included in the analyses.
